# Epigenomics, Genomics, Resistome, Mobilome, Virulome and Evolutionary Phylogenomics of Carbapenem-resistant *Klebsiella pneumoniae* clinical strains

**DOI:** 10.1101/2020.06.20.20135632

**Authors:** Katlego Kopotsa, Nontombi M Mbelle, Osei Sekyere John

**Author notes:** Address correspondence to Dr. John Osei Sekyere, Department of Medical Microbiology, University of Pretoria, South Africa. **Tweet:** Epigenomic and genomic factors influence the resistome, mobilome, virulence and epidemiology of *K. pneumoniae*. They harbour multiple resistance genes, insertion sequences, transposons, prophages and restriction modification systems on mobile plasmids, influencing their genomic plasticity and threatening antimicrobial and bacteriophage chemotherapy.

## Abstract

**Background:** Carbapenem-resistant *Klebsiella pneumoniae* (CRKP) remains a major clinical pathogen and public health threat with few therapeutic options. The mobilome, resistome, methylome, virulome and phylogeography of CRKP were characterised.

**Methods:** CRKP collected in 2018 were subjected to antimicrobial susceptibility testing, screening by multiplex-PCR, genotyping by Repetitive Element Palindromic-Polymerase Chain Reaction (REP-PCR), plasmid size, number, incompatibility, and mobility analyses, and PacBio’s SMRT sequencing (n=6).

**Results & conclusion:** There were 56 multidrug-resistant CRKP, having *bla*_OXA-48_-like and *bla*_NDM-1/7_ carbapenemases on self-transmissible IncF, A/C, IncL/M and IncX_3_ plasmids endowed with prophages, *traT*, resistance islands and type I and II restriction modification systems (RMS). These plasmids were of close evolutionary relationship to several plasmids globally whilst the strains also clustered with several global clades, evincing transboundary horizontal and vertical dissemination. Reduced susceptibility to colistin occurred in 23 strains. Common clones included ST307, ST607, ST17, ST39, and ST3559. IncFII_k_ virulent plasmid replicon was present in 56 strains. The six strains contained at least 41 virulence genes and four different K- and O-loci types: KL2, KL25, KL27, KL102, O1, O2, O4 and O5. Types I, II, and III RMS, conferring m6A (G**<u>A</u>**TC, G**<u>A</u>**TGNNNNNNTTG, CA**<u>A</u>**NNNNNNCATC motifs) and m4C (C**<u>C</u>**WGG) modifications on chromosomes and plasmids, were found.

There is plasmid-mediated, clonal, and multiclonal dissemination of *bla*_OXA-48_-like and *bla*_NDM-1_ in South Africa, mirroring international epidemiology of similar clones and plasmids. Plasmid-mediated transmission of RMS, virulome and prophages influence bacterial evolution, epidemiology, pathogenicity, and resistance, threatening infection treatment. RMS influence on antimicrobial and bacteriophage therapy needs urgent investigation.

**Highlights/Importance:** *K. pneumoniae* is a major pathogen implicated in numerous nosocomial infections. Worryingly, we show that *K. pneumoniae* isolates from South Africa, Africa and globally are endowed with rich resistomes and mobilomes that make them almost pandrug resistant. The isolates in this study contained rich virulomes and prophages on both chromosomes and plasmids, with close evolutionary kith or kin to other plasmids identified worldwide. There was a rich diversity of restriction modification systems that regulate virulence, transcription, and plasmid mobility in bacteria, facilitating the epidemiology, resistance, pathogenicity and genomic evolution of the strains, and threatening antimicrobial and bacteriophage therapy.

## Introduction

*Klebsiella pneumoniae* is an encapsulated, non-motile, Gram-negative bacterium first isolated from the lung of a demised patient who was suffering from pneumonia in 1882 ^1^. These bacteria are known to colonize human gastrointestinal (GI) tract and oropharynx mucosal surfaces ^2^. To date, *K. pneumoniae* causes most nosocomial infections, accounting for 3% to 8% of all reported nosocomial infections ^3^. These infections are specifically a problem in the elderly, immunocompromised patients and neonates; but less frequently, *K. pneumoniae* infections such as sepsis and pneumonia are community-acquired ^4^.

Antimicrobial resistance (AMR) in bacteria such as *K. pneumoniae* has become a major public health concern worldwide, fuelled by misuse and overuse of antibiotics, which increases the evolution of AMR genes (ARGs) and antimicrobial-resistant bacteria^5^. Resistance may be intrinsic i.e., acquired through mutations, and/or transferred horizontally from one bacterium to another through mobile genetic elements (MGEs) ^6^. Among the various AMR mechanisms, acquired resistance through MGEs is most reported ^7^. Acquisition of antimicrobial-inactivating enzymes and efflux pump systems are important in the development of multi-drug resistant (MDR) in Enterobacterales, including *K. pneumoniae* ^8,9^, with the resistance-nodulation-division (RND) family of efflux pumps being responsible for ejecting charged and amphiphilic antimicrobials such as aminoglycosides, β-lactams and fluoroquinolones ^9,10^. The use of β-lactams over the years has resulted in widespread escalation of β-lactamases, including carbapenemase-producing *K. pneumoniae* ^11,12^, resulting in increased treatment failure, morbidity, and mortality ^13^.

Carbapenemases are categorised into three classes, class A (e.g. KPC, SME, IMI, and GES), class B (e.g. NDM, VIM, and IMP), and class D (OXA-48-like) ^14,15^. Carbapenemases are capable of slightly and/or completely hydrolysing β-lactams, including “last resort” carbapenems^16^. Class B carbapenemases, particularly *bla*_NDM_ genes, have been reported to be more potent than the other groups and cannot be inhibited by commercially available β-lactamase inhibitors such as clavulanic acid, tazobactam, or sulbactam ^17,18^.

Mobile genetic elements such as plasmids, transposons, prophages and integrons play a major role in the acquisition and dissemination of antimicrobial resistance genes (ARGs) in carbapenem-resistant strains ^12,19–21^. Among these are large conjugative plasmids that have been associated with horizontal gene transfer (HGT) of carbapenemases between and within Gram-negative bacteria^7^. These plasmids have been reported in *K. pneumoniae* strains and are associated with multiple replicon groups such as IncF, A/C, L/M, N, and X ^19^. IncF replicon plasmids are the most predominant and are mainly reported to carry the *bla*_KPC_ and *bla*_NDM_ genes in the United States, Canada, Greece, South Africa and Taiwan ^12,19,22,23^. Besides resistance, plasmids are being increasingly associated with the transmission of virulence factors, resulting in transmission of important pathogenic traits ^24–26^. Recently, it is increasingly being realised that restriction modification systems (RMS), comprising of restriction endonucleases (REs) and DNA methylases (MTases), regulate virulence, plasmid mobilization, bacterial immunity, DNA repair, transcription, replication, regulation and resistance in bacteria ^27–31^. These suggest the important role of the methylome in bacterial virulence, mobilome and resistome, which ultimately affect infectious diseases epidemiology. This study thus seeks to characterise these factors in this important pathogen.

## Results

### Clinical demographics and genome characteristics

The *K. pneumoniae* isolates were isolated from aspirates (n = 4), blood cultures (n = 17), catheter tips (n = 7), swabs (n = 11), tissue (n = 3) and urine (n = 14) (Tables 1 & S1), and were submitted to the referral laboratory from six hospitals and centres including Kalafong hospital (n = 10), Mamelodi hospital (n = 1), Olievenhoutbosch clinic (n = 1), Steve Biko academic hospital (n = 36), Tembisa hospital (n = 5) and Tshwane rehabilitation centre (n = 3). The study population consisted of males (58.9%) more than females (39.3%) and results were not available for one participant (Table S1).

### Antibiogram-resistome associations

Of the 60 *K. pneumoniae* with reduced susceptibility to carbapenems by VITEK®, carbapenem resistance could only be confirmed in 56 isolates by the MicroScan system (Table S1). The isolates were multidrug-resistant (MDR), with a substantial number being extensively drug resistant (e.g. KP51, KP27, KP 42 etc.); a few showed pandrug resistance phenomes (e.g. KP56) (Fig. 1A & 1Bi). Almost all isolates showed reduced susceptibility to ertapenem (98.2%), followed by imipenem (66.1%), doripenem (50%) and meropenem (48.2%). Reduced susceptibility to colistin was also observed in 23 (41.1%) isolates. Among all the tested antibiotics, the isolates were most susceptible to amikacin (82.1%), fosfomycin (82.1%), tigecycline (76.8%), and levofloxacin (60.7%) (Figure 1). The most prevalent carbapenemase detected by PCR was *bla*_OXA-48_ (65%), followed by *bla*_NDM-1_ (29%). No *bla*_GES_, *bla*_KPC_, *bla*_IMP_ and/or *bla*_VIM_ were detected (Fig. 1Bii).

**Figure 1.**
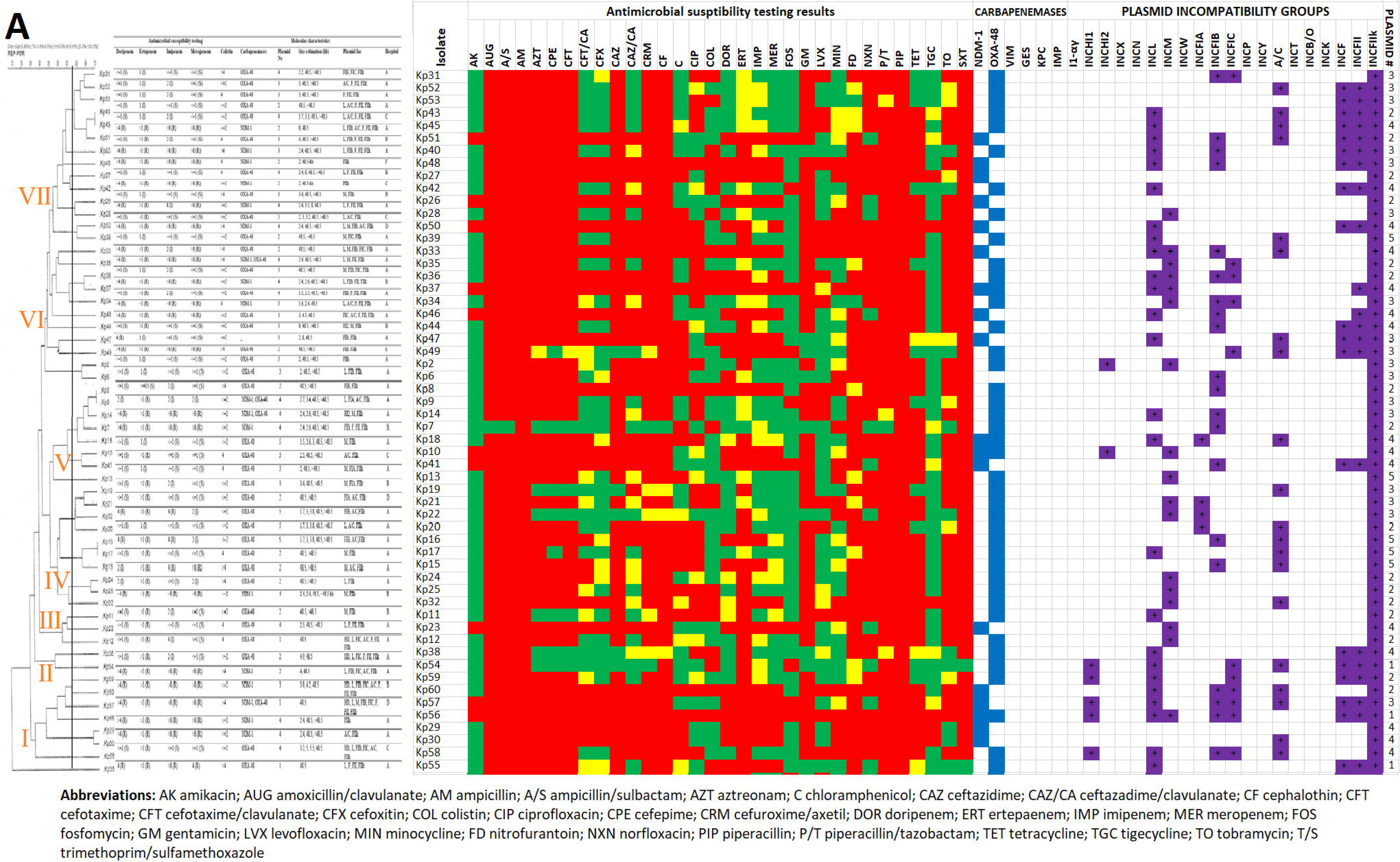
Antibiotic susceptibility patterns, carbapenemase genes and plasmid characteristics of 56 clinical *K. pneumoniae* isolates arranged according to their clustering patterns on a REP-PCR dendrogram. The isolates clustered into 7, based on their gel patterns. Antibiotic resistance is shown as red, intermediate resistance is shown as yellow and susceptibility is shown as green checks. Carbapenemase genes are shown as blue and plasmid replicons are shown as violet/mauve. The number of plasmids is shown in the last column (**A**). The frequency of resistance to each profiled antibiotic is shown as either resistant (red), intermediate resistance (yellow), and susceptible (green); the frequency of resistance shows MDR phenotypes among the collection (**i**). Most of the isolates harboured OXA-48 (65%) compared to NDM-1 (29%) whilst a minority had both genes (6%) (**ii**). IncF plasmid replicons were the commonest plasmid types, being found in all the isolates; IncL, A/C, and IncM were also common types (**iii**). A gel image of the REP-PCR patterns of the various isolates similar band patterns among the isolates (**iv**). A conjugation experiment with a receiver Escherichia coli showed that 20 out of the 26 meropenem-resistant isolates had mobile plasmids, as shown in the gel image, confirming the reception of carbapenemases by the hitherto susceptible *E. coli* J53-A^r^ (sodium-azide resistant) strain (**v**), (**B**).

*bla*_OXA-48_-containing isolates were mostly non-resistant to the carbapenems except to ertapenem in selected cases (e.g., KP44, KP40, KP39, KP2 etc.) and to the other carbapenems in a few cases (e.g., KP36, KP49, KP8, and KP55). Contrarily, all isolates harbouring *bla*_NDM-1_ were all resistant to all the carbapenems. Further, isolates containing both *bla*_OXA-48_ and *bla*_NDM-1_ genes (e.g. KP10 and KP56) were also resistant to all carbapenems except KP18, which was only resistant to ertapenem and intermediate resistant to the rest (Table S1 and Fig. 1).

The resistome of the sequenced isolates were similar and mostly agreed with their respective antibiograms except for fosfomycin to which all the isolates were susceptible even though they all harboured the *fosA* gene. The six isolates carried *bla*_OXA-181_ (n = 2), *bla*_OXA-48_ (n = 1), *bla*_NDM-1_ (n = 2), and *bla*_NDM-7_ (n =1) alongside other resistance determinants causing resistance to aminoglycosides (except amikacin)[*aac(3)-lla, aac(6’)-lb-cr, aadA16, aph(3’)-lb, aph(6)-ld, rmtC*], quinolones [*aac(6’)-lb-cr, oqxA, oqxB*, qnrB1, *qnrS1*], β-lactams (*bla*_OXA-1_, *bla*_CTX-M-15_, *bla*_SHV_, *bla*_TEM-1B_), tetracycline (*tetA*), sulphonamides (*sul1, sul2*), trimethoprim (*dfrA14*/*27*), phenicol (*catB3*/ *catA2*), and fosfomycin (*fosA*/*A7*). KP29 had no *tet*(A) but was resistant to the tetracyclines; *bla*_SCO-1_ was found in only this isolate. The 16S rRNA methyltransferase, rmtC, was found in only KP10 and KP33; yet, KP33 was susceptible to amikacin while KP10 was resistant. Chromosomal mutations in parC (S104I) and gyrA (S83I), conferring high-level fluoroquinolone MICs, were only seen in KP8. Nevertheless, the other isolates, which had no mutations, were also resistant to the fluoroquinolones (KP29 was susceptible to ciprofloxacin and levofloxacin) (Table S1 & Supplemental data 2).

Mobile colistin resistance determinants, *mcr-1* to -*10*, were not identified. although colistin resistance was recorded in three isolates. Particularly, KP10 was susceptible to colistin, had no *ccrB* gene and had M66I mutation in pmrA whilst KP15, which was also susceptible to colistin, also had no mutations in *pmrAB, phoQP, mgrB* and *kpnEF*, and had no *ccrB*. Although KP29 had mutations in *ccrB* (D189E), *kpnE* (K112Q) and *pmrA* (E57G), it was also susceptible. *OqxAB, fosA* and *bla*_SHV_ were all found on the chromosomes (Supplemental data 2).

### REP-PCR phylogenetics

The Repetitive extragenic palindromic (REP) – PCR dendrogram showed seven main clusters, with little or no similarities in the antibiogram, resistome or mobilome of isolates in the same clusters (Fig. 1); i.e., they were not clonally specific. Notwithstanding, isolates from the same hospital/ward had much similar antibiograms, resistomes and mobilomes (Fig. 1 & Table S1). The six sequenced isolates were collected from three different hospitals. Four isolates were from the same hospital but different wards and collection sites including ward 4, urine (Kp10); vascular surgery ward 4, catheter tip (Kp15); neurology ward, swab (Kp29); and high-care multidiscipline ward, urine (Kp33) (Table S1). Five different sequence types (STs) were identified among the isolates including ST39, ST307, ST607, ST17, and ST3559. Isolates Kp10 and Kp33, carrying *bla*_NDM-1_, both belonged to sequence type-39 (ST39), although they clustered differently on the dendrogram.

### Mobilome

Plasmid characterization by gel electrophoresis revealed that most isolates (n = 17) carried four plasmids, followed by 16 isolates with two plasmids, 15 isolates with three plasmids, five isolates with five plasmids, and three isolates with only one plasmid. Plasmids sizes ranged from 1.4-kb to >48.5-kb (Fig. 1; Table S1). Eleven plasmid replicon groups were identified in all the isolates (n=56), which were all positive for IncFII_k_ (virulent plasmid). Majority of the isolates were positive for IncF (FII, FIB, FIC, FIB), IncL, A/C, and IncM plasmids, while only a few isolates were positive for IncHI1 and IncHI2 (Fig. 1B; Table S1). Multi-replicons were reported in 75% (n = 42) of the tested isolates. Two isolates showed the highest multi-replicon combination: one *bla*_OXA-48_-producer and one *bla*_NDM-1_-producer had six and seven replicon groups, respectively (Figure 5).

Conjugation experiments were successful on 20 out of the 26 meropenem-resistant *K. pneumoniae* isolates; 20 donor strains transferred their plasmids to *E. coli* J53-A^r^ strain. Among the 20 transferred plasmids, 16 were positive for the *bla*_NDM-1_ gene, followed by three *bla*_OXA-48_ and one isolate with both *bla*_NDM-1_ and *bla*_OXA-48_ genes (Fig. 1B). Sequencing analyses of six isolates showed that five had two major plasmids whilst KP29 had three, which were different from the numbers obtained from the gel electrophoresis analyses, except for KP8 (Fig. 1-6; Table S1); fragments (contigs) of these plasmids were found but had no replicon genes (Table S1). The sizes obtained from the sequencing analyses were also different from that obtained from the gel electrophoresis analyses: KP8 [pKP8.6_CTX-M-15 (198, 548bp) and pKP8.12_OXA181 (51, 479bp)]; KP10 [pKP10.8_NDM-1 (101,574bp) and pKP10.4_OSEI (181, 384bp)]; KP15 [pKP15.4_KATLEGO (311, 287bp) and pKP15.12_OXA-181 (153, 467bp)]; KP29 [pKP29.8_CTXM-15 (237, 044bp), pKP29.11_NDM-7 (45, 184bp) and pK29.13_MBELLE (83, 675bp)]; KP32 [pKP32.5_OXA-48 (85, 104bp) and pKP32.3_CTXM-15 (247, 163bp)]; and KP33 [pKP33.8_NDM-1 (101,574bp) and pKP33.4_OSEI (181, 384bp)] (Fig. 2-6 & S1-S6).

**Figure 2.**
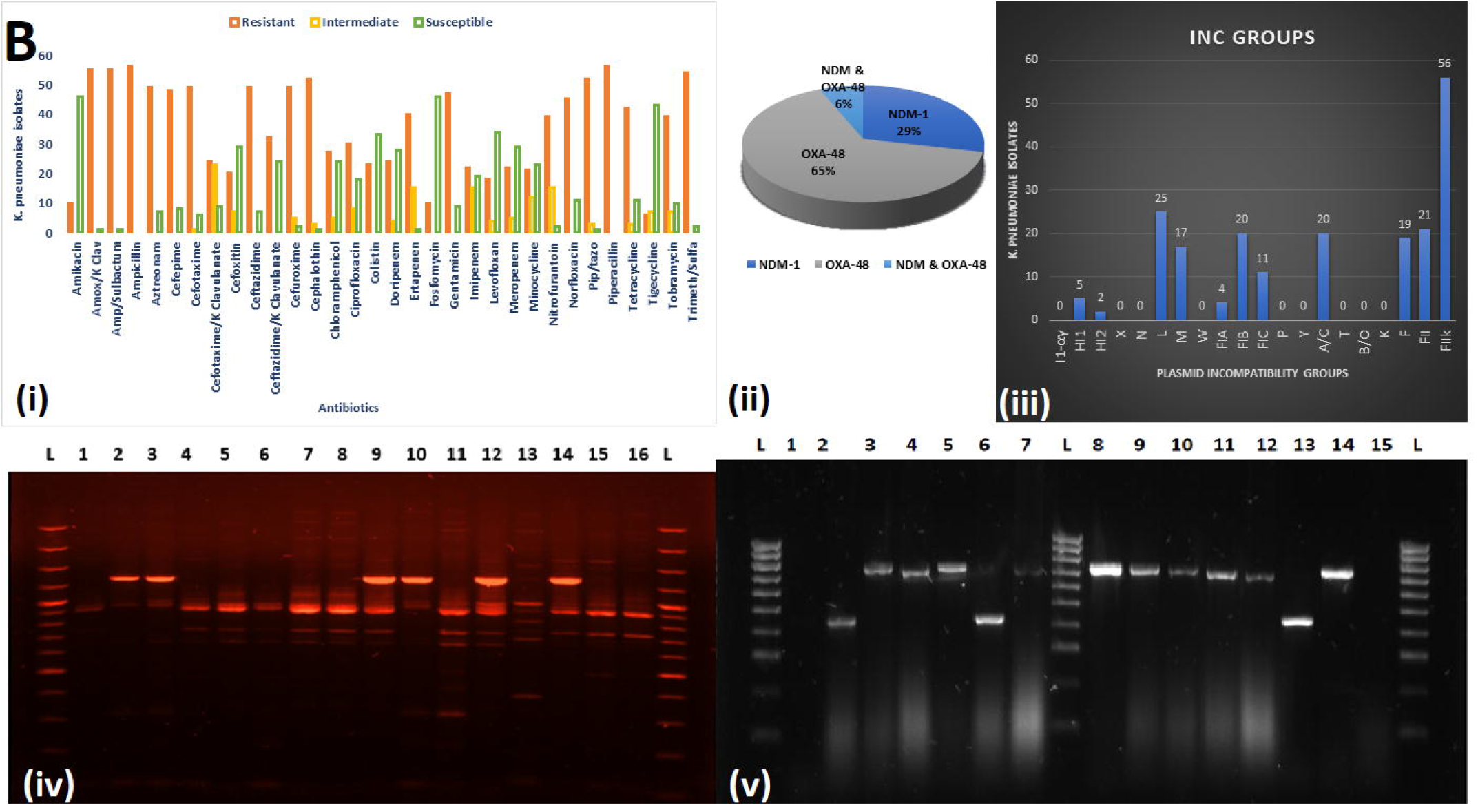
Graphical annotation and comparative alignment of pKP8.12_OXA181. The resistance genes (light green-coloured arrows), transposons (orange-coloured arrows), insertion sequences (orange-coloured arrows), integrons (red-coloured arrows), resolvases (red-coloured arrows), and recombinases/integrases (red-coloured arrows) on the plasmid is shown with their orientation (direction of arrow), synteny and immediate environment. Other genes with unknown functions are hidden to make the image less cluttered. A circular version of the plasmid is shown in (**A**). A linear version of the plasmid and its alignment with other similar plasmids (linear arrows with yellow highlighted background) are shown in (**B**); regions of alignment are shown as red-filled portions whilst non-aligned areas are shown as empty arrows. Enlarged section of the plasmid focusing on the resistance genes (genomic resistance island) are shown in (**C**). This plasmid (VXJB01000012) contains OXA-181.

Three of these plasmids viz., pKP32.5_OXA-48, pKP29.9_CTXM-15, and pK29.13_MBELLE, had overlapping ends, making them completely circularised whilst the remaining were partial with no overlapping ends (Fig. 2-6). *Bla*_NDM-1_ was present on pKP10.8_NDM-1 (IncFII_(yp)_) and pKP33.8_NDM-1 (IncFII_(yp)_) alongside *ble, rmtC, sul1* and cytosine MTase within *IS*1/5/3 and *ISKpn*26 ISs (insertion sequences) (Fig. 3 & S6.1). *Bla*_NDM-7_ was found on pKP29.11_NDM-7 (IncX_3_) alongside *ble* within *IS*5, *IS*26, *ISKox*3 and a resolvase (Fig. 5). *Bla*_OXA-181,_ alongside *ereA* and *QnrS1*, was found on pKP8.12_OXA181 (ColKP3 and IncX_3_) within composite *Tn*3-like transposons and *IS*26, *ISKpn*19 and *ISKox*3 ISs; *bla*_OXA-181_ was also found on pKP15.12_OXA-181 (IncU, IncC and colKP3) with *ereA, Qnrs1, aac(6’)-Ib3, mph*(A), *tet*(A), *aph(3”)-Ib, aph(6)-Id, sul2*, and adenine and cytosine MTases bracketed by *Tn*3 composite transposons, *IS*26, *IS*3000, *IS*6100, and *IntI*1 class 1 integron (Fig. 2 & 4). *Bla*_OXA-48_ was found on pKP32.5_OXA-48 (IncL) alone within *IS10*A, *IS*1 and *IS*4 ISs (Fig. 6).

**Figure 3.**
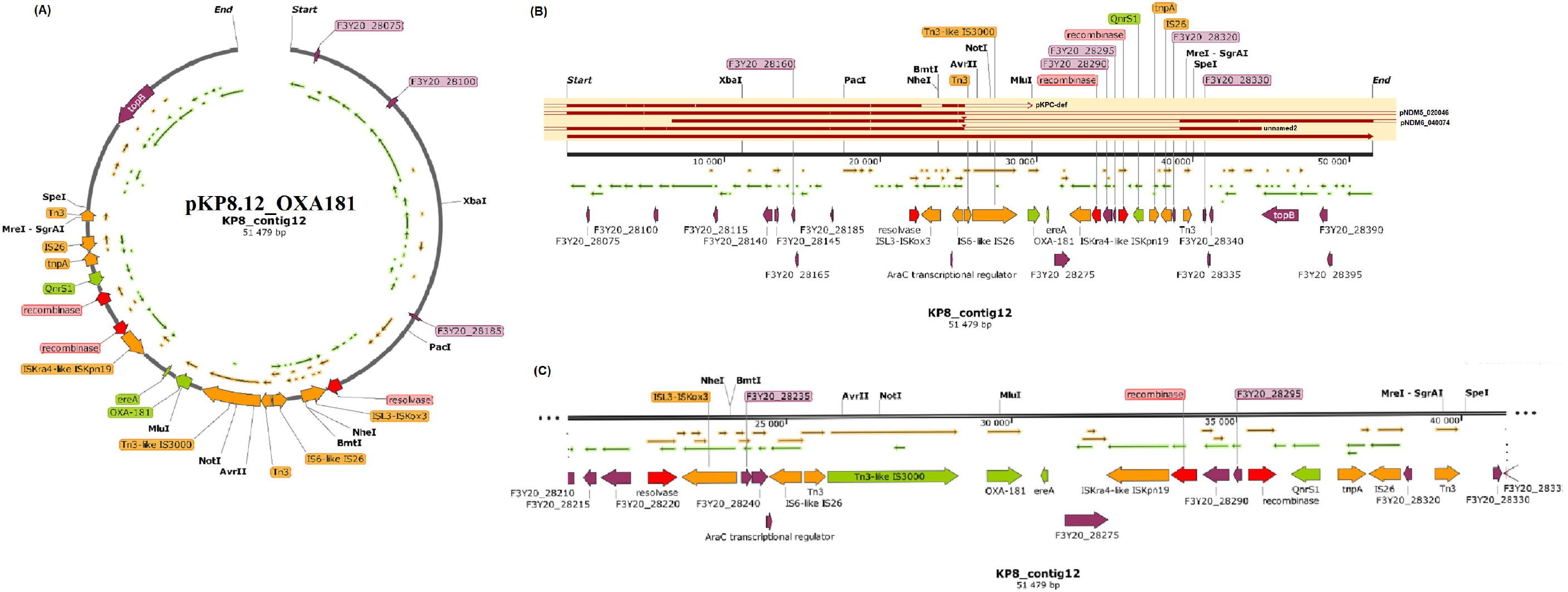
Graphical annotation and comparative alignment of pKP10.8_NDM-1. The resistance genes (light green-coloured arrows), replicase genes (blue-coloured arrows), methyltransferases/restriction endonucleases (rose-coloured arrows), transposons (orange-coloured arrows), insertion sequences (orange-coloured arrows), integrons (red-coloured arrows), resolvases (red-coloured arrows), and recombinases/integrases (red-coloured arrows) on the plasmid is shown with their orientation (direction of arrow), synteny and immediate environment. Other genes with unknown functions are hidden to make the image less cluttered. A circular version of the plasmid is shown in (**A**). A linear version of the plasmid and its alignment with other similar plasmids (linear arrows with yellow highlighted background) are shown in (**B**); regions of alignment are shown as red-filled portions whilst non-aligned areas are shown as empty arrows. Enlarged section of the plasmid focusing on the resistance genes (genomic resistance island) are shown in (**C**) and (**D**). This plasmid (VXJA01000008) contains NDM-1.

*Bla*_CTX-M-15_ (bracketed by *ISEc*9, Tn3 transposon, IS1 and IS26) and *bla*_TEM-1B_ (bracketed by a recombinase/integrase, IS91, *aph(6)-Id, aph(3”)-Ib, sul2*, IS5075, IS5, IS91) were found in close synteny on the same plasmids in all the isolates; *bla*_TEM-1B_ genetic environment only differed slightly in pKP29.8_CTXM-15 (Fig. S1-S6). Adenine and cytosine MTases were also found the same plasmids as *bla*_CTX-M-15_ and *bla*_TEM-1B_, within ISs and transposons. *Bla*_OXA-1_ (with *aac(3)-IIa, catB3, aac(6’)-Ib-cr5, IS*26, *TnAs*1, a relaxase, *tet*(A-R) and *Tn*3) and *bla*_SCO-1_ (found within ISs, MTase, and a recombinase on pKP29.8_CTXM-15) were only found on four and one plasmids respectively; notably, *bla*_OXA-1_ and its rich genetic environment of resistance genes and ISs were on the sample plasmids as *bla*_CTX-M-15_ and *bla*_TEM-1B,_ albeit quite distant from the latter, forming a resistance island. Thus, *bla*_CTX-M-15_ and *bla*_TEM-1B_ formed a genomic resistance island with their immediate genetic environment of rich ISs and transposons just as *bla*_OXA-1._ Furthermore, other antibiotic (*dfrA14, QnrB, aadA, aac(6’)-Ib-cr5, arr-3, catA, qacE*Δ*1, sul1*) and mercury (*merABCDETPR*) resistance genes, *tolA* efflux pump, NH_2_ restriction endonucleases and MTases were also found on ISs, transposons and mainly, on *IntI*1 class 1 integrons (Fig. S1-S6).

Complete and partial prophage DNA were identified on both chromosomes and plasmids, including the Klebsi_phiKO2, Cronob_ENT47670, Edward_GF_2, Pectob_ZF40, Phage_Gifsy, and different variants of Salmon (6), Entero (2), Escher (3) (Supplemental data 3 and Fig. S7-S11). Complete and incomplete prophages were found in the plasmids of all the isolates. KP15 alone had four complete prophages on two plasmids (pKP15.4_KATLEGO and pKP15.12_OXA-181) whilst the single complete prophages were found on single plasmids in the other isolates. Furthermore, complete and incomplete prophages were also found on chromosomes in these isolates (Fig. S7-S11). The same prophages were also found in different isolates (Supplemental data 3).

### Plasmid evolutionary epidemiology

A phylogenetic analysis of the plasmids obtained in this study, shows that some of these plasmids, which belonged to different replicon groups and harboured different ARGs from different isolates, were evolutionary closer to each other (Fig. 7). For instance, pKP32.3_CTXM-15 and pKP8.6_CTX-M-15 were of different sizes and from different isolates, but had similar ARGs and replicon types (IncFII(K) and IncFIB(K)) clustered together on the same branch. The same observation was made for pKP10.8_NDM-1 and pKP33.8_NDM-1 as well as for pKP10.4_OSEI and pKP33.4_OSEI, which harboured the same ARGs and replicon types in two different isolates of the same clone (ST39). Contrarily, IncX_3_ *bla*_NDM-7_-bearing pKP29.11_NDM-7 plasmid clustered closely with pKP29.9_CTXM-15, an IncFIB(K) and IncFII(K) plasmid, albeit all were from the same isolate. Nevertheless, pKP15.4_KATLEGO and pK29.13_MBELLE, which had IncR and IncFIA(HI1) replicons, clustered separately. IncX_3_-ColKP3 pKP8.12_OXA181plasmid was distant from all the other plasmids (Fig. 7). Mauve alignment of these plasmids with other closely related plasmids from GenBank showed similarities in sequence identity and synteny as well as sequence rearrangements (Fig. S1-S6).

**Figure 4.**
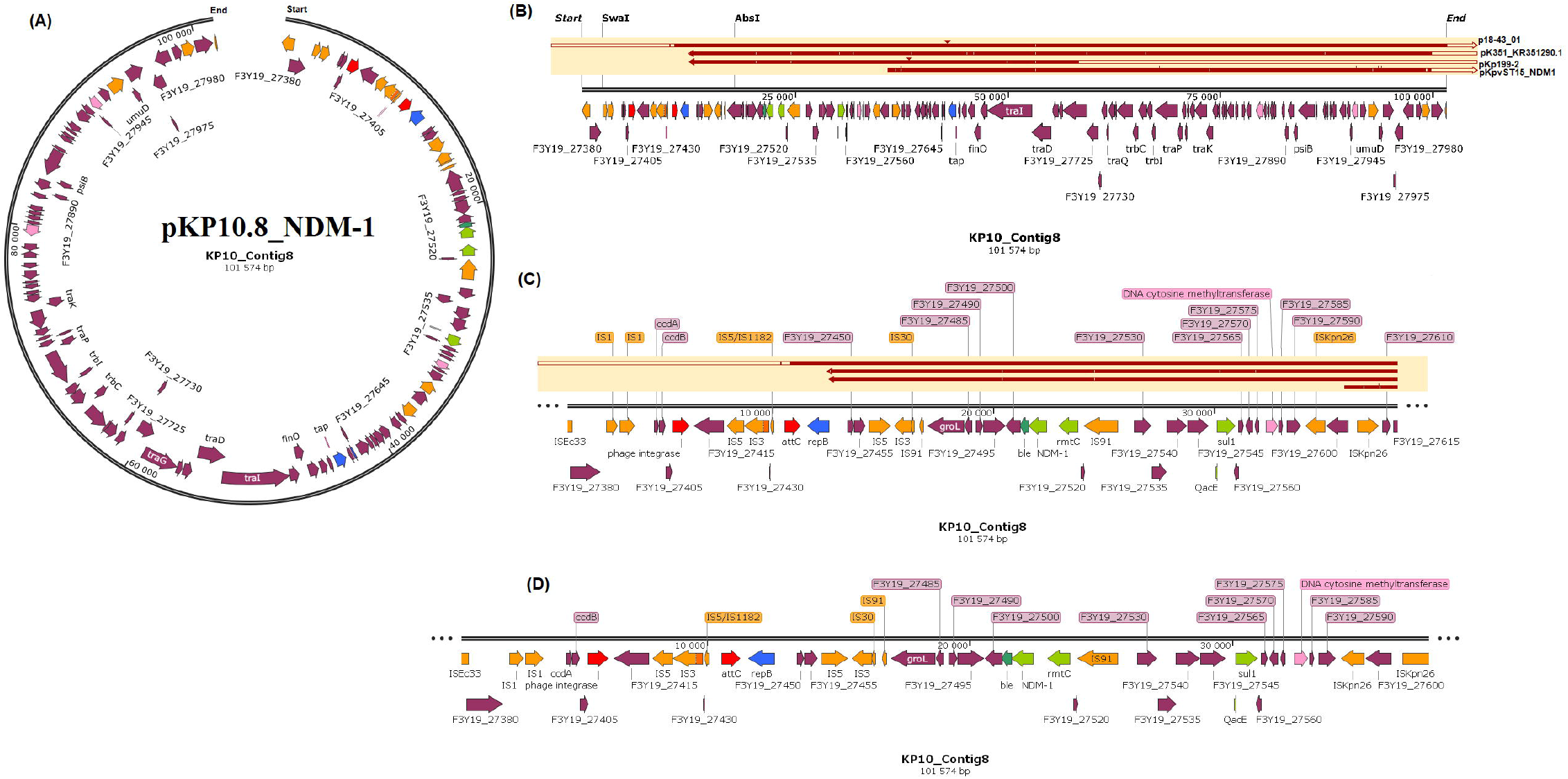
Graphical annotation and comparative alignment of pKP15.12_OXA-181. The resistance genes (light green-coloured arrows), transporter genes (deep green-coloured arrows), methyltransferases/restriction endonucleases (rose-coloured arrows), transposons (orange-coloured arrows), insertion sequences (orange-coloured arrows), integrons (red-coloured arrows), resolvases (red-coloured arrows), and recombinases/integrases (red-coloured arrows) on the plasmid is shown with their orientation (direction of arrow), synteny and immediate environment. Other genes with unknown functions are hidden to make the image less cluttered. A circular version of the plasmid is shown in (**A**). A linear version of the plasmid and its alignment with other similar plasmids (linear arrows with yellow highlighted background) are shown in (**B**); regions of alignment are shown as red-filled portions whilst non-aligned areas are shown as empty arrows. Enlarged section of the plasmid focusing on the resistance genes (genomic resistance island) are shown in (**C**). This plasmid (VXIZ01000012) contains OXA-181.

**Figure 5.**
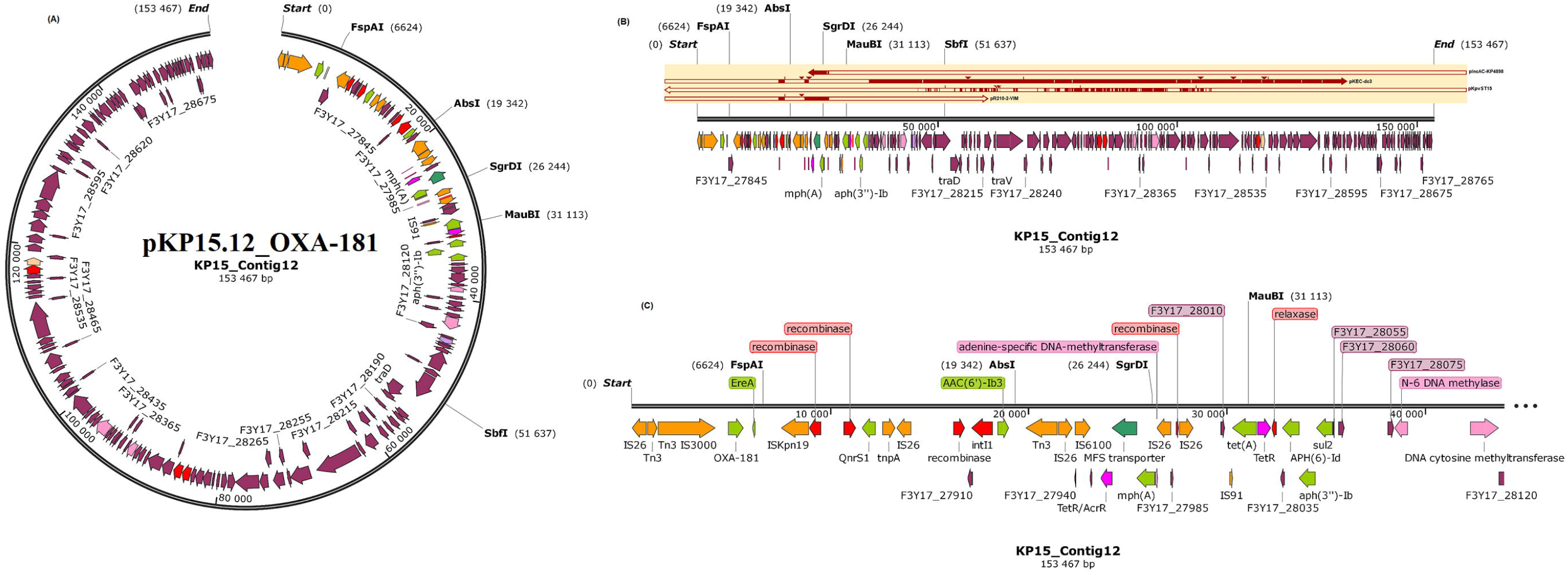
Graphical annotation and comparative alignment of pKP29.11_NDM-7. The resistance genes (light green-coloured arrows), methyltransferases/restriction endonucleases (rose-coloured arrows), transposons (orange-coloured arrows), insertion sequences (orange-coloured arrows), integrons (red-coloured arrows), resolvases (red-coloured arrows), and recombinases/integrases (red-coloured arrows) on the plasmid is shown with their orientation (direction of arrow), synteny and immediate environment. Other genes with unknown functions are hidden to make the image less cluttered. A circular version of the plasmid is shown in (**A**). A linear version of the plasmid and its alignment with other similar plasmids (linear arrows with yellow highlighted background) are shown in (**B**); regions of alignment are shown as red-filled portions whilst non-aligned areas are shown as empty arrows. Enlarged section of the plasmid focusing on the resistance genes (genomic resistance island) are shown in (**C**). This plasmid (VXIY01000011) contains NDM-7.

**Figure 6.**
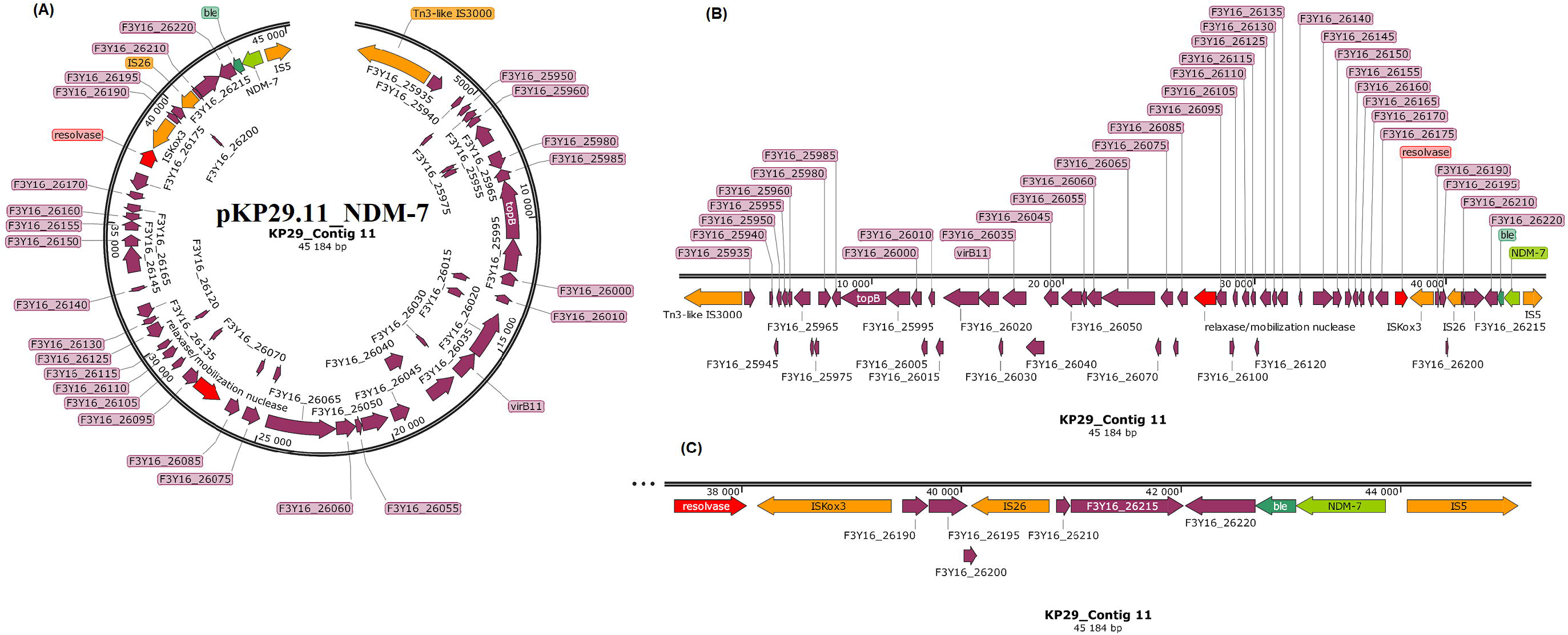
Graphical annotation and comparative alignment of pKP32.5_OXA-48. The resistance genes (light green-coloured arrows), replicase genes (blue-coloured arrows), methyltransferases/restriction endonucleases (rose-coloured arrows), transposons (orange-coloured arrows), insertion sequences (orange-coloured arrows), integrons (red-coloured arrows), resolvases (red-coloured arrows), and recombinases/integrases (red-coloured arrows) on the plasmid is shown with their orientation (direction of arrow), synteny and immediate environment. Other genes with unknown functions are hidden to make the image less cluttered. A circular version of the plasmid is shown in (**A**). A linear version of the plasmid and its alignment with other similar plasmids (linear arrows with yellow highlighted background) are shown in (**B**); regions of alignment are shown as red-filled portions whilst non-aligned areas are shown as empty arrows. Enlarged section of the plasmid focusing on the resistance genes (genomic resistance island) are shown in (**C**). This plasmid (VXIX01000005) contains OXA-48.

**Figure 7.**
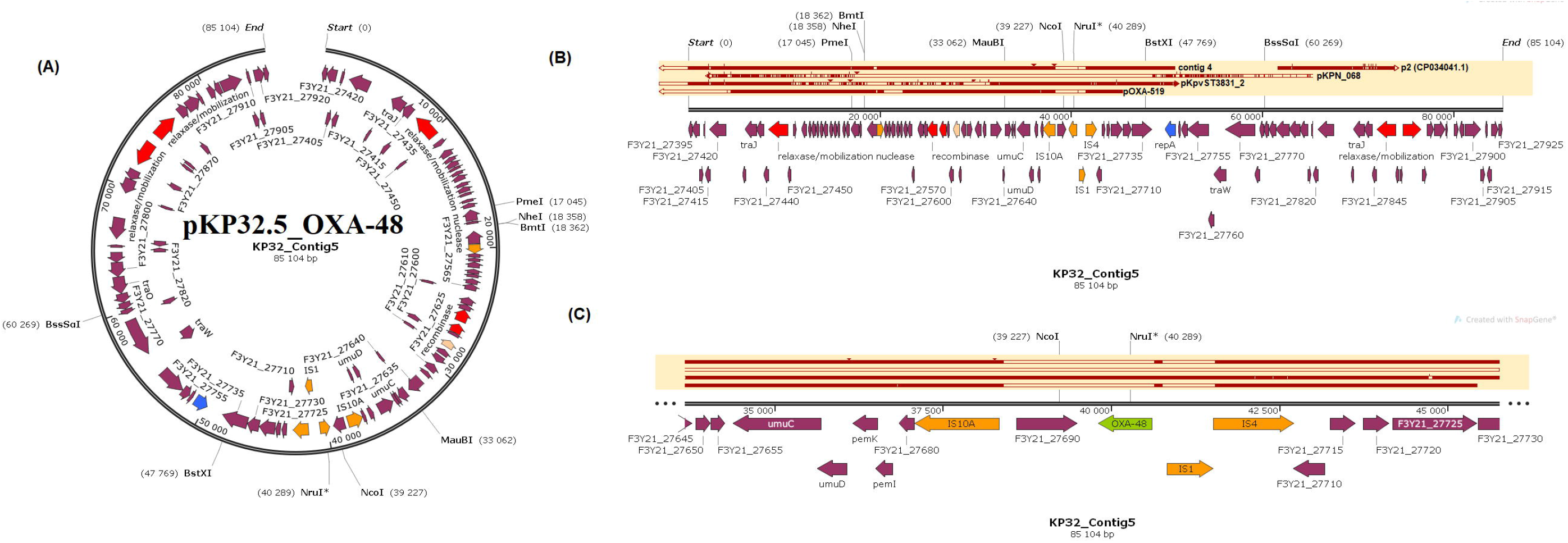
Global phylogenetic, resistome and phylogeographic characteristics of carbapenemase-hosting plasmid incompatibility types. The phylogenetic relationship between the various plasmids obtained in this study are shown in (**A**), with those bearing CTX-M and/or TEM ESBLs labelled with green text whilst those bearing OXA-181, OXA-48 and NDM-1/7 are labelled with purple, blue and red text respectively; branches of the same clade have the same highlights. The resistomes of carbapenemase-bearing plasmids isolated worldwide are shown in (**B**) under various plasmid replicon types; the various plasmid types harbouring carbapenemases and associated resistance genes are also shown. The geographical distribution of the plasmid incompatibility groups are shown in (**C**), and it is evident that most of these plasmids were isolated from North America, Brazil, Columbia, Europe, South Africa, Kenya, Middle East, South-Eastern Asia and Australia.

Nucleotide BLAST analyses and comparative genomics of the carbapenemase-bearing plasmid genomes from this study, identified highly similar plasmids with the same replicon types, ARGS and host bacterial species from different countries globally; in a few cases, the replicon types and ARGS differed slightly, albeit the country and bacterial species differed substantially (Fig. 7 & S13; Table S2). In most cases, the plasmids from this study aligned most closely viz., more than 90% nucleotide sequence identity and coverage, with plasmids deposited on Genbank, which were from different countries (Table S2; Fig. S13). For instance, pKP8.12_OXA181, a multi-replicon plasmid, was closely aligned with 201 other plasmids of the same or similar replicon group (IncX/ColKP3) from different countries and bacterial hosts but having *bla*_OXA-181_ or *bla*_NDM_. Similarly, pKP10.8_NDM-1 was closely aligned to 31 other plasmids of the same replicon with *bla*_NDM_ from the USA, Canada, China, New Zealand, and Europe. Similar trends were also observed for pKP15.12_OXA-181 (aligned to 33 other IncC/U and ColKP3 plasmids of the same replicons from the US, China, Europe, and Vietnam, although they contained few carbapenemases), pKP29.11_NDM-7 (aligned to 231 other IncX_3_ plasmids worldwide that harboured *bla*_OXA-181,_ *bla*_KPC_ or *bla*_NDM_) and pKP32.5_OXA-48 (aligned with 59 other IncL plasmids worldwide that also harboured *bla*_OXA-48_). Notably, IncX_3_ plasmids harboured NDM whilst IncX_3_-ColKP3 plasmids harboured OXA-181 (Fig. S13).

Resistome analyses of the various plasmids show that IncF, IncN, A/C, ColRNAI/KP3, and IncH had more diverse ARGs repertoire compared to IncX, IncL, IncR and IncI (Fig. 7). *bla*_NDM_, *bla*_KPC_, *bla*_TEM_ *aac(6’)-Ib, aac(3)-II, aadA, dfrA, sul, qnrB/S*, and *rmtB/C* were commonly found on all the replicon types except IncX; however, *bla*_NDM,_ *bla*_KPC_ and *bla*_SHV_ were common on IncX plasmids. Indeed, *bla*_NDM_ was more concentrated on IncX and A/C plasmids than other plasmid types. A phylogenetic analysis of these plasmids could not be undertaken due to their inability to align substantially. Notably, most of these plasmids have been reported from the USA, Europe and South-East Asia, with few being reported from South America, Africa and Australia; South Africa and Brazil reported more plasmid replicon groups in Africa and South America respectively (Fig. 7 & S13).

### Virulome

A total of 51 virulence genes were found in the chromosomes of the six genomes, with *ccI* and *traT* being found on plasmids. KP8 and KP32 had the least set of virulence genes (n=40) whilst the other isolates had all 51 genes. Hypervirulent genes were absent in all the genomes (Table S3; Fig. S14.1-2). The capsule polysaccharide-based serotyping or the K-loci results showed four different serotypes among the sequenced isolates: KL2 (KP10 and KP33), KL25 (KP15 and KP29), KL27 (KP32) and KL102 (KP8). As well, the O-loci results also showed four O serotypes: O1v1 (KP10, KP33, and KP15), O2v2 (KP8), O4 (KP32), and O5 (KP29). As shown in Fig. S14.3-8, the K- and O-serotyping was not only clone-specific as different clones shared the same K and O serotypes.

### Methylome

Types I, II and III MTases were found in the isolates, with type II MTases being the most abundant followed by type I MTases; a single type III MTase (M.Kpn214ORFGP or M.Kpn30104ORFBP) was found chromosomally on all isolates with no known motif. As well, no motif was identified for the type I RMS in all the isolates and except for KP15 and KP29, all the type I RMS were found on chromosomes. A complete RMS consisting of REs, MTases and a specificity subunit, representing the *hsd*RMS operon, were only found on either chromosomes or plasmids (KP15 and KP29) of all but KP32 and KP33; the position of these RMS components on the chromosomes or plasmids show that they were in very close or overlapping synteny. KP32 had only type I REs with no MTases whilst KP33 had no specificity subunit. Except for KP10 and KP33, all the other isolates had very unique type I RMS, with R2.KpnLAUORFGP (found on only chromosomes) being the sole common RE in all isolates. Furthermore, the type I REs on the chromosomes of KP15 and KP29 were different from that on the plasmids. Notably, all the isolates had only single type I MTases on either plasmids or chromosomes except KP8, which had two. There were mutations observed in the type I REs (Table S4).

Type II RMS adenine (Dam) and cytosine (Dcm) MTases, which respectively binds to and methylate G**<u>A</u>**TC (all isolates), G**<u>A</u>**TGNNNNNCTG/CA**<u>A</u>**NNNNNNCATC (KP29 only) and C**<u>C</u>**WGG or C**<u>C</u>**NGG (all isolates) were identified. Nevertheless, type II MTases with no known motifs were also identified. Noteworthily, all Dams (with GATC motifs) were only found on chromosomes in all isolates whilst most Dcms (with CCWGG or CCNGG motifs) were mainly found on plasmids with a few exceptions: KP8 (*M*.*Kpn10PVDcmP*), KP10 and KP33 (*M*.*Kpn0718ORF12365P* and *M*.*Sfl2ORFAP*), KP15 (*M*.*Kpn3210ORFFP*), and KP29 (*M*.*KpnLAUORFBP* and *M*.*Kpn3210ORFFP*) and KP32 (*M*.*Kpn3210ORFFP, M*.*KpnNIH30Dcm* and *M*.*Sfl2ORFAP*) had few chromosomal Dcms. As well, all the type II REs were on plasmids and were not more than two per isolate compared to several unique MTases on either plasmids or chromosomes. The absence of REs on the chromosomes makes many of these Dcms orphans with no known motifs (Table S4). An interesting observation was the multiple copies of *M*.*Sfl2ORFAP* on both plasmids and chromosomes in single strains as well as its common presence in all the strains. No CCWGG motif was found by MotifMaker and GATGNNNNNCTG/CAANNNNNNCATC was only identified by MotifMaker but absent from REBASE. Notably, the type II RE and MTases on either plasmids or chromosomes shared the same C**<u>C</u>**WGG motif (Table S4).

All isolates, including both KP15 and KP29, had mostly m6A modifications (methylated adenines) i.e. resulted in N6-methyladenine (^6m^A); more than 92% of G**<u>A</u>**TC and 84% of G**<u>A</u>**TGNNNNNCTG motifs were found in KP15 and KP29, respectively. Moreover, m4C modification types (methylated cytosines) i.e., resulted in N4-methylcytosine (^4m^C), were also seen in all the isolates, albeit fewer than 6mA modifications (Table S4). Analyses of all the *K. pneumoniae* genomes (from Africa and globally) used in this study showed a higher prevalence of Dams than Dcms in their genomes (data not shown).

### Global phylogenomics, phylogeography and resistome

The genomes of the six isolates ranged from 5.6-5.9Mb, with five to 13 contigs, a GC content of 57 and coding sequences ranging from 5461 to 5629. The N50s varied widely between the isolates although the L50 was only between one and two (Table S1 & S5).

The six isolates belonged to five different clones that clustered differently, with KP10 and KP33 falling within ST39 (Fig. 8A). The isolates also clustered with other *K. pneumoniae* isolates from Durban, Ozwatini, Pretoria, and Pietermaritzburg (clades II, VI, VIII, IX and XII) out of 82 isolates and 13 clades. Same clones/clades were seen circulating within and between Pretoria (Gauteng Province), Ozwatini, Pietermaritzburg, and Durban (KwaZulu-Natal Province). The evolutionary trajectory of the strains, as shown on the tree (Fig. 8B), depicts that some of the clades emerged from other clades or from a common ancestor: for instance, clades IV and V share the same ancestor as clade III, albeit clades IV and V were mainly of ST152. Evidently, ST14 and ST15 as well as ST983 and ST17 were of very close evolutionary distance (Fig. 8B). *K. pneumoniae* ST101 is the most prevalent clone in South Africa, followed by ST152; ST101 was most prevalent in Durban, followed by Pretoria. Notably, all the *K. pneumoniae* strains from South Africa had multiple ARGs, with ST101 and ST152 having the most abundant and diverse repertoire of ARGs, including NDM, GES-5 and CTX-M-15 (Fig. 8C; Tables S5-S6).

**Figure 8.**
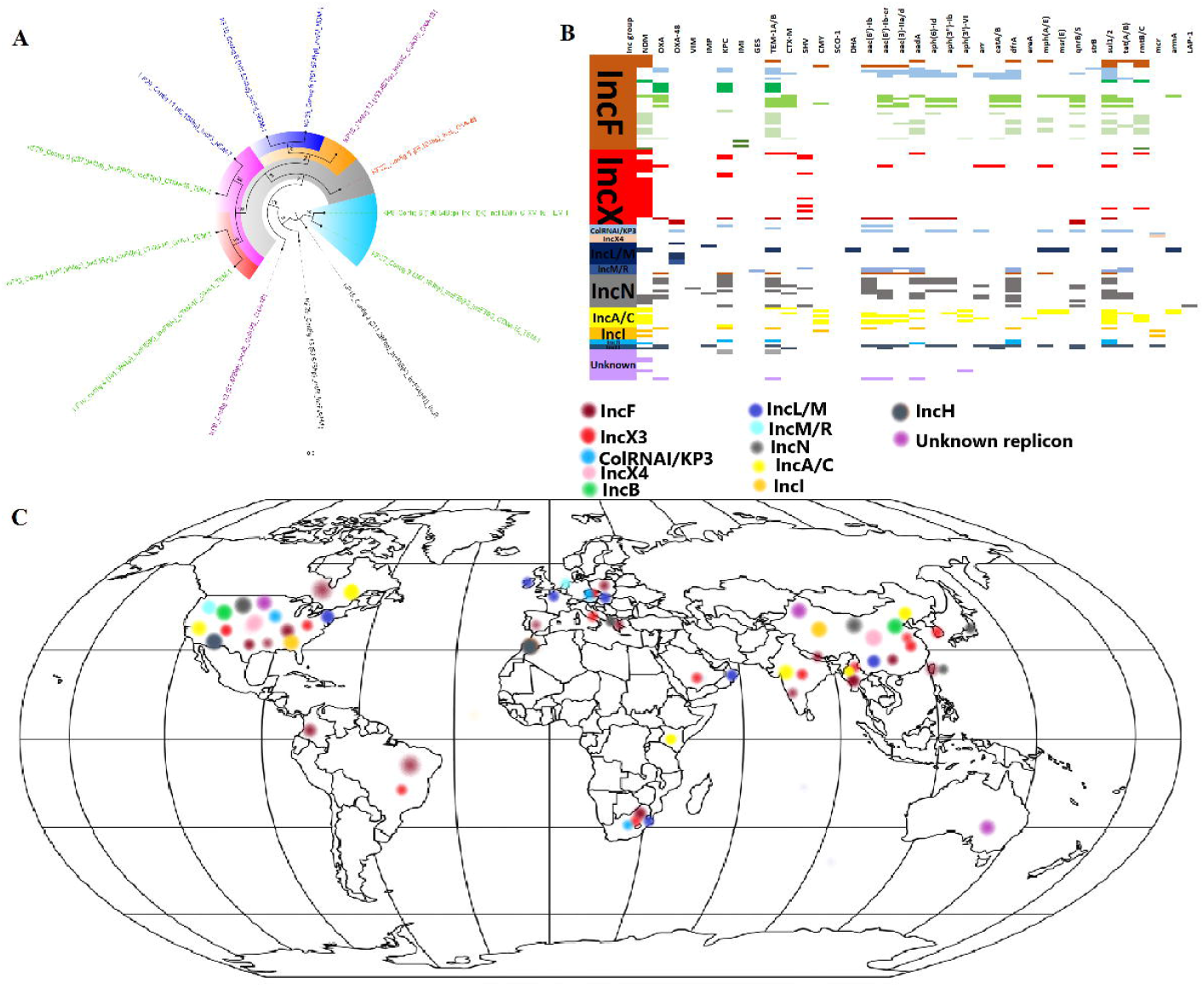
Phylogeography and resistome dynamics of *Klebsiella pneumoniae* isolates from South Africa. Each strain is expressed in specie name, strain, sequence type, date and country of isolation, and host. The phylogenetic relationship between the six isolates are shown in (**A**) whilst the phylogenetic relationship of the isolates with other isolates from South Africa are shown in (**B**): isolates from this study are labelled red, those from Pretoria are labelled in mauve/purple text, those from Durban are labelled in blue text, those from Pietermaritzburg are labelled in green text, and those from Ozwatini are labelled in turquoise; members of the same clade (labelled I to XIII) are highlighted with the same colour on the branches. The resistomes of the isolates are shown in (**C**) under the various clades, with members of the same clade having the same colour as the highlights in the phylogenetic tree in (**B**); blanks refer to absence of resistance genes and filled sections refers to presence of resistance genes. The phylogeography of the various clades are shown in (**D**); the isolates were mostly from Pretoria and Durban, with some being from Pietermaritzburg and Ozwatini.

Genomes of *K. pneumoniae* isolates from Africa were mainly obtained from Algeria, Cameroon, Egypt, Ghana, Guinea, Malawi, Nigeria, Niger, South Africa, Sudan, Tanzania, Togo, Tunisia, and Uganda. These clustered in 17 clades and were not restricted to a single country except clades II, VIII, X, XIII and XVI, showing the distribution of same clades across several African countries. ST101, which clustered into two clades, was the commonest clone in Africa (South Africa, Egypt, Tunisia, and Ghana) whilst ST152 was also common, but was mainly found in South Africa (Fig 9). Individual clones were mostly found on the same branches, albeit different clones were also found on within the same clade; particularly, ST716 and ST1552 as well as ST25, ST307 and ST152 clustered on the same branches despite their different clonalities. Multiple ARGs were present in the genomes of all these *K. pneumoniae* genomes from Africa; notwithstanding, genes encoding NDM, GES-5, and OXA-48 were only found in a few clades viz., IX, X, XII, XVII (Fig. 9).

**Figure 9.**
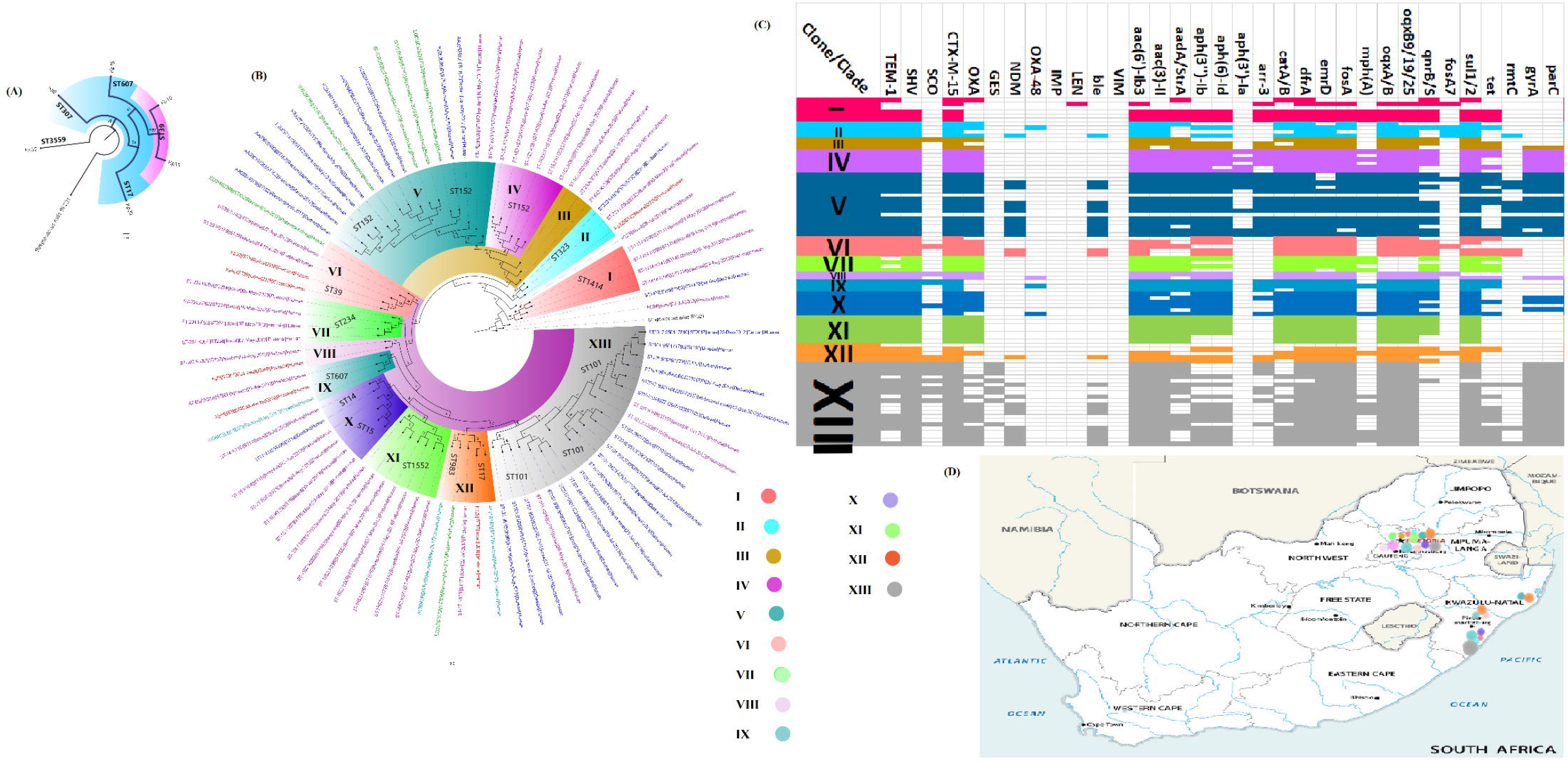
Phylogeography and resistome dynamics of *Klebsiella pneumoniae* isolates from Africa. Each strain is expressed in specie name, strain, sequence type, date and country of isolation, and host on the phylogenetic tree shown in (**A**) under different label colours representative of the country of origin: green (South Africa), red (Ghana), brown (Nigeria), purple (Algeria), blue (Egypt), turquoise (Tunisia), gold (Cameroon), and black (Sudan, Uganda, unknown country). Members of the same clade (labelled I to XVII) are highlighted with the same colour on the branches. The resistomes of the isolates are shown in (**B**) under the various clades, with members of the same clade having the same colour as the highlights in the phylogenetic tree in (**A**); blanks refer to absence of resistance genes and filled sections refers to presence of resistance genes. The phylogeography of the various clades are shown in (**C**) and most of the clades are concentrated in South Africa, Nigeria, Ghana, Tanzania, Malawi, Algeria, Tunisia, and Egypt.

Globally, *K. pneumoniae* strains producing carbapenemases clustered into 15 main clades, with intercountry transmission being identified in most clades. Italy had numerous ST101, ST2502 and ST512 clones suggestive of local outbreaks. Vietnam had substantial ST15 clones whilst South Africa had ST101 and ST152 clones, suggestive of local clonal transmissions. Worryingly, multiple resistance determinants were seen in all clades (Fig. 10; Table S6).

**Figure 10.**
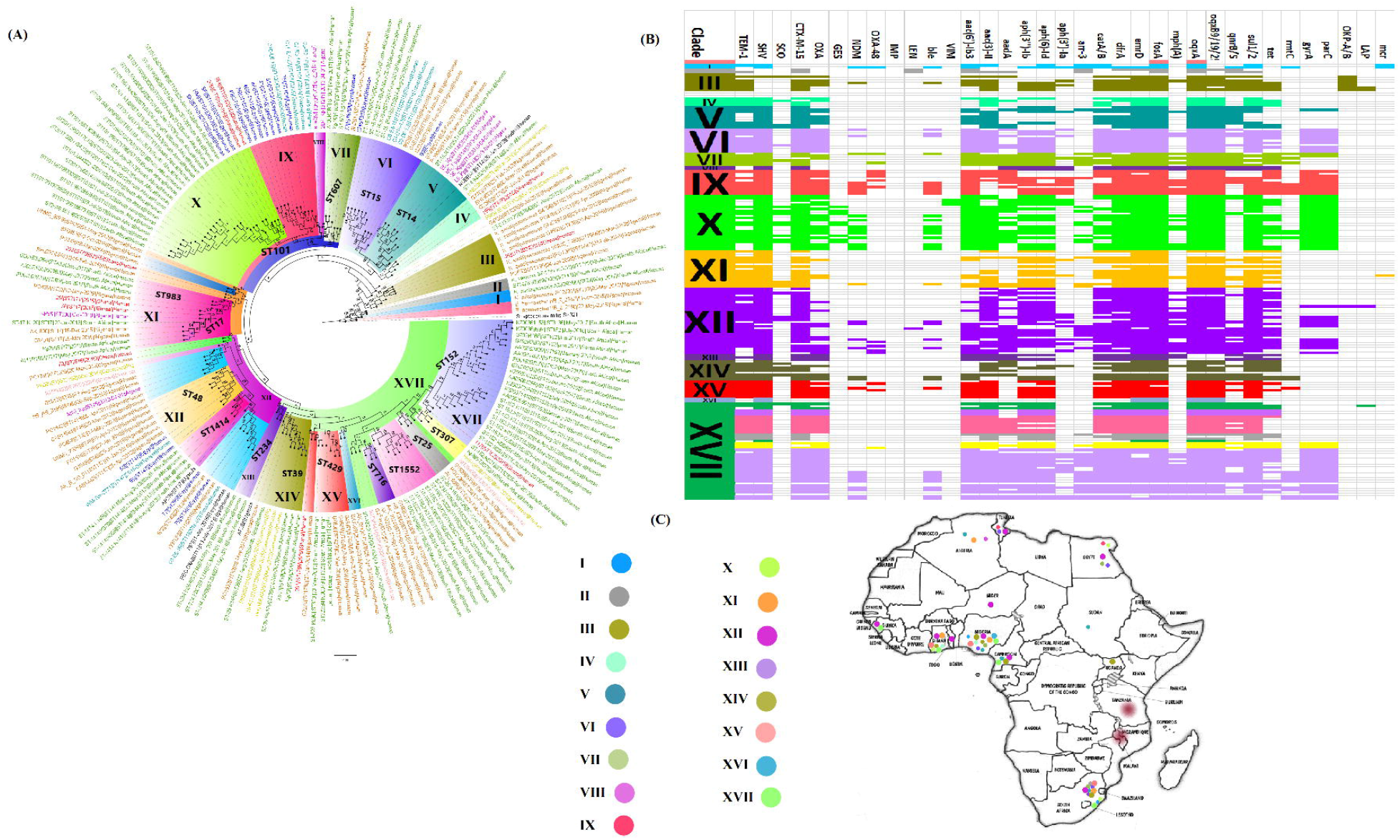
Global phylogeography and resistome dynamics of carbapenemase-producing *Klebsiella pneumoniae* isolates. Each strain is expressed in specie name, strain, sequence type, date and country of isolation, and host on the phylogenetic tree shown in (**A**) under different label colours representative of the country of origin: purple (Italy), green (South Africa), red (China), blue (United States), lemon green (Singapore), orange (U.A.E), pink (Sweden), turquoise (Thailand), gold (Vietnam), mauve (Malaysia), orange (UK, India or Spain). Members of the same clade (labelled I to XVI) are highlighted with the same colour on the branches. The resistomes of the isolates are shown in (**B**) under the various clades, with members of the same clade having the same colour as the highlights in the phylogenetic tree in (**A**); blanks refer to absence of resistance genes and filled sections refers to presence of resistance genes. The phylogeography of the various clades are shown in (**C**) and most of the clades are concentrated in the USA, Brazil, Europe, UAE, South Africa, and South-Eastern Asia.

## 3.4 Discussion

*K. pneumoniae* is becoming a common cause of fatal and untreatable nosocomial infections worldwide with perennial local outbreaks. Herein, we comprehensively characterised the genomic and epigenomic determinants mediating MDR, virulence and evolutionary epidemiology of this species. We show that *K. pneumoniae* strains circulating in South Africa, Africa and globally are bearers of multiple resistance and virulence determinants through clonal, multiclonal and plasmid-mediated dissemination. We also identified and characterised plasmids, transposons, integrons and ISs mobilising ARGs, virulence genes and RMS within and between *K. pneumoniae* strains, evinced by a global plasmid evolution and resistome analyses that showed the global distribution of similar plasmid types. More revealing is the RMS and methylation signatures in these strains, which play a crucial role in their virulence, transposition, transcription, replication, DNA repair and resistance regulation ^28,29,31^.

A striking but worrying feature of the isolates used in this study is their MDR phenomes, including resistance to last resort antibiotics such as colistin and carbapenems, and their widespread distribution in six major healthcare centres in Pretoria (Tshwane municipality) (Table S1). Indeed, the precarious nature of this situation is evinced by the recent outbreaks of carbapenem-resistant *K. pneumoniae* in hospitals in Pretoria and Johannesburg that took the lives of several infants ^32,33^. Priorly, other studies already reported of such fatal outbreaks in other public and private sector hospitals in other provinces in South Africa ^12,22,34^, making this a national more than a local problem necessitating urgent public health interventions to stem this menace in the bud ^35^.

Herein, the CRKP isolates mainly harboured *bla*_OXA-48_ and *bla*_NDM-1,_ which differs from a South African report by the National institute for communicable diseases (NICD) in 2015 in which a high prevalence of *bla*_NDM-1_-producing *K. pneumoniae* in the Gauteng and Kwazulu-Natal provinces and a few reports of *bla*_OXA-48_-producing *K. pneumoniae* in Gauteng and Eastern Cape provinces were described ^36^. Similar results were also reported in 2016 by Perovic and colleagues, where a high prevalence of *bla*_NDM-1_ was observed in the Gauteng province^37^. However, a recent study (2019) reported on an exponential increase in *bla*_OXA-48_-like producing *K. pneumoniae* strains^38^, which agrees with this study and suggests a change in carbapenem resistance determinants in *K. pneumoniae* in Gauteng. Evidently, NDM-positive strains had higher carbapenem resistance than OXA-48/181 strains (Table S1), which further agrees with previous reports that OXA-48-like enzymes are less hydrolytic than NDM ones ^19^. Although isolates with both carbapenemases were also highly resistant, they did not have a higher MIC than those with only NDM (Table S1), which could be due to the lower hydrolytic ability of OXA-48-like enzymes. The absence of *mcr* in the isolates suggest chromosomal mutations-mediated colistin resistance.

The REP-PCR, albeit of a poorer resolution than whole-genome-based typing, revealed a major strain that was reported in majority of the *K. pneumoniae* isolates in this study, mainly carrying the *bla*_OXA-48_ gene. One of the isolates in this group was sequenced and we identified *bla*_OXA-181_ and ST307, which has been associated with hospital outbreaks and multiple ARGs such as *bla*_CTX-M-15_, *bla*_NDM-1_, *bla*_KPC_, *bla*_OXA-48_, and *mcr*-1 genes ^39–43^. In South Africa, OXA-181-producing *K. pneumoniae* ST307 isolates were reported previously in the private sector hospitals in six provinces, including Gauteng ^38^. In this study, the *K. pneumoniae* ST307 isolates were collected from government sector hospitals in the Tshwane area.

Another strain detected by REP-PCR was found to contain *bla*_OXA-181_ and belonged to ST607, a rare clone reported in China and in a neonatal ICU in France ^44^. The *bla*_OXA-48_-producing *K. pneumoniae* strain, ST3559, has been recently reported as a novel clone among CRKP isolates collected from hospital wastewater, influent wastewater, river water, and riverbed sediments in South Africa^45^. These *K. pneumoniae* isolates shared the same molecular and MDR characteristics with isolates from this present study (Data S2), suggesting that the same strain is now circulating in hospitals in the Tshwane area. *Bla*_NDM-7,_ the first to be found in *K. pneumoniae* ST17 in S. Africa, was reported in *K. pneumoniae* ST147 and ST273 involved in nosocomial cases in Canada, Gabon, Philippines, US and India ^46–50^.

Moreover, the phylogeography and resistome analyses of *K. pneumoniae* genomes from South Africa, Africa, and global carbapenemase-producing *K. pneumoniae* (CPKP) strains shows the rich resistance repertoire of this species. Notably, similar *K. pneumoniae* clones are circulating within mainly Gauteng (Pretoria) and Kwazulu-Natal (Durban, Pietermaritzburg and Ozwatini) provinces in South Africa (Fig. 8), suggesting the exchange of carriers of these strains between these distant provinces. Indeed, other non-human factors could be involved in this interprovincial transmission and the earlier these carriers are identified the better. Furthermore, the presence of same clades/clones, particularly ST101, ST14, ST15 and ST17 between South Africa, West Africa, and North Africa, bearing multiple clinically important ARGS is revealing. Globally, CPKP is mainly concentrated in the US, South Africa, Europe and South-East Asia, with relatively fewer reports from the Arabian peninsula and Brazil, a further evidence that AMR transcends borders and requires epidemiological investigations to detect and break the transmission chain.

Worryingly, almost all these *K. pneumoniae* and CPKP isolates co-harboured numerous other ARGs, which makes them potentially multi-, extensively and pan-drug resistant, an observation already made in this (Fig. 1 &Table S1) and other studies ^12,22^. Hence, the administration of non-carbapenem antibiotics has a higher chance for selecting CRKP (carbapenem-resistant *K. pneumoniae*) and CPKP strains due to their rich resistome repertoire. It is thus not surprising that most *K. pneumoniae* infections are less responsive to treatment and have easily spread worldwide ^51^. Coupled with this is the rich virulence genes and capsule repertoire of *K. pneumoniae* that enable them to escape host immune forces ^52^. As shown previously, the virulome was not affected by the isolation source ^12^. Worryingly, *traT* genes, encoding an outer membrane protein that resists complement proteins, were found on plasmids in all the isolates (Table S3). Gordon et al. (2020) recently found O1 and O2 capsule serotypes, also found in this study, to be most associated with MDR; they also found the K2 serotype, present in KP10 and KP33, to be most common among global *K. pneumoniae* isolates ^52^.

Almost 79 capsule polysaccharide types based on the K-loci of *K. pneumoniae* have been described, and of these, only K1 and K2 serotypes are associated with hypervirulent strains whilst the others are associated with classical strains of *K. pneumoniae*^53,54^. Our results showed that two of the sequenced isolates (Kp10 and Kp33), which are highly resistant to carbapenems and harboured the *bla*_NDM-1_ gene, were KL2 serotypes. This might indicate that these isolates are K2-hypervirulent *K. pneumoniae* (K2-hvKP) strains. K2-hvKP strains were not given attention until the report of multidrug-resistant K2-hvKP strain harbouring the *bla*_KPC-2_ and *bla*_IMP-4_ in China^55^. Following this report, multiple studies, including this study, have reported on carbapenemase production in highly virulent stains of the K2 serotype^56,57^. Another study in China reported on an ST11 *bla*_KPC-2_-producing strain (CR-HvKP1), which is closely related to strains in this study (Supplemental data 2); this CR-HvKP1 harboured a virulence plasmid (pLVPK-like) and showed a highly resistant profile ^58^.

The plasmid evolutionary epidemiology of the plasmids identified in this study, as well as their resistomes, provide deeper insights into the role of plasmids in the dissemination ARGs in Enterobacterales. Notably, the close evolutionary alignment/distance of plasmids bearing same or different ARGs, but belonging to different incompatibilities in the same isolate (Fig. 7a), depicts the genetic exchanges (recombinations and rearrangements) that occur between plasmids and between plasmids and chromosomes during replication ^59,60^. Evidently, the very close sequence and resistome similarity between this study’s plasmids and those obtained from different and same species in other studies worldwide portrays the global dissemination of IncF, IncX, A/C, IncN, and IncI plasmids and their role in ARGs dissemination among bacteria. Furthermore, it shows that not all plasmid types harbour rich resistomes as IncX was mainly limited to only three ARGs. The relatively lower diversity and abundance of ARGs on these plasmids (Fig. 7) than the resistomes observed in the *K. pneumoniae* strains (Fig. 8-10) is because the *K. pneumoniae* strains contain multiple plasmids alongside chromosomes, which bear additional ARGs.

Previous studies identified *bla*_NDM-1_ on IncF, IncL/M, IncN, A/C, and IncX plasmids ^19^. Herein, *bla*_NDM-1_-producing *K. pneumoniae* were mostly associated with IncF (FII, F, FIB, FIC), followed by IncL and A/C plasmid replicons (Table S1), which agree with reports in Nepal, Taiwan, Oman, Myanmar, Canada and South Africa^22,61–63^. However, in China, Gabon, India, and Japan, NDM-1/7-variants in *K. pneumoniae* were on IncX_3/4_ plasmids ^57,64^. The *bla*_OXA-181_-producing isolates were on ColKP3, IncX_3_, and IncF plasmids, similar to previous studies from Czech Republic, Denmark, Sao Tome & Principle, and South Africa^38,65–67^. In South Africa, IncX3 was found in *K. pneumoniae* collected during a hospital outbreak^38^. An earlier study reported the significant role that IncL/M plasmids play in the dissemination of *bla*_OXA-48_ genes in *K. pneumoniae* strains worldwide^67–72^. Our findings also prove that *bla*_OXA-48_ gene is usually located on conjugative IncL/M plasmids. The IncHI1B plasmid replicon was responsible for the carriage of *bla*_NDM-1_ in strain KP33_1^73^, different from *bla*_NDM-1_-producers in this study. A strain reported in the United States (CN1) which harboured an IncFII/FIB multi-replicon showed similar molecular characteristics with bla_NDM-1_-producers in this study, but it belonged to ST392. Thus, ARGs dissemination may occur in diverse clones via different plasmid replicon types.

There were discrepancies in plasmid numbers and sizes between the gel-based plasmid characterisation and sequencing analyses, which is expected. This could be due to the break-up of the plasmids during the extraction process, leading to smaller sizes and higher plasmid numbers. The PCR-based plasmid typing scheme used in this study was unable to detect IncX3 plasmids, which were revealed with whole-genome sequencing. This is a limitation in areas where this typing scheme is solely used as it needs to be modified with new primers targeting all subtypes of the replicon groups.

No single plasmid harboured both ESBL and carbapenemase genes together and the plasmids harbouring the ESBL genes were of larger sizes with richer resistomes that clustered together on resistance islands, further supporting the fewer resistomes observed on carbapenemase plasmids worldwide (Fig. 7). The genetic environment of all the ARGs and mercuric resistance operons on these self-conjugative plasmids were surrounded by MGEs, which undoubtedly will facilitate their horizontal transmission. Notably, the genetic support of these ARGs, particularly of the ESBLs, were similar to those already reported in other studies: *bla*_CTX-M-15_ was always next to ISEc9, *bla*_TEM_ was always next to an integrase/recombinase in close synteny to *bla*_CTX-M-15,_ and *bla*_OXA_ was always surrounded by *cat* and *aac(3’)-II* genes. As well, *bla*_NDM-1/7,_ *bla*_OXA-181_, and *bla*_OXA-48_ were also found within several MGEs (Fig. 2-6), supporting their horizontal transposition.

Prophages (bacteriophages), which are involved in transduction of genetic material horizontally, were abundant on plasmids and chromosomes, with several partial prophage DNA, suggesting the occurrence of a transduction activity (Fig. S7-S12). Indeed, the presence of same prophages in different isolates and on both plasmids and chromosomes depict their horizontal movement and importance in influencing MDR and genomic plasticity or evolution. Evidently, their presence on these self-transmissible plasmids suggests that they can be also shared during conjugation. Yet, the presence of prophage DNA and RMS on plasmids and chromosomes is intriguing as RMS identify and destroy foreign DNA, including prophages ^28,29,74^. However, prophages found on plasmids with the same methylation signature as the host bacterium may escape destruction by REs and CRISPR-Cas complexes. Notwithstanding, the interplay between phages and RMS on the same plasmids and chromosomes will need further investigation to appreciate their co-existence and co-evolution.

The isolates were remarkably endowed with rich RMS comprising types I, II and III RMS, with type II RMS being more abundant as reported ^29,31,74^. Huang et al. (2020) recently showed that type I RMS were scarce in CPKP, obviously due to the destruction of carbapenemase-bearing plasmids by the type I RMS; the exception was in isolates that had the type I RMS on plasmids to protect them from REs ^75^. It is thus not surprising that plasmids harbouring ESBLs and carbapenemases in this study’s isolates also had RMS that shared the same Dam and Dcm motifs with those on the chromosomes and CCWGG motifs of type II Dcms were virtually absent on chromosomes but ubiquitous on plasmids (Table S4). Indeed, these RMS were found within MGEs, as already shown ^74^, which evidently facilitates their movement between plasmids, chromosomes and bacteria. Specifically, the multiple copies of *M*.*Sfl2ORFAP* on both plasmids and chromosomes in several isolates suggest a transposition event ^74^. Due to the destruction of plasmids or DNA without the same methylation signatures by REs, the presence of the same RMS on plasmids and chromosomes facilitate their safe entry into host bacteria, enhancing dissemination of virulence and resistance plasmids between different species.

The ubiquitous presence of G**<u>A</u>**TC motifs on only chromosomes and their absence on plasmids evince the conservation of these motifs and their associated Dams in prokaryotes. Notably, the preponderance of type II Dcms with C**<u>C</u>**WGG motifs on plasmids suggest that their relatively limited presence on chromosomes is due to transposition from plasmids to chromosomes. Recently, environmental bacteria were found to be abundant in Dams with G**<u>A</u>**TC and **<u>A</u>**TGNNNNNNGCT motifs ^28^, which where were mostly found on this study’s isolates’ chromosomes. G**<u>A</u>**TC and C**<u>C</u>**AGG (C**<u>C</u>**WGG or C**<u>C</u>**NGG in this study) motifs have been also identified in *E. coli* ^59,74^, showing that plasmids with these RMS and methylation signatures can be shared between environmental and clinical prokaryotes. Therefore, the important role of RMS in horizontal resistomes and virulome transmission and regulation cannot be gainsaid. Further, the regulatory roles of RMS in bacterial virulence was recently confirmed in hvKP strains in Taiwan; *dam*^*-*^ mutants were found to be less pathogenic in mice and serum than their wild-type *dam*^*+*^ strains. Hence, the presence of these rich RMS in the isolates will facilitate their virulence in human and animal hosts. More concerning is the ability of these RMS to defeat bacteriophage therapy. Indeed, a recent study showed how *K. pneumoniae* quickly developed resistance to bacteriophages during treatment ^76^. Subsequently, detailed investigations should be undertaken to find ways to protect bacteriophage therapy from destruction or resistance by these RMS systems.

## 3.5 Conclusion

This study has shown the dissemination of *bla*_OXA-48_-like and *bla*_NDM_ carbapenemases in MDR *K. pneumoniae* isolates in hospitals in Gauteng, South Africa, as well as MDR *K. pneumoniae* in Africa and globally through self-transmissible IncF, A/C, IncX3 and IncL/M plasmids. The global evolutionary epidemiology of *K. pneumoniae* and associated plasmids show the dissemination and preponderance of MDR, XDR and PDR CRKP globally, including the possibility of transmitting these plasmids to other bacterial hosts, threatening infectious disease management. Notably, the important role of RMS in regulating and facilitating the transcription, transposition, and dissemination of resistance and virulence plasmids is revealing and requires further investigation as it also threatens both antimicrobial and bacteriophage chemotherapy. It is essential that rigorous infection prevention and control are instituted to avoid the selection and dissemination of plasmids harbouring RMS, virulence and MDR genes in prokaryotes.

This study showed *K. pneumoniae* isolates to have resistance profiles to most antibiotics, including colistin. Among all the tested antibiotics, only a few (amikacin, fosfomycin, and tigecycline) were still active against these isolates. This raises more concern about treatment options for CRKP, because colistin is one of the last-resorts for infections caused by these pathogens. Due to this reason, and reports of poor outcome of colistin monotherapy ^13^, clinicians are left with limited or no treatment options.

## Materials and Methods

### Bacterial strains and antimicrobial susceptibility testing

A total of 60 non-repetitive *K. pneumoniae* isolates were collected from a referral laboratory (National Health Laboratory Service/NHLS) in Pretoria. The VITEK 2® automated system (BioMerieux-Vitek, Marcy-l’Étoile, France) was used for species identification and antimicrobial susceptibility testing; only those resistant to at least one carbapenem (ertapenem, meropenem, imipenem, doripenem) were included in further analyses. The *K. pneumoniae* isolates were received on blood agar plates (NHLS, SA) and incubated at 37°C for 24 hours. Following incubation, confirmation of the Minimal Inhibitory Concentrations (MIC) of the isolates were determined using the MicroScan automated system (Beckman Coulter, California, United States). The results were interpreted according to the Clinical and Laboratory Standards Institute (CLSI) breakpoints.

### DNA extraction of carbapenem-resistant K. pneumoniae (CRKP) isolates

Total genomic DNA was extracted from all carbapenem-resistant isolates. The DNA was extracted from an overnight Brain Heart Infusion (BHI) broth using the boiling method. The cells were heated at 95°C using a digital dry bath (Labnet International, New York, United States) for 15 minutes and transferred to an ultrasonic bath (Lasec Ltd, Midrand, South Africa) for another 15 minutes. The resulting supernatant was stored at −20°C freezer until needed for further analysis and it was used as a template for the PCR assays.

### Detection of carbapenemase genes using PCR assays

PCR was used to screen for the presence of six carbapenemase genes viz., *bla*_IMP_, *bla*_KPC_, *bla*_VIM_, *bla*_OXA-48_, *bla*_NDM_, and *bla*_GES_. Specifically, multiplex PCR was used for determining the presence of *bla*_VIM_, *bla*_OXA-48_, and *bla*_NDM_ while simplex PCR was used for *bla*_IMP_, *bla*_KPC_, and *bla*_GES_ screening. The oligonucleotide primers were synthesized by Inqaba Biotechnical Industries (Pretoria, SA) (Supplemental data 1). For the PCR reaction, 1 µl of template DNA was added to 12.5 µl of MyTaq™ HS mix (Bioline, London, United Kingdom) while 0.4 µM of each primer and nuclease-free water (Qiagen, Hilden, Germany) were added to make up the volume to 25 µl in each PCR tube. The multiplex PCR conditions were as follows: 95°C for 5 min, followed by 25 cycles of 95°C for 30 sec, 57°C for 45 sec, and 72°C for 30 sec, and a final extension step at 72°C for 7 min. The PCR amplicon were analysed using 1.5% Seakem agarose gel (Whitehead Scientific (Pty) Ltd, Cape Town, SA) with 5 µl ethidium bromide and visualised under Ultraviolet light using the Gel Doc™ EZ Gel (BioRad Laboratories, California, US) bioimaging system. A 100bp ready-to-use DNA ladder (Celtic Molecular Diagnostics, Cape Town, SA) was used to determine the size of the expected genes. All PCR amplicons were run alongside a positive and negative control.

### Genotyping using Repetitive Extragenic Palindromic (REP) PCR assay

Total genomic DNA from all carbapenem-resistant *K. pneumoniae* isolates were used as template in the REP-PCR assay. The primer pair sequences REP 1 (5’-IIIGCGCCGICATCAGGC-3’) and REP 2 (5’-ACGTCTTATCAGGCCTAC-3’) and PCR conditions described previously were used in this assay ^77^. For the PCR reaction, 1 µl of template DNA was added to 12.5 µl of MyTaq™ HS mix (Bioline, London, United Kingdom) while 0.4 µM of each primer and nuclease free water (Qiagen, Hilden, Germany) was added to make up the volume to 25 µl in each PCR tube. The PCR conditions were as follows: an initial denaturation of 94°C for 3 min, followed by 30 cycles of 94°C for 45 sec, 45.8°C for 1 min, and 72°C for 8 min and a final extension step of 72°C for 16 min. The amplified DNA amplicons (10 µl) were separated by electrophoresis using 1.5% SeaKem agarose gel (Whitehead Scientific (Pty) Ltd, Cape Town, South Africa) with 5 µl ethidium bromide. The gels were run for 3 hour 20 minutes at 80 volts. The DNA amplicons bands were visualised under Ultraviolet light using the Gel Doc™ EZ Gel (BioRad Laboratories, California, United States) bioimaging system and banding patterns were compared to a 1 kb plus ready-to-use DNA ladder (Thermo Fisher Scientific, Massachusetts, United States). Analysis of REP-PCR fingerprints was performed using the GelCompare II software (Applied Maths, Belgium, Europe). Relatedness was determined by means of the Dice coefficient and unweighted pair group method with arithmetic mean (UPGMA). In this study a similarity coefficient of 75% was used to determine different strains of CRKP, i.e. isolates that showed a similarity of 75% were considered part of the same strain.

### Plasmid characterisation using the PBRT scheme

Plasmid DNA extracted using the plasmid midi kit (Qiagen, Hilden, Germany) was used as template in characterising plasmids using the PCR-based inc/rep typing scheme. These plasmids were typed by targeting 19 replicon groups reported in *Enterobacteriaceae* and one replicon group targeting a virulence plasmid in *K. pneumoniae*. This method was carried out as previously described with few modifications ^78,79^. Modifications were made in multiplex 5, where A/C and IncT were detected in a multiplex and IncFII plasmids were detected in a simplex PCR assay instead of multiplex. The IncFII_k_ virulence plasmids in *K. pneumoniae* were also detected. The PCR assays were performed using a SimpliAmp Thermal cycler mini (Thermo Fisher Scientific, Massachusetts, US) and the PCR conditions used were described previously ^78,79^. Briefly: initial denaturation of 94°C for 5 min, followed by 30 cycles of 94°C for 1 min, 60°C for 30 sec, and 72°C for 1 min, and a final extension of 72°C for 5 min. For IncF, IncFII, and IncFII_k_ plasmids, same conditions were used except that an annealing temperature of 54°C for 30 sec was used instead. Supplemental data 1 shows all the primer sequences that were used for these assays.

### Resistance plasmid transferability/mobility

Transferability of resistance plasmids was determined using conjugation experiments. The experiments were performed on 26 isolates showing reduced susceptibility to meropenem using a broth mating method. The meropenem-resistant isolates were used as plasmid donors and the *E. coli* J53-A^r^ (sodium-azide resistant) strain served as a recipient strain. For broth mating, 3-hour growth cultures of donor and recipient strains grown in Luria Bertani (LB) broth (VWR international, Pennsylvania, US) were mixed with each other at a ratio of 1:4 (donor to recipient) and incubated at 37°C for 3 hours. Grown cells (200 µl) of the mixtures were spread onto Mueller-Hinton agar (Sigma-Aldrich (Pty) Ltd, Missouri, US) containing 0.5 µg/ml meropenem (Sigma-Aldrich (Pty) Ltd, Missouri, US) and 100 µg/ml sodium azide (VWR international, Pennsylvania, US) to select only for plasmid-encoded carbapenem resistance and then incubated at 37°C for 24 or 48 hours. PCR assay was used to confirm the carbapenemase gene (*bla*_NDM-1_ and/or *bla*_OXA-48_) carriage by transconjugants.

### Whole-genome sequencing of K. pneumoniae isolates

A total of 6 representative isolates, based on their carbapenemase gene, REP pattern, plasmid number and type, were selected for WGS. Genomic DNA was extracted from the *K. pneumoniae* isolates Kp8, Kp10, Kp15, Kp29, Kp32, and Kp33 using a Zymo Research Fungal/Bacterial kit (Inqaba biotec, Pretoria, South Africa) according to the manufacturer’s instructions. Genomic DNA was sent for sequencing at Inqaba Biotec (Pretoria, South Africa) on the PacBio RSII sequencer (Pacific Biosciences, Menlo Park, CA, United States) at an average coverage of 90x.

### Genomic analyses and annotation

PacBio’s SMRT® Link v8.0 software suite was used for trimming the raw reads, assembling with HGAP, and determining methylation modifications and motifs (using MotifMaker: https://github.com/PacificBiosciences/MotifMaker) in the genomic sequences. DNA methylases (MTases), restriction endonucleases (REases) and their motifs were searched from the Restriction Enzyme Database (REBASE)^80^. Complete genomic annotations were done with NCBIs PGAP^81^. Clonal sequence types, resistance and virulence genes, plasmid typing, integrons, transposons, insertion sequences and prophages were determined using online databases including MLST2.0 ^82^, ResFinder ^83^, BacWGSTdb ^84^, Plasmidfinder^85^, INTEGRALL (http://integrall.bio.ua.pt/), ISFinder^86^ and PHASTER^87^, respectively. *K. pneumoniae* capsule polysaccharide-based serotyping (K-type) was performed using the Kaptive Web database^88^.

### Chromosomal colistin and fluoroquinolones resistance mutations

Mutations conferring resistance to colistin and fluoroquinolones were determined from the assembled genomes using BLASTn. Briefly, *mgrB, crrB, kpnEF, phoPQ, pmrAB, gyrA, gyrB parC* and *pare* genes in reference K. pneumoniae ATCC 13883 (PRJNA244567) were aligned with this study’s genomes using BLASTn. The mutations in the study’s isolates’ genomes were manually curated.

### Phylogenomic, phylogeography and resistome analyses

Genome sequences of CRKP strains from South Africa (n=88), Africa (n=380) and globally (n=343) were downloaded from the PATRIC website (https://www.patricbrc.org/); carbapenemase-producing *K. pneumoniae* genomes (n=190) were further culled from the global CRKP genomes for the global carbapenemase-producing *K. pneumoniae* phylogenomics. These genomes, alongside those from this study, were used for the whole-genome phylogenomics. Four phylogenomic trees viz., one for this study’s genomes, one for South African genomes, one for African genomes and one for global genomes, were drawn using RAxML’s maximum-likelihood based phylogenetic inference. A bootstrap reassessment of 1000x was used and the trees were annotated using Figtree (http://tree.bio.ed.ac.uk/software/figtree/). Isolates with strong bootstrap support values (>50) were clustered into a clade and highlighted with the same colour. The resistomes of these genomes were downloaded from NCBI’s Pathogen/Isolate Browser database (https://www.ncbi.nlm.nih.gov/pathogens/isolates#/search/) and the clades’ phylogeographies were manually mapped.

### Genomic plasmid typing, evolution, and resistome analyses

Plasmid genomes (n=26) from this study were aligned with MUSCLE. The aligned files were used to draw a phylogenetic tree with PhyML (http://www.phylogeny.fr/index.cgi), using a bootstrap sampling of 100x. The newick tree file was annotated with Figtree. The genomes of the 26 plasmids were aligned with those of closely related plasmids using progressive Mauve ^89^. The carbapenemase-bearing plasmids of this study were parsed through the BacWGSTdb database/server to determine other closely related plasmids, their source, incompatibility, and geographical locations ^84^. Carbapenemase-bearing plasmid genomes and meta-data were downloaded from NCBI and PATRIC (https://www.patricbrc.org/). Their plasmid replicons and resistomes were determined using PlasmidFinder^85^ and ResFinder^83^. These were arranged according to their replicons/incompatibility groups and mapped to show their geographical distribution.

### Data availability

All data used in this study are found in the supplemental data and Tables (S1-S5). The genomes of the isolates used in this study have been deposited in DDBJ/ ENA/GenBank under BioProject number **PRJNA565241** (https://www.ncbi.nlm.nih.gov/bioproject/PRJNA565241) and accession numbers VXIW00000000 (KP33), VXIX00000000 (KP32), VXIY00000000 (KP29), VXIZ00000000 (KP15), VXJA00000000 (KP10) and VXJB00000000 (KP8); the versions used in this study are versions VXIW01000000, VXIX01000000, VXIY01000000, VXIZ01000000, VXJA01000000, and VXJB01000000.

The accession numbers of complete circular plasmids are VXIX01000005 (pKP32.5_OXA-48), VXIY01000008 (pKP29.9_CTXM-15), VXIY01000013 (pK29.13_MBELLE), whilst those of partial plasmids are VXJB01000006 (pKP8.6_CTX-M-15), VXJB01000012 (pKP8.12_OXA181), VXJA01000008 (pKP10.8_NDM-1), VXJA01000004 (pKP10.4_OSEI), VXIZ01000004 (pKP15.4_KATLEGO), VXIZ01000012 (pKP15.12_OXA-181), VXIY01000011 (pKP29.11_NDM-7), VXIX01000003 (pKP32.3_CTXM-15), VXIW01000008 (pKP33.8_NDM-1), VXIW01000004 (pKP10.4_OSEI).

All kinetic data (methylation) files have been deposited in GEO under accession number **GSE138949** (https://www.ncbi.nlm.nih.gov/geo/query/acc.cgi?acc=GSE138949) [GSM4125137 (Kp8); GSM4125138 (Kp10); GSM4125139 (Kp15); GSM4125140 (Kp29); GSM4125141 (Kp32); GSM4588290 (Kp33).

### Research ethics

Ethical approval was provided by Faculty of Health Sciences Research Ethics Committee (209/2018). All protocols and consent forms were executed according to the agreed ethical approval terms and conditions. All clinical samples were obtained from a reference laboratory and not directly from patients, who agreed to our using their specimens for this research. The guidelines stated by the Declaration of Helsinki for involving human participants were followed in the study

## Data Availability

All data used in this article are included as supplemental files and links to genomic data have been provided

## Funding

Funding for this study was provided by the NHLS, NRF (National Research Foundation) and the University of Pretoria.

## Acknowledgements

We are grateful to Sebastien Santini - CNRS/AMU IGS UMR7256 for curating the phylogeny (http://www.phylogeny.fr/index.cgi) database.

## Author contributions

**KK** undertook the laboratory work, data curation, initial descriptive statistics and initial draft of manuscript for her MSc work; **NMM** supervised the study and provided funding; **JOS** conceived, designed and supervised the study, undertook bio-informatics analyses and descriptive statistics, data curation, image designs, and the complete write-up, review and formatting of the manuscript.

---

**Figure S1.**
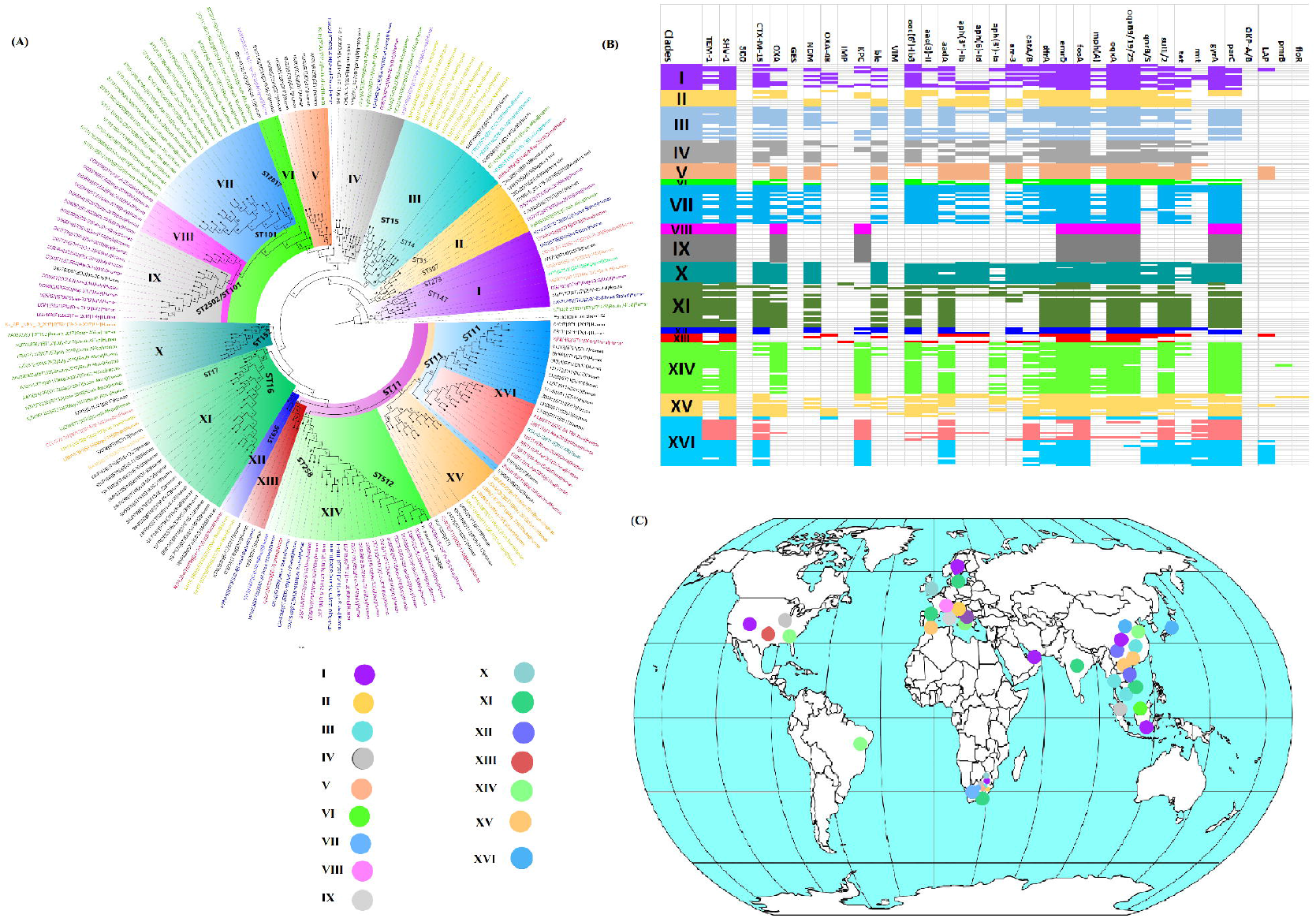
Graphical annotation and comparative alignment of pKP8.6_CTX-M-15. **(1.1)** The resistance genes (light green-coloured arrows), replicase genes (blue-coloured arrows), methyltransferases/restriction endonucleases (rose-coloured arrows), transposons (orange-coloured arrows), insertion sequences (orange-coloured arrows), integrons (red-coloured arrows), resolvases (red-coloured arrows), and recombinases/integrases (red-coloured arrows) on the plasmid is shown with their orientation (direction of arrow), synteny and immediate environment. Other genes with unknown functions are hidden to make the image less cluttered. A circular version of the plasmid is shown in (**A**). A linear version of the plasmid and its alignment with other similar plasmids (linear arrows with yellow highlighted background) are shown in (**B**); regions of alignment are shown as red-filled portions whilst non-aligned areas are shown as empty arrows. Enlarged section of the plasmid focusing on the resistance genes (genomic resistance island) are shown in (**C**), (**D**) and (**E**). This plasmid (VXJB01000006) contains CTX-M-15. (**1.2**) Mauve alignment of pKP8.6_CTX-M-15 (**A**) and pKP8.12_OXA181 (**B**) with other plasmids shows rearrangement of blocks (sections) of the closely related plasmids. Sections of the aligned plasmid genomes that aligns perfectly have the same colour.

**Figure S2.**
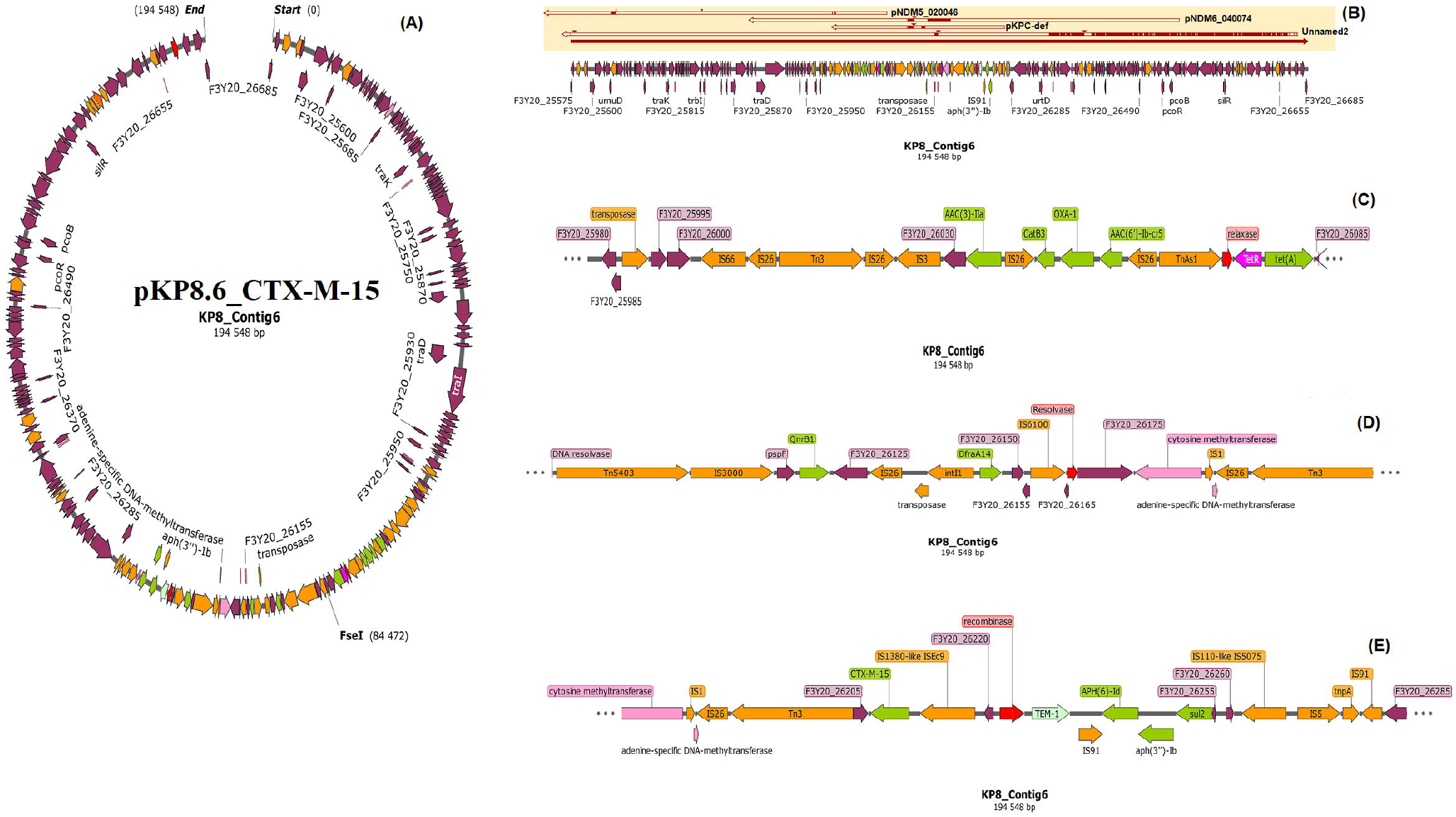
Graphical annotation and comparative alignment of pKP10.4_OSEI. **(2.1)** The resistance genes (light green-coloured arrows), replicase genes (blue-coloured arrows), methyltransferases/restriction endonucleases (rose-coloured arrows), transposons (orange-coloured arrows), insertion sequences (orange-coloured arrows), integrons (red-coloured arrows), resolvases (red-coloured arrows), and recombinases/integrases (red-coloured arrows) on the plasmid is shown with their orientation (direction of arrow), synteny and immediate environment. Other genes with unknown functions are hidden to make the image less cluttered. A circular version of the plasmid is shown in (**A**). A linear version of the plasmid and its alignment with other similar plasmids (linear arrows with yellow highlighted background) are shown in (**B**); regions of alignment are shown as red-filled portions whilst non-aligned areas are shown as empty arrows. Enlarged section of the plasmid focusing on the resistance genes (genomic resistance island) are shown in (**C**) and (**D**). This plasmid (VXJA01000004) contains TEM-1. (**2.2**) Mauve alignment of pKP10.8_NDM-1 (**A**) and pKP10.4_OSEI (**B**) with other plasmids shows rearrangement of blocks (sections) of the closely related plasmids. Sections of the aligned plasmid genomes that aligns perfectly have the same colour.

**Figure S3.**
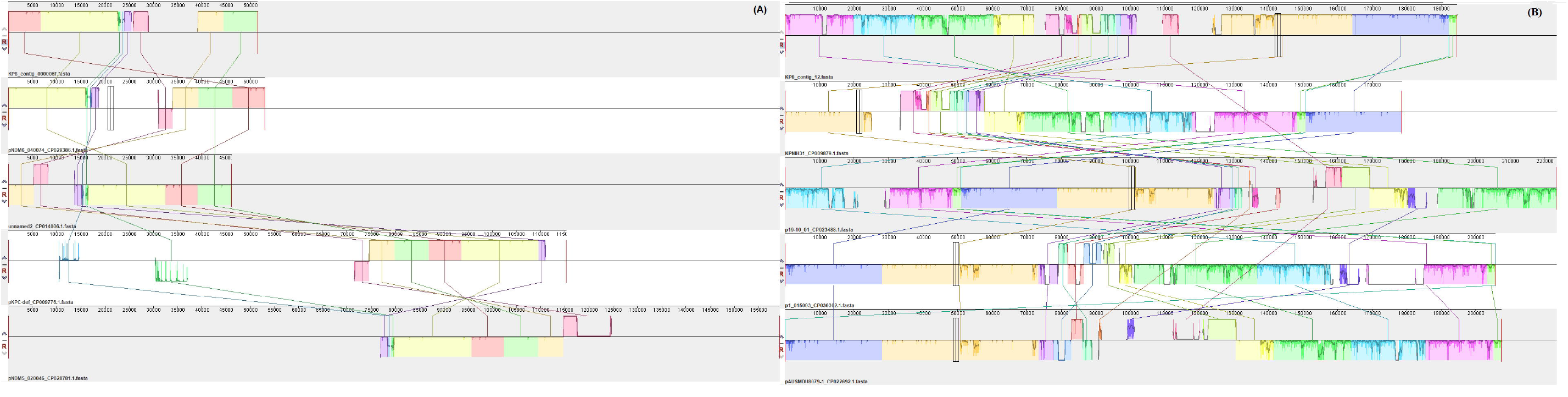
Graphical annotation and comparative alignment of pKP15.4_KATLEGO. **(3.1)** The resistance genes (light green-coloured arrows), mercury resistance and tolA genes (deep green-coloured arrows), methyltransferases/restriction endonucleases (rose-coloured arrows), transposons (orange-coloured arrows), insertion sequences (orange-coloured arrows), integrons (red-coloured arrows), resolvases (red-coloured arrows), and recombinases/integrases (red-coloured arrows) on the plasmid is shown with their orientation (direction of arrow), synteny and immediate environment. Other genes with unknown functions are hidden to make the image less cluttered. A circular version of the plasmid is shown in (**A**). A linear version of the plasmid and its alignment with other similar plasmids (linear arrows with yellow highlighted background) are shown in (**B**); regions of alignment are shown as red-filled portions whilst non-aligned areas are shown as empty arrows. Enlarged section of the plasmid focusing on the resistance genes (genomic resistance island) are shown in (**C**) and (**D**). This plasmid (VXIZ01000004) contains no ESBLs. (**3.2**) Mauve alignment of pKP15.12_OXA-181 and pKP15.4_KATLEGO (**B**) with other plasmids shows rearrangement of blocks (sections) of the closely related plasmids. Sections of the aligned plasmid genomes that aligns perfectly have the same colour.

**Figure S4.**
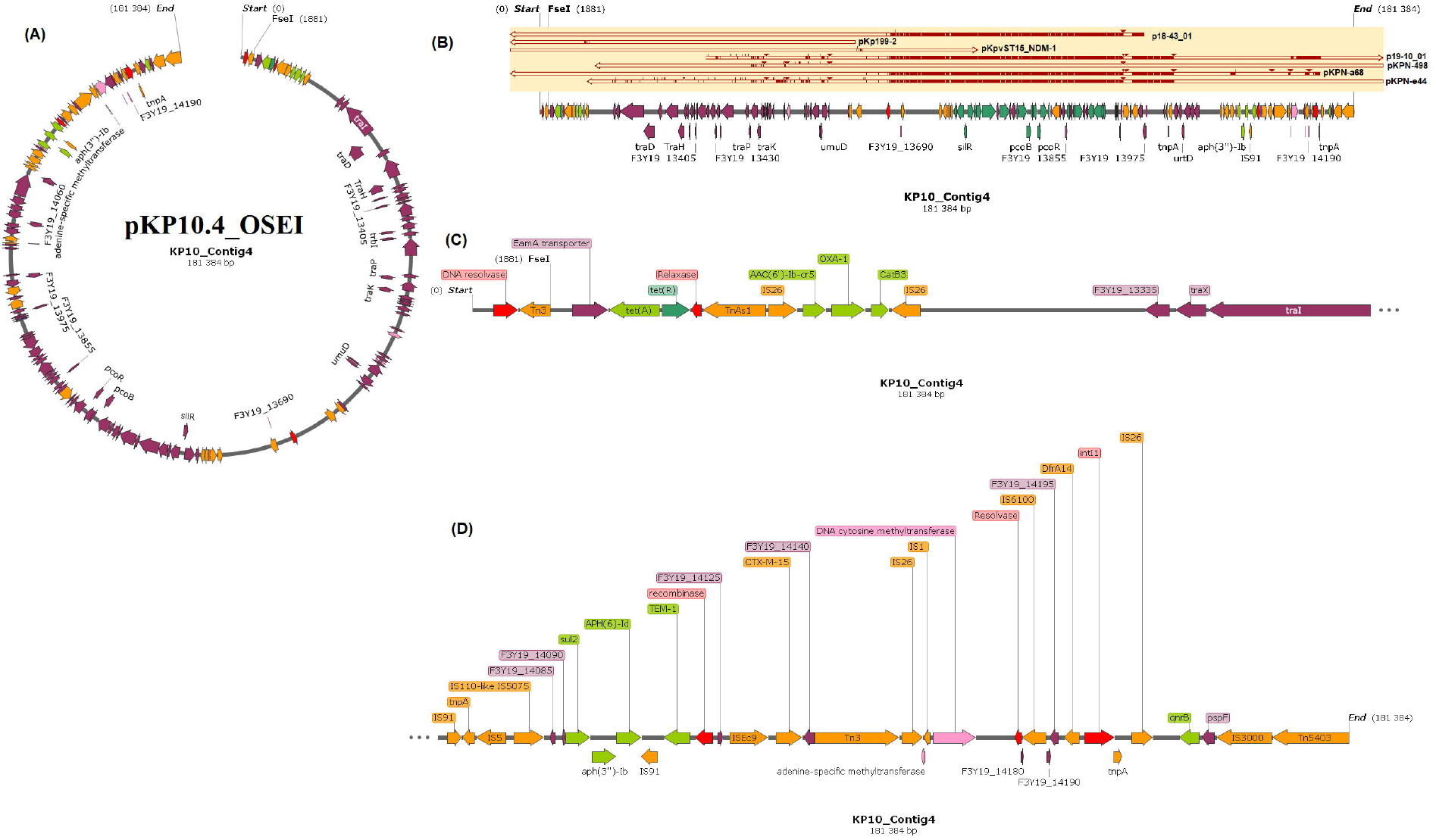
Graphical annotation and comparative alignment of pKP29.8_CTXM-15 and pK29.13_MBELLE (4.1 & 4.2) The resistance genes (light green-coloured arrows), mercury resistance genes (deep green-coloured arrows), methyltransferases/restriction endonucleases (rose-coloured arrows), transposons (orange-coloured arrows), insertion sequences (orange-coloured arrows), integrons (red-coloured arrows), resolvases (red-coloured arrows), and recombinases/integrases (red-coloured arrows) on the plasmid is shown with their orientation (direction of arrow), synteny and immediate environment. Other genes with unknown functions are hidden to make the image less cluttered. A circular version of the plasmid is shown in (**A**). A linear version of the plasmid and its alignment with other similar plasmids (linear arrows with yellow highlighted background) are shown in (**B**); regions of alignment are shown as red-filled portions whilst non-aligned areas are shown as empty arrows. Enlarged section of the plasmid focusing on the resistance genes (genomic resistance island) are shown in (**C**). These plasmids (VXIY01000008 & VXIY01000013) contain CTX-M-15 and no ESBLs respectively. (**4.3**) Mauve alignment of pKP29.8_CTXM-15 (**A**), pK29.13_MBELLE and pKP29.11_NDM-7 (**C**) with other plasmids shows rearrangement of blocks (sections) of the closely related plasmids. Sections of the aligned plasmid genomes that aligns perfectly have the same colour.

**Figure S5.**
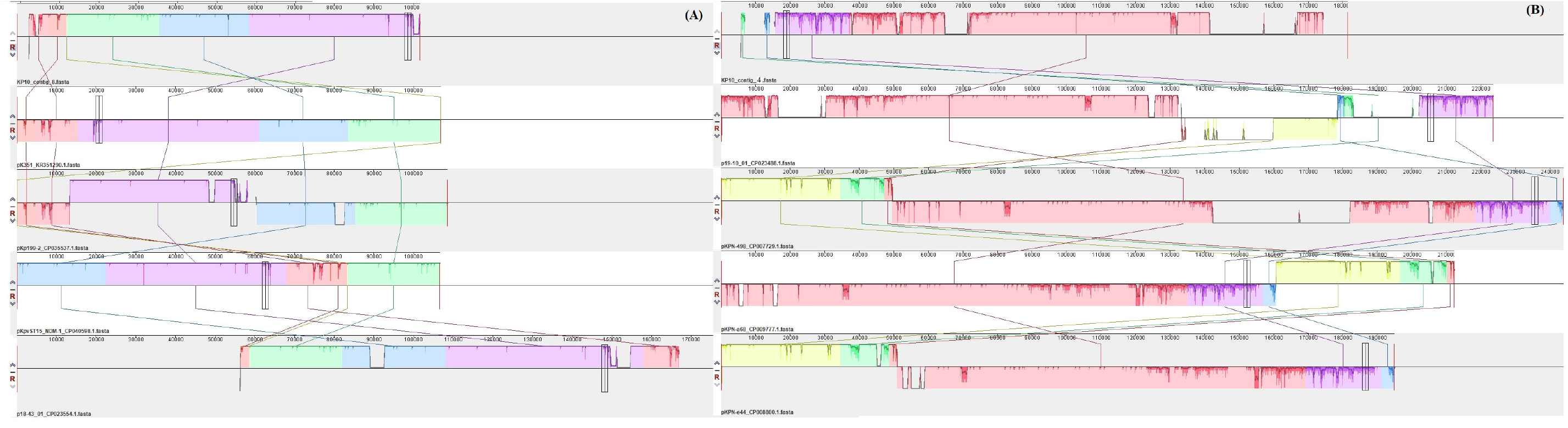
Graphical annotation and comparative alignment of pKP32.3_CTXM-15. **(5.1)** The resistance genes (light green-coloured arrows), replicase and transporter genes (blue-coloured arrows), mercury resistance and tolA genes (deep green-coloured arrows), methyltransferases/restriction endonucleases (rose-coloured arrows), transposons (orange-coloured arrows), insertion sequences (orange-coloured arrows), integrons (red-coloured arrows), resolvases (red-coloured arrows), and recombinases/integrases (red-coloured arrows) on the plasmid is shown with their orientation (direction of arrow), synteny and immediate environment. Other genes with unknown functions are hidden to make the image less cluttered. A circular version of the plasmid is shown in (**A**). A linear version of the plasmid and its alignment with other similar plasmids (linear arrows with yellow highlighted background) are shown in (**B**); regions of alignment are shown as red-filled portions whilst non-aligned areas are shown as empty arrows. Enlarged section of the plasmid focusing on the resistance genes (genomic resistance island) are shown in (**C**) and (**D**). This plasmid (VXIX01000003) contains CTX-M-15. (**5.2**) Mauve alignment of pKP32.3_CTXM-15 (**A**) and pKP32.5_OXA-48 (**B**) with other plasmids shows rearrangement of blocks (sections) of the closely related plasmids. Sections of the aligned plasmid genomes that aligns perfectly have the same colour.

**Figure S6.**
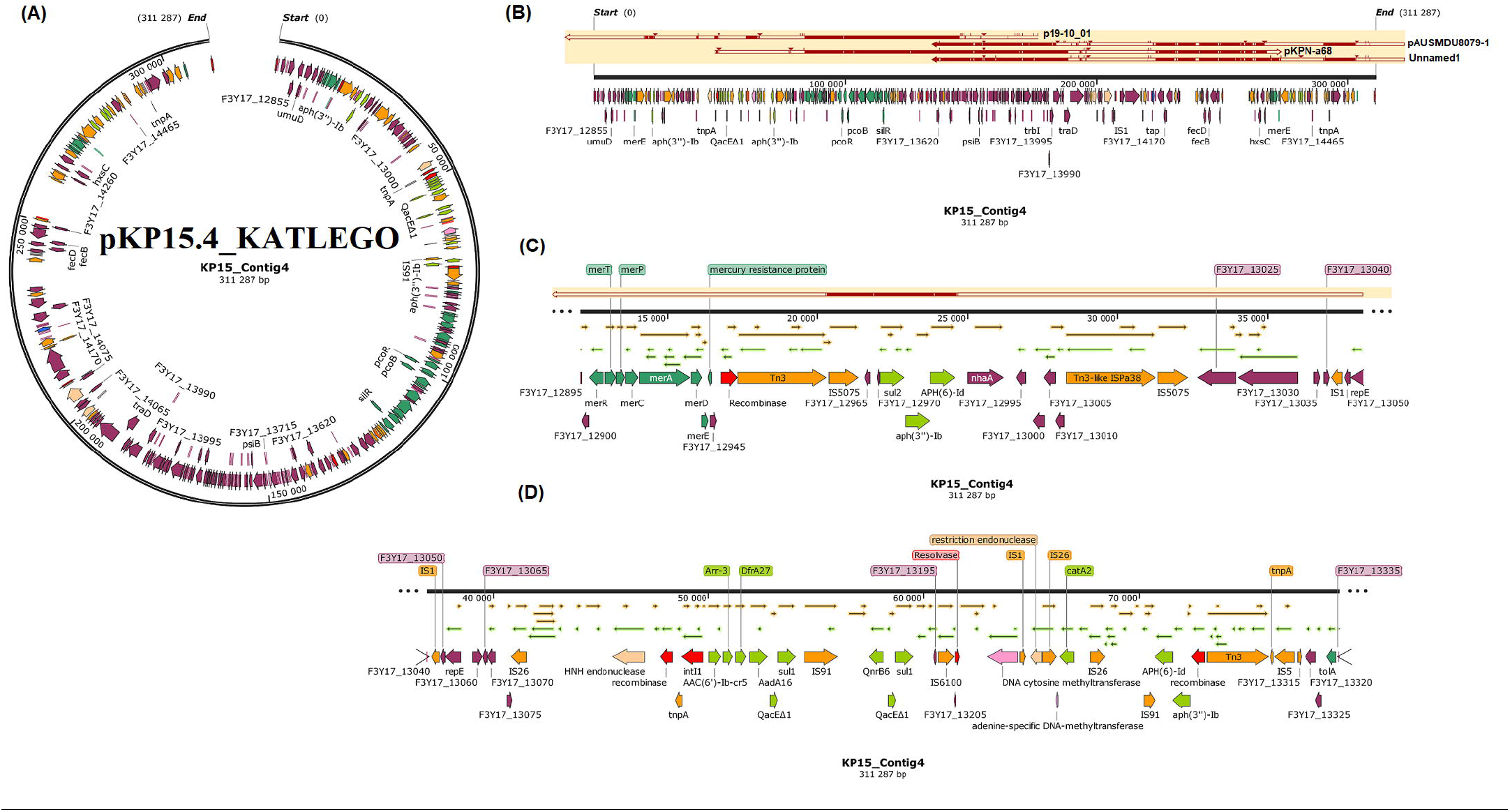
Graphical annotation and comparative alignment of pKP33.4_OSEI and pKP33.8_NDM-1. **(6.1)** The resistance genes (light green-coloured arrows), replicase and transporter genes (blue-coloured arrows), mercury resistance and tolA genes (deep green-coloured arrows), methyltransferases/restriction endonucleases (rose-coloured arrows), transposons (orange-coloured arrows), insertion sequences (orange-coloured arrows), integrons (red-coloured arrows), resolvases (red-coloured arrows), and recombinases/integrases (red-coloured arrows) on the plasmid is shown with their orientation (direction of arrow), synteny and immediate environment. Other genes with unknown functions are hidden to make the image less cluttered. A circular version of the plasmid is shown in (**A**). A linear version of the plasmid and its alignment with other similar plasmids (linear arrows with yellow highlighted background) are shown in (**B**); regions of alignment are shown as red-filled portions whilst non-aligned areas are shown as empty arrows. Enlarged section of the plasmid focusing on the resistance genes (genomic resistance island) are shown in (**C**) and (**D**). These plasmids (VXIW01000004 & VXIW01000008) contains CTX-M-15 and NDM-1, respectively. (**6.2 & 6.3**) Mauve alignment of pKP33.8_NDM-1 (**A**) and pKP33.4_OSEI (**B**) with other plasmids shows rearrangement of blocks (sections) of the closely related plasmids. Sections of the aligned plasmid genomes that aligns perfectly have the same colour.

**Figure S7.**
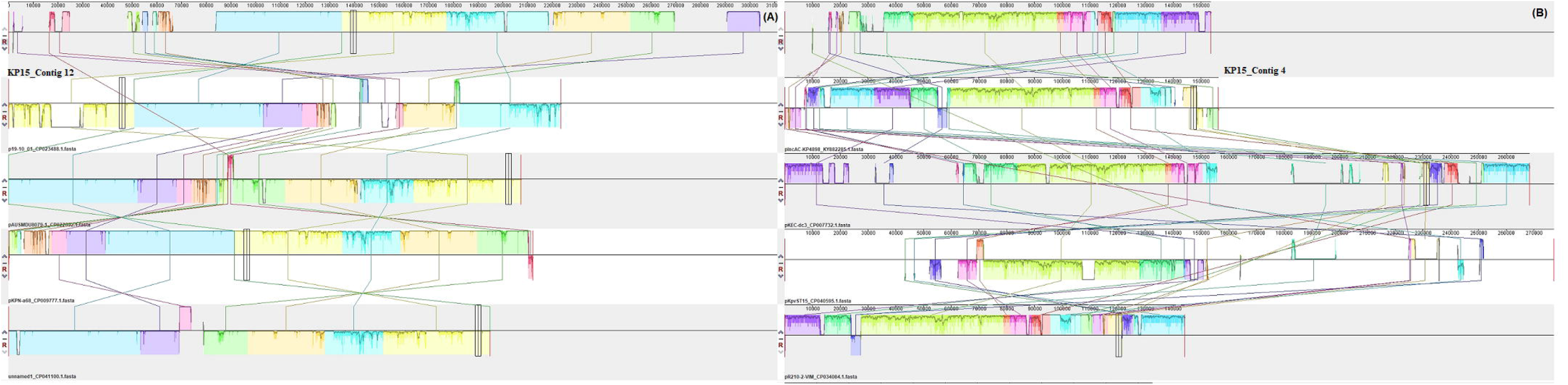
Types and locations (regions) of prophage DNA in isolate KP8. Seven complete prophage DNA and two incomplete prophage DNA were identified in KP8, with two complete prophages being found on plasmids (**7.1**). The protein components of the various prophages are shown in **7.2**

**Figure S8.**
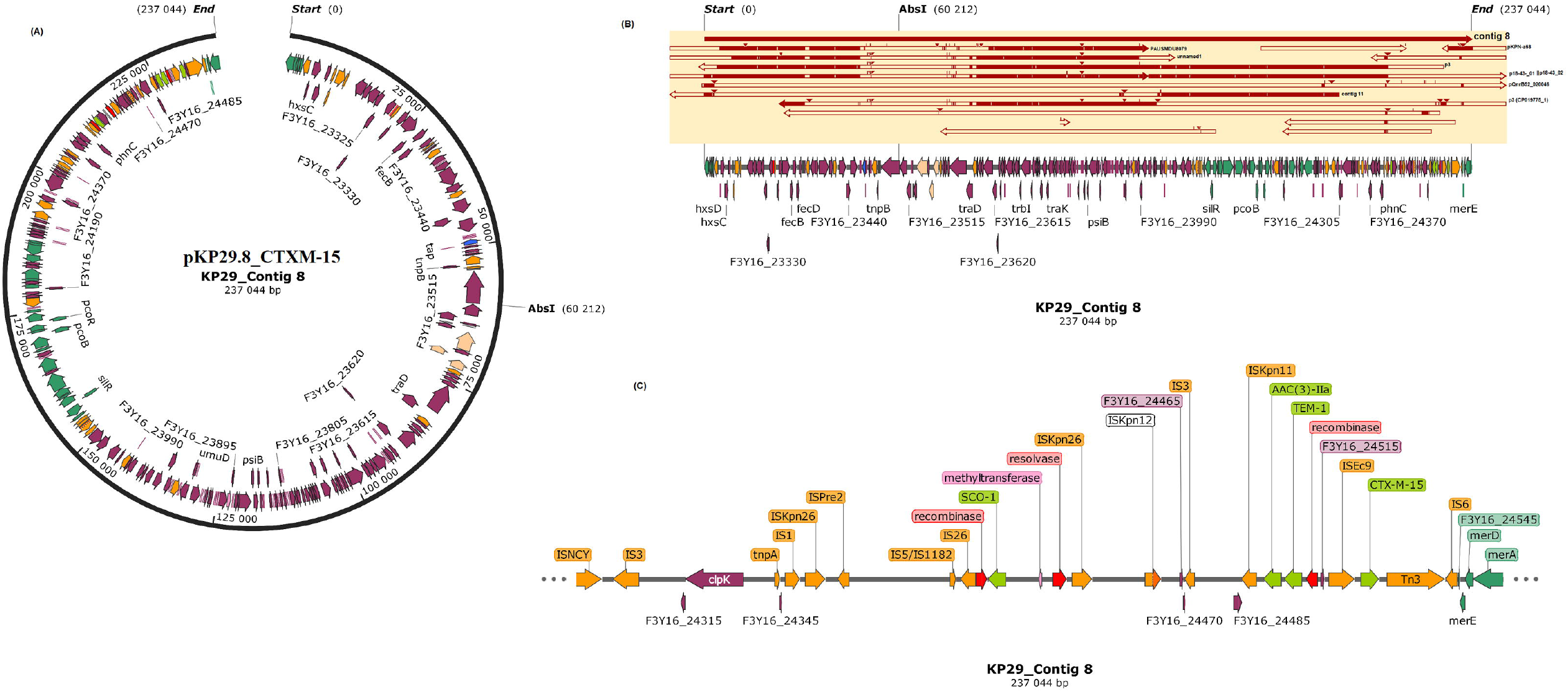
Types and locations (regions) of prophage DNA in isolate KP10. Three complete prophage DNA and four incomplete prophage DNA were identified in KP10, with one complete prophage being found on a plasmid (**8.1**). The protein components of the various prophages are shown in **8.2**

**Figure S9.**
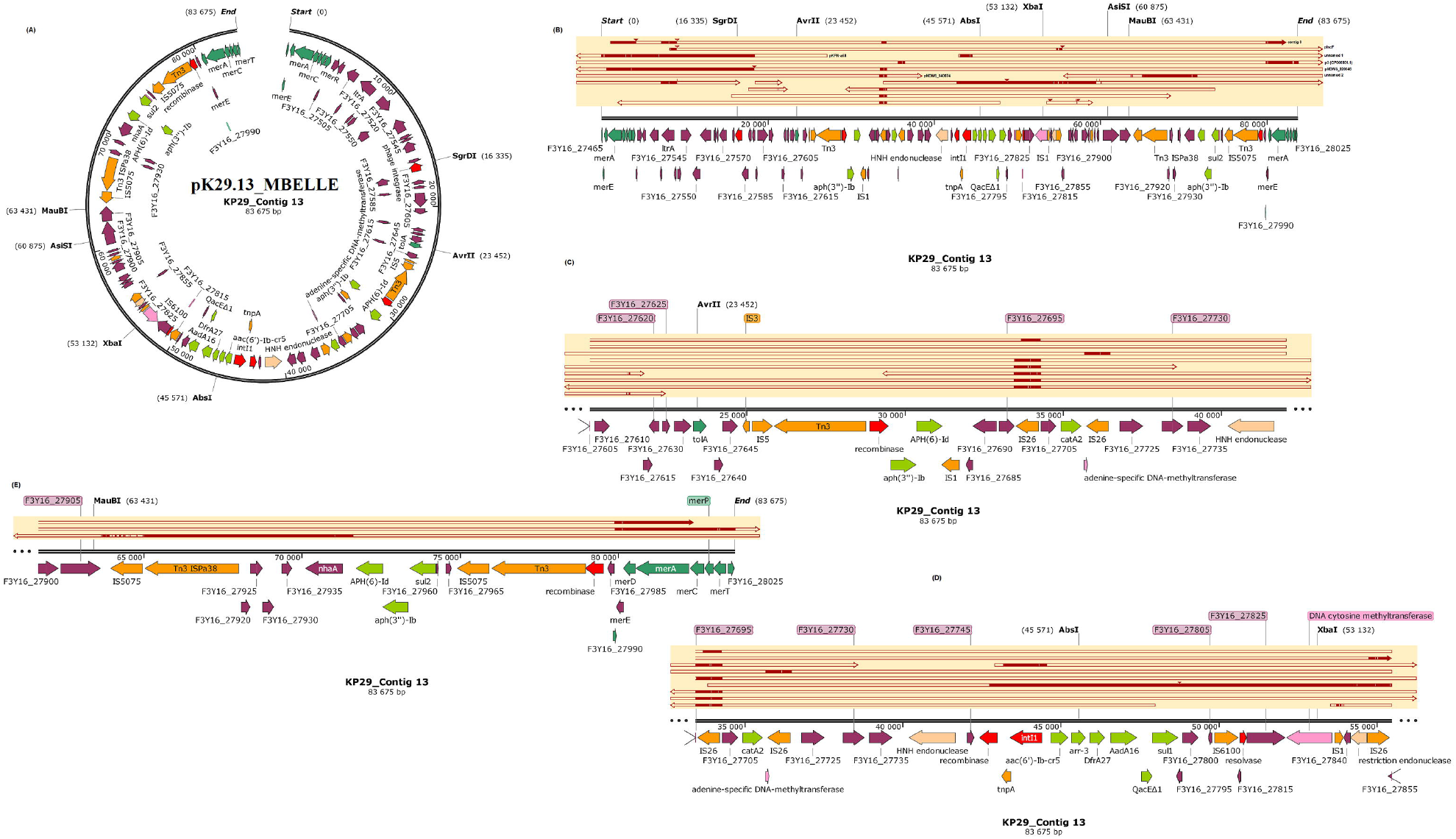
Types and locations (regions) of prophage DNA in isolate KP15. Five complete prophage DNA and two incomplete prophage DNA were identified in KP15, with two complete prophages being found on plasmids (**9.1**). The protein components of the various prophages are shown in **9.2**

**Figure S10.**
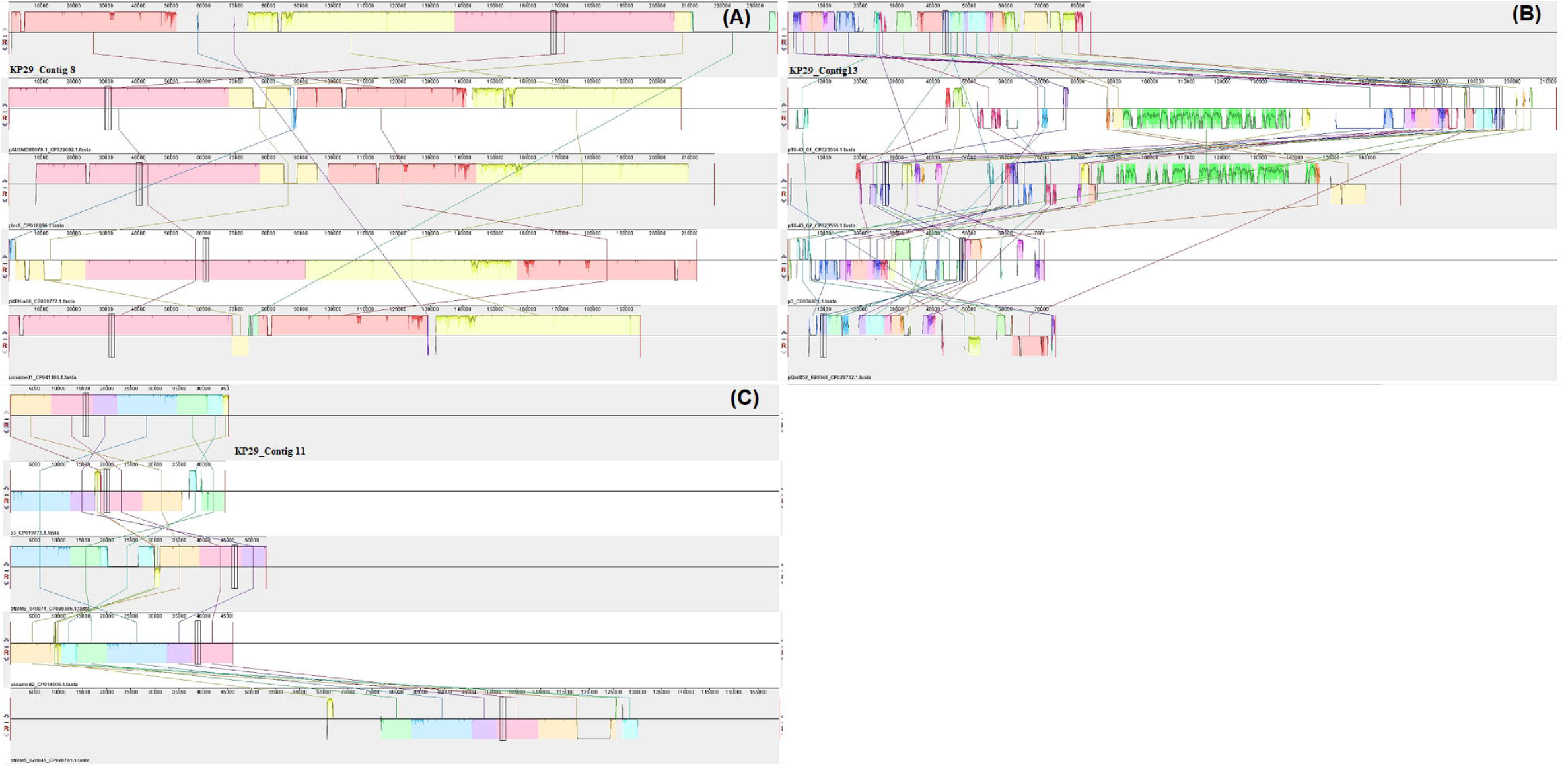
Types and locations (regions) of prophage DNA in isolate KP29. Five complete prophage DNA and three incomplete prophage DNA were identified in KP29, with one complete prophage being found on a plasmid (**10.1**). The protein components of the various prophages are shown in **10.2**

**Figure S11.**
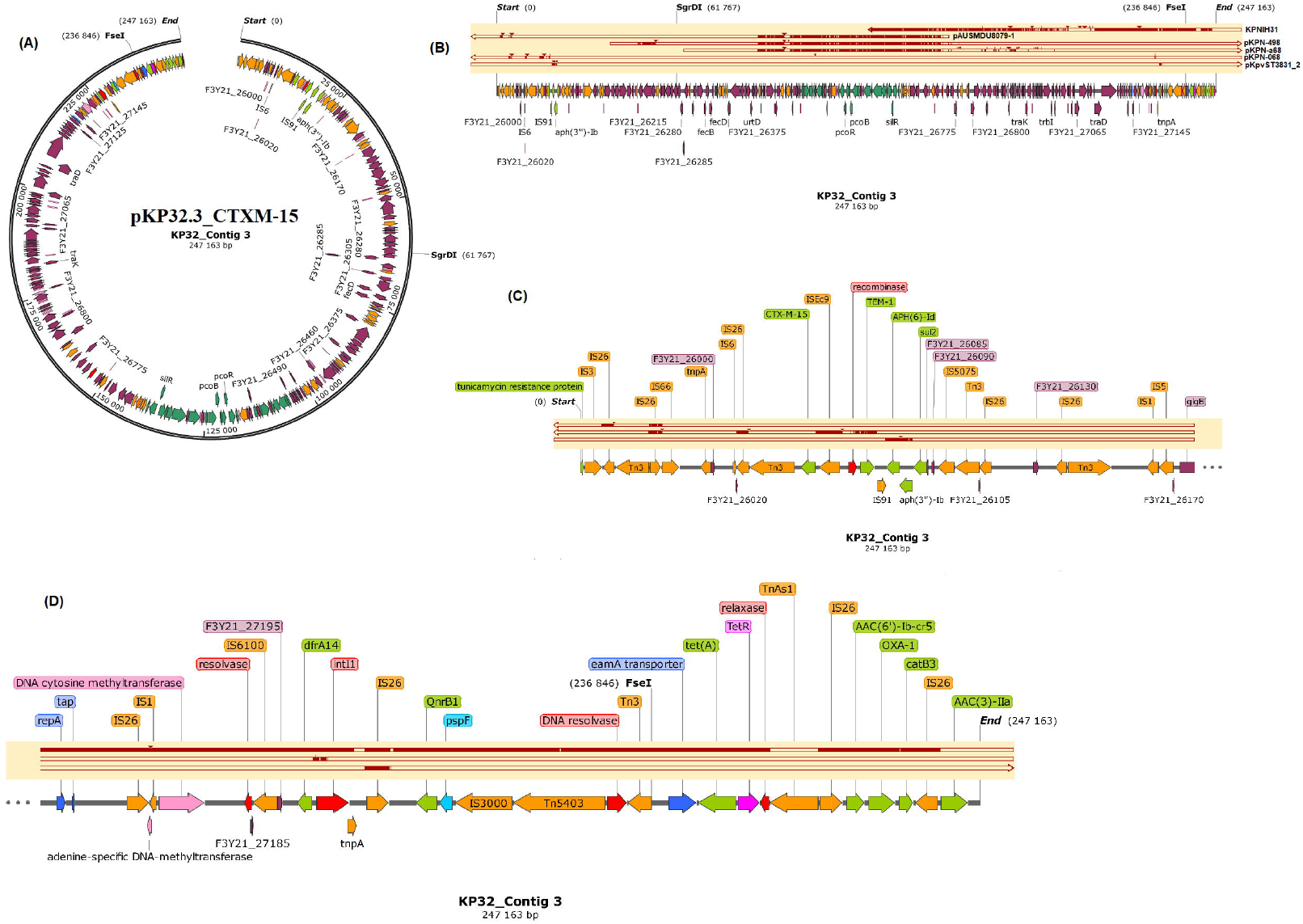
Types and locations (regions) of prophage DNA in isolate KP32. Four complete prophage DNA and four incomplete prophage DNA were identified in KP32, with one complete prophage being found on a plasmid (**11.1**). The protein components of the various prophages are shown in **11.2**

**Figure S12.**
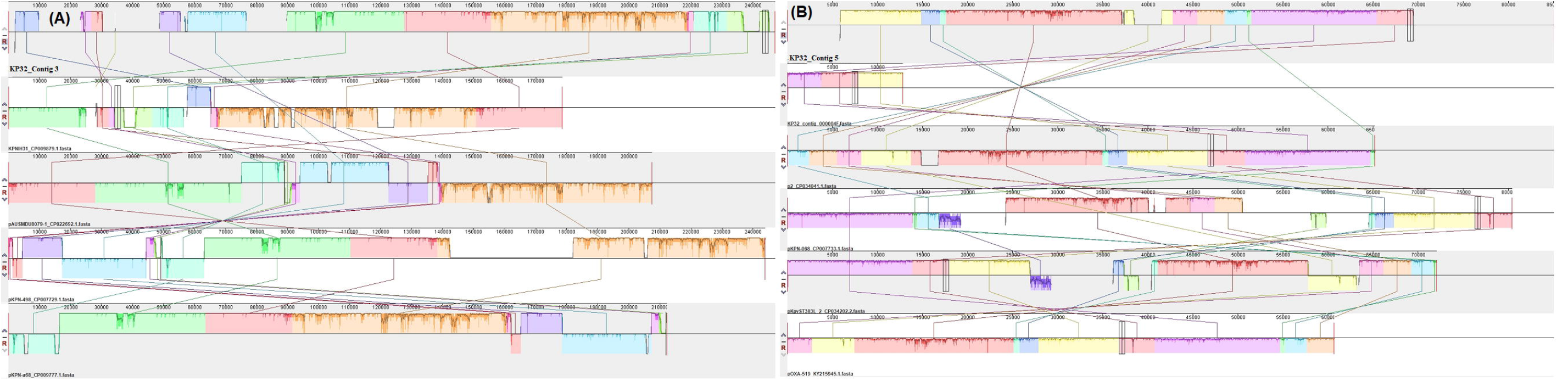
Types and locations (regions) of prophage DNA in isolate KP33. Three complete prophage DNA and four incomplete prophage DNA were identified in KP33, with one complete prophage being found on a plasmid (**33.1**). The protein components of the various prophages are shown in **33.2**

**Figure S13.**
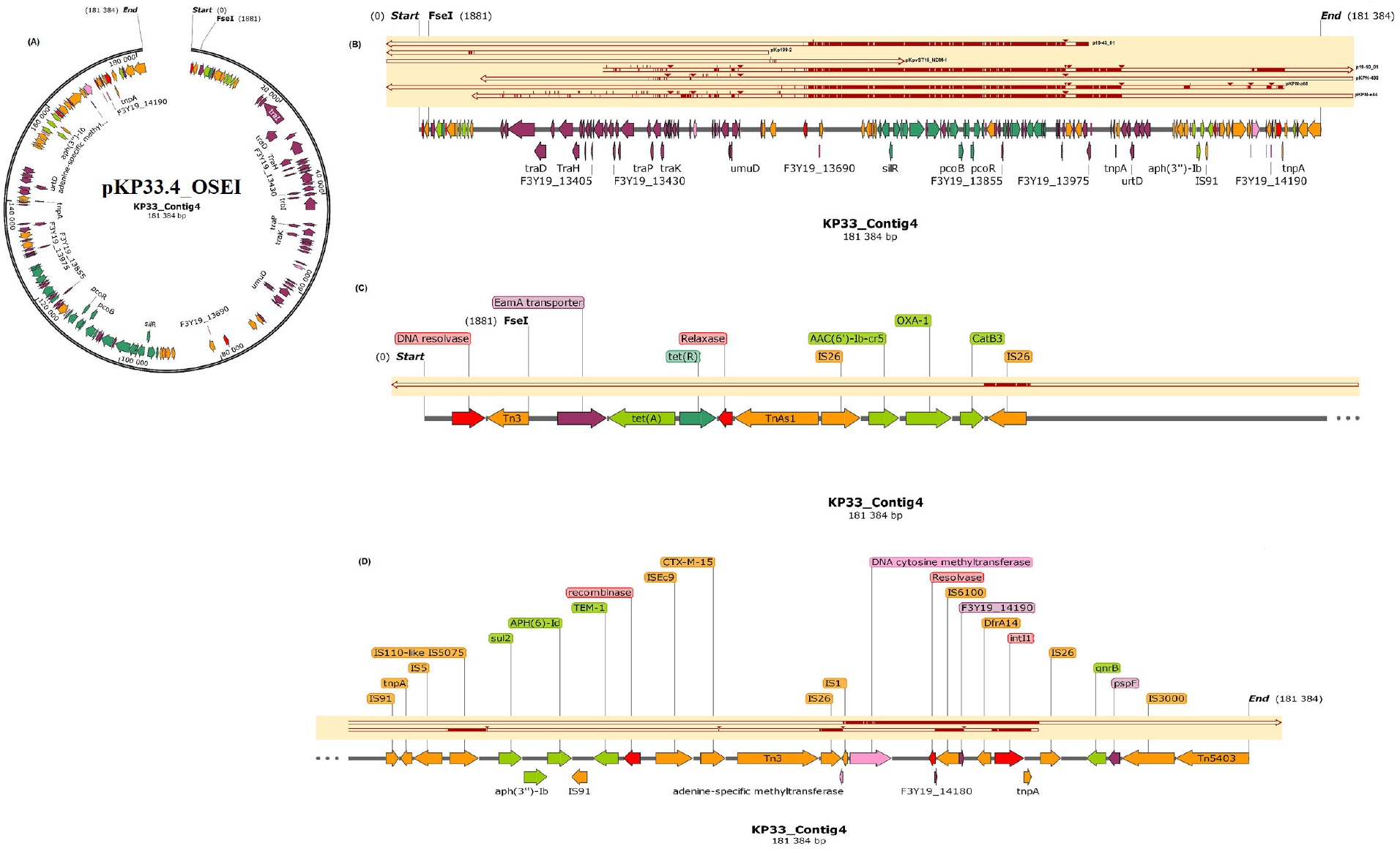
Geographical distribution of plasmids with close nucleotide sequence alignment to this study’s carbapenemase-harbouring plasmids. Closely aligned to pKP8.12_OXA181 plasmids were 201 other plasmids of either IncX_3_ or IncX_3-_ColKP3 replicons from mainly *Escherichia coli* and *Klebsiella pneumoniae* isolates and harbouring mostly *bla*_NDM_ genes, followed by OXA-181, a few KPC and SHV genes; these were mainly from the US & Canada, Europe, South-East Asia, Nigeria and Australia (**13.1**). pKP10.8_NDM-1 and pKP33.8_NDM-1 were closely aligned to 31 other IncFIB(pB171)-IncFII(Yp) plasmids harbouring NDM-1, sul1 and rmtC ARGs, which were isolated from *Enterobacter cloacae/hormaechei, K. pneumoniae/michiganensis, Serratia marcescens, E. coli* etc. from North America, Europe, South-East Asia (**13.2 & 13.6**). Thirty-three IncC plasmids with diverse ARGs but with no carbapenemases, were found to be closely aligned to pKP15.12_OXA-181. These plasmids were mainly obtained from *Proteus mirabilis, E. coli, K. pneumoniae, Vibrio spp*., etc. from the US, Greece, and South-East Asia (**13.3**). pKP29.11_NDM-7 was of very close nucleotide sequence alignment with 231 IncX_3_-harbouring NDM (and SHV on some plasmids) or InX_3_-ColKP3-harbouring OXA-181 plasmids from mainly *E. coli, K. pneumoniae* and other Enterobacterales from North America, Europe, South East Asia, Australia and Nigeria (**13.4**). pKP32.5_OXA-48 aligned closely to 59 IncL (and a few IncM and IncFII) plasmids also harbouring OXA-48 from mainly *E. coli, K. pneumoniae* and other Enterobacterales isolated from North America, Europe, the Middle-East, South East Asia, and Morocco (**13.5**)

**Figure S14.**
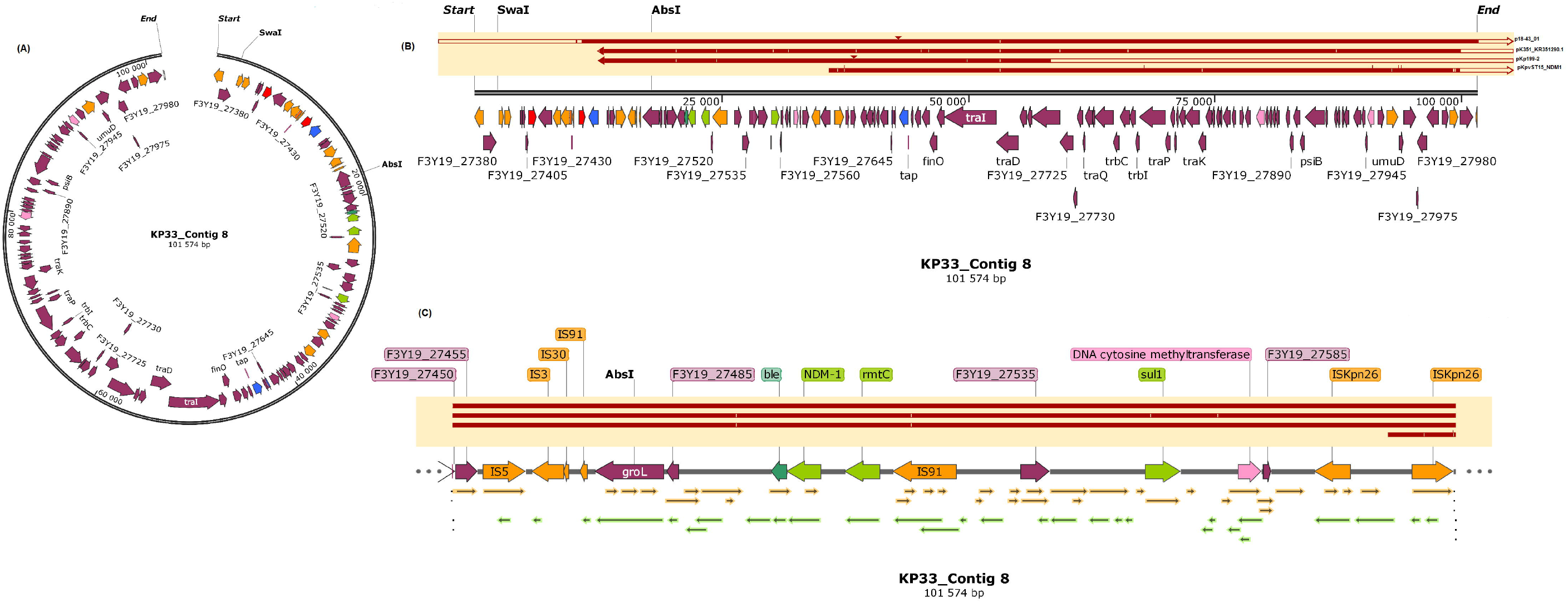
Virulence factors/determinants found in the genomes of the six isolates. A total of 51 virulence genes were identified in the genomes, with all 51 being found in four isolates irrespective of isolation source OR clonality and 40 being found in KP8 and KP32 (**14.1-2**). The K and O capsule typing using Kaptive Web showed diversity among both the K and O capsule types among the isolates. The K-loci results showed four different serotypes among the sequenced isolates: KL2 (KP10 and KP33), KL25 (KP15 and KP29), KL27 (KP32) and KL102 (KP8). As well, the O-loci results also showed four O serotypes: O1v1 (KP10, KP33, and KP15), O2v2 (KP8), O4 (KP32), and O5 (KP29). The K- and O-serotyping was not only clone-specific as different clones shared the same K and O serotypes, suggesting that mutations or evolutions in K and O serotypes can occur in individual cells and are not necessarily conserved within clones (**14.3-8**).

**Supplemental data 1. Primers used to amplify carbapenemases and plasmid replicons used in this study**. This data consists of two Tables (1 & 2): Table 1 is primer sequences used for detecting carbapenemase genes in PCR assays of the K. pneumoniae isolates; and Table 2 is primer sequences used for detecting replicons in CRKP isolates using the PBRT assay.

**Supplemental data 2. Resistance mechanisms of the sequenced *Klebsiella pneumoniae* isolates and their closely related phyletic strains**. This data shows three Tables: Table 1 shows the resistance determinants of the carbapenem-resistant *K. pneumoniae* isolates; Table 2 shows the ARGs and colistin- and fluoroquinolone-resistance-conferring mutations found on chromosomes of the *Klebsiella pneumoniae* strains; and Table 3 shows the comparison of closely related *K. pneumoniae* strains on the phylogeny tree reported in different countries

**Supplemental data 3. Intact phages identified in carbapenem-resistant *K. pneumoniae* clinical isolates**.

This data shows the distribution of intact phages on the genomes of the various *K. pneumoniae* clinical isolates.

**Table S1. Dataset showing the clinical demographics, antibiotic susceptibility profile, REP-PCR-ordered strains, and chromosomal and plasmid characteristics of the isolates**.

**Table S2. Dataset showing the plasmid evolutionary epidemiology and resistomes of this study’s plasmids and that of other closely related plasmids**.

**Table S3. Virulence factor analysis of the *Klebsiella pneumoniae* strains showing the various virulence genes in the six sequenced isolates**.

**Table S4. Methylome data of the *Klebsiella pneumoniae* strains showing the restriction enzymes, type I and type II methylases, their modifications and DNA motifs**.

**Table S5. Genomic metadata of *Klebsiella pneumoniae* strains (genomes) from this study, South Africa, Africa and globally, downloaded from NCBI and PATRIC**

**Table S6. Genomic and resistome metadata of *Klebsiella pneumoniae* strains (genomes) from this study, South Africa, Africa and globally, downloaded from NCBI and PATRIC**

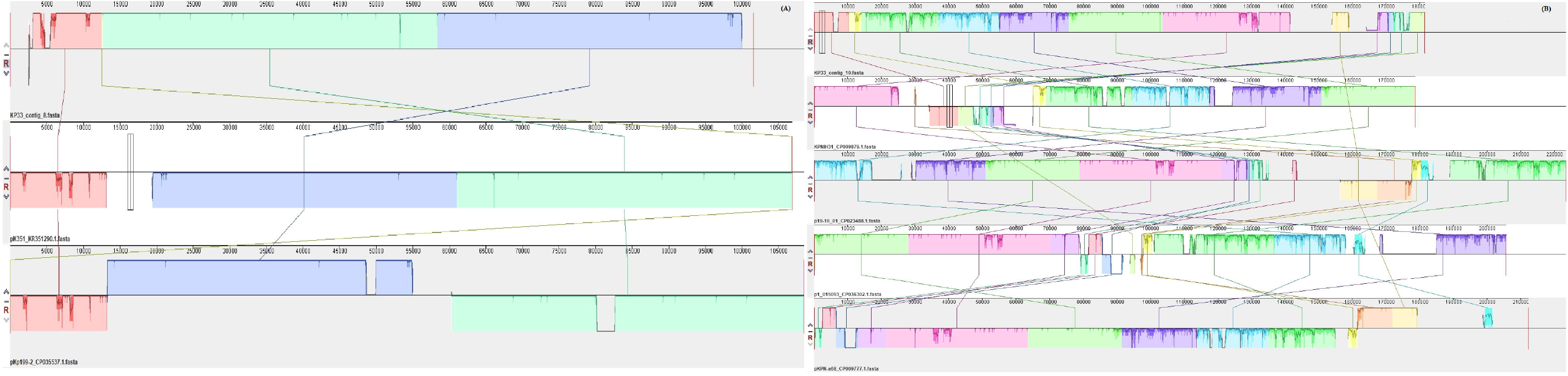

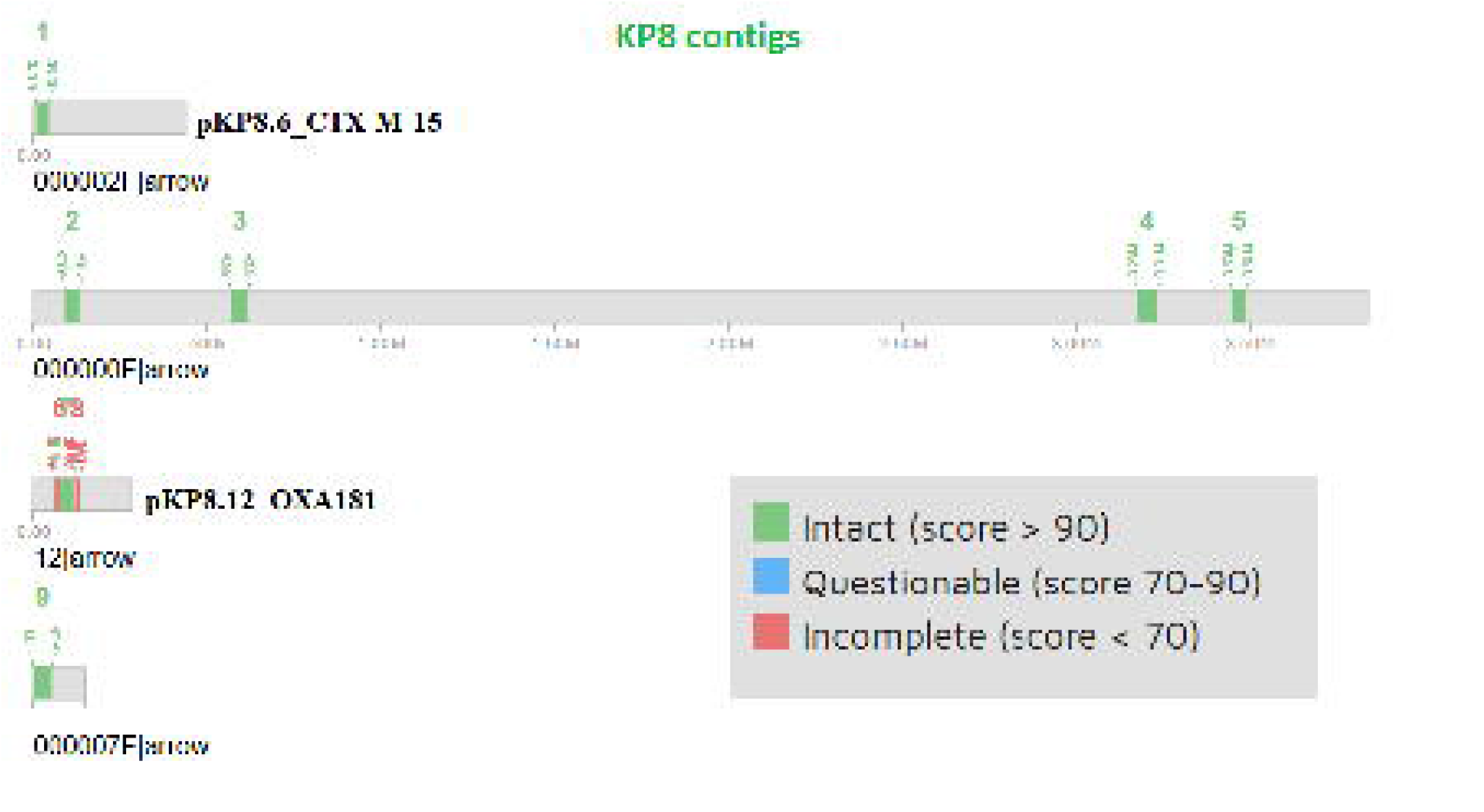

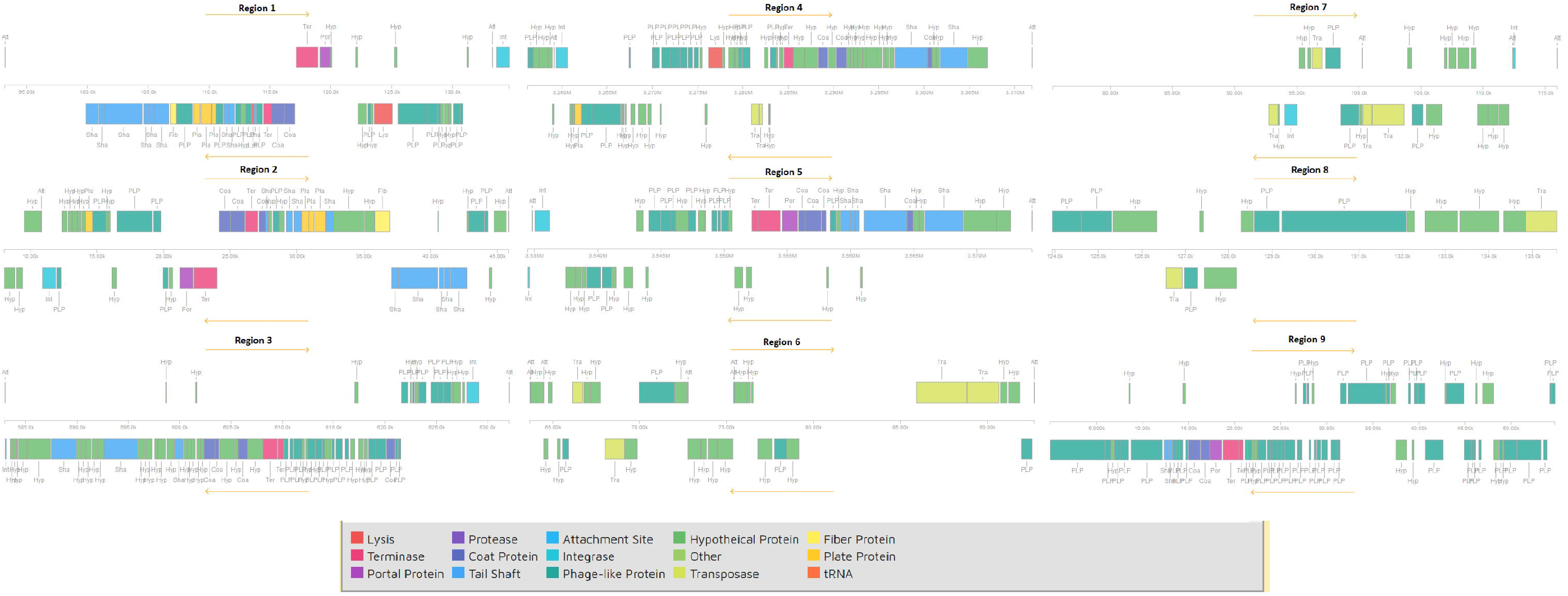

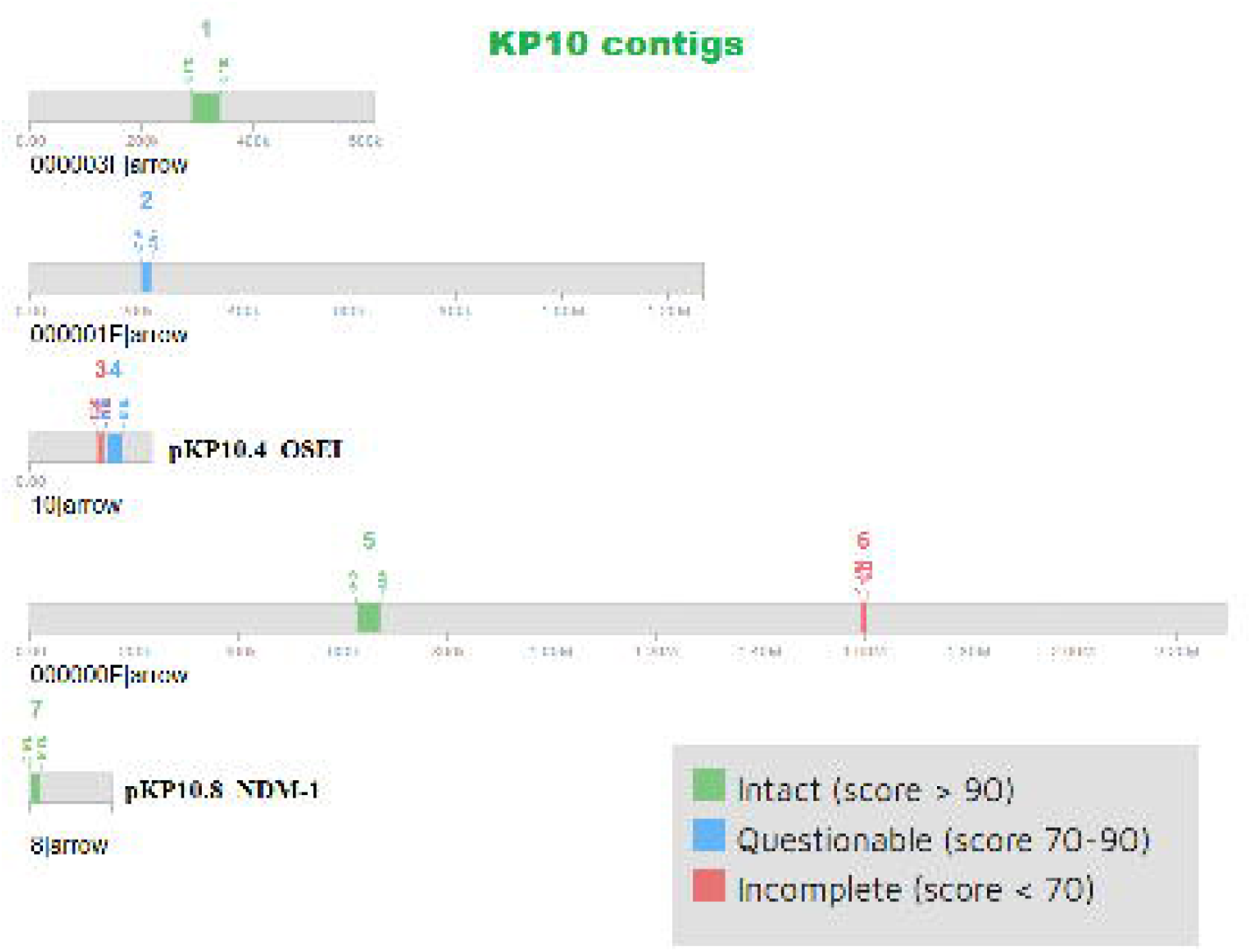

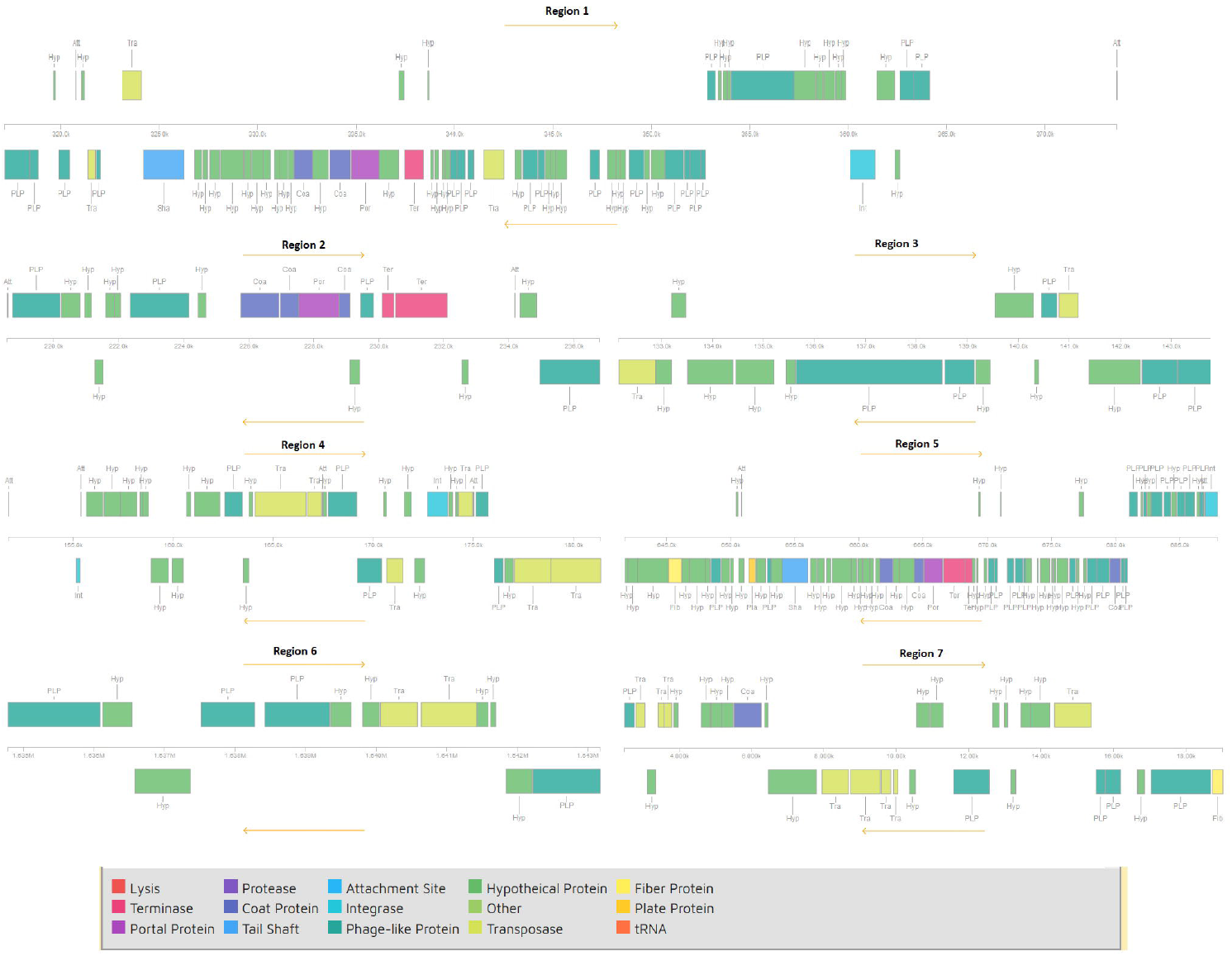

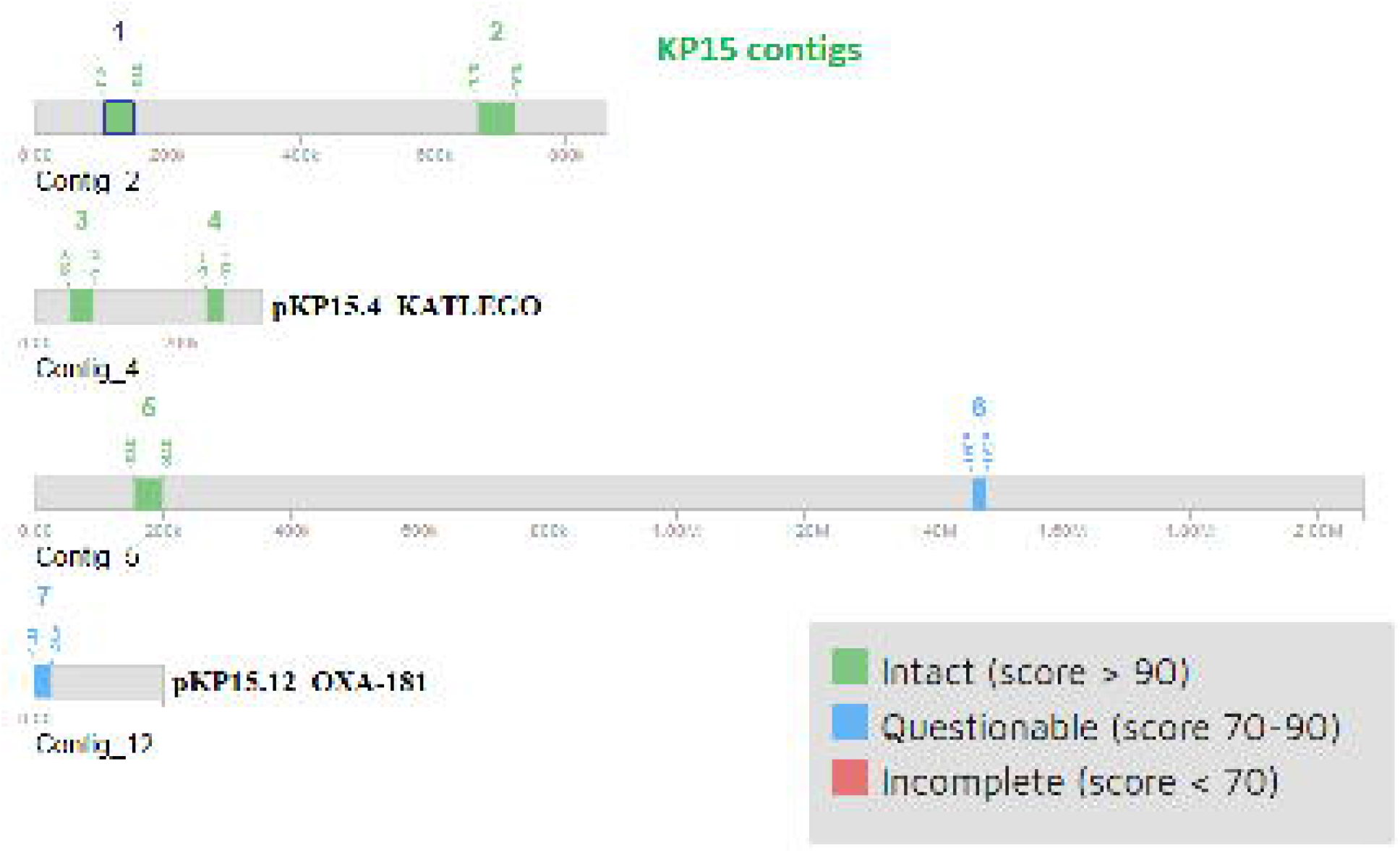

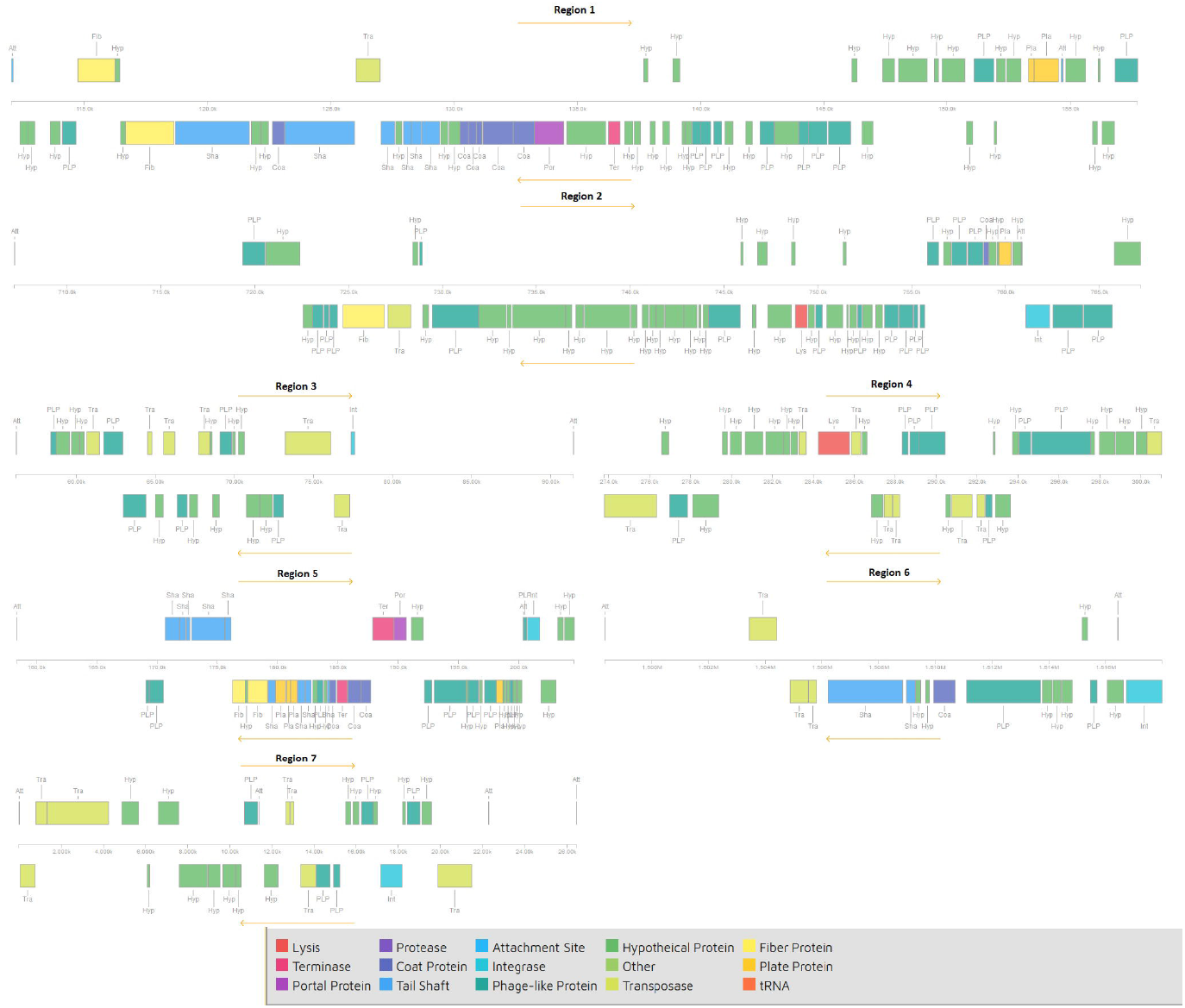

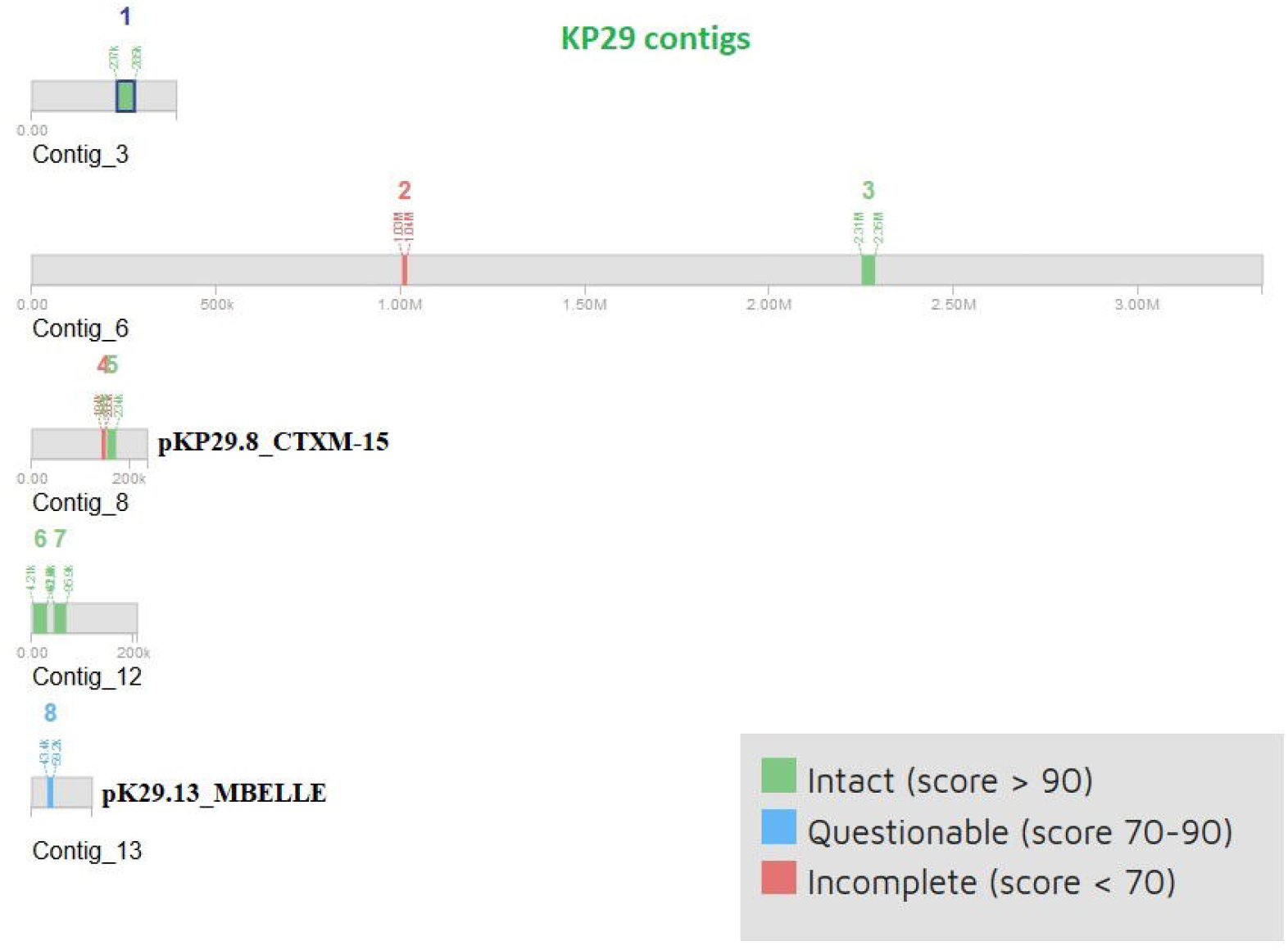

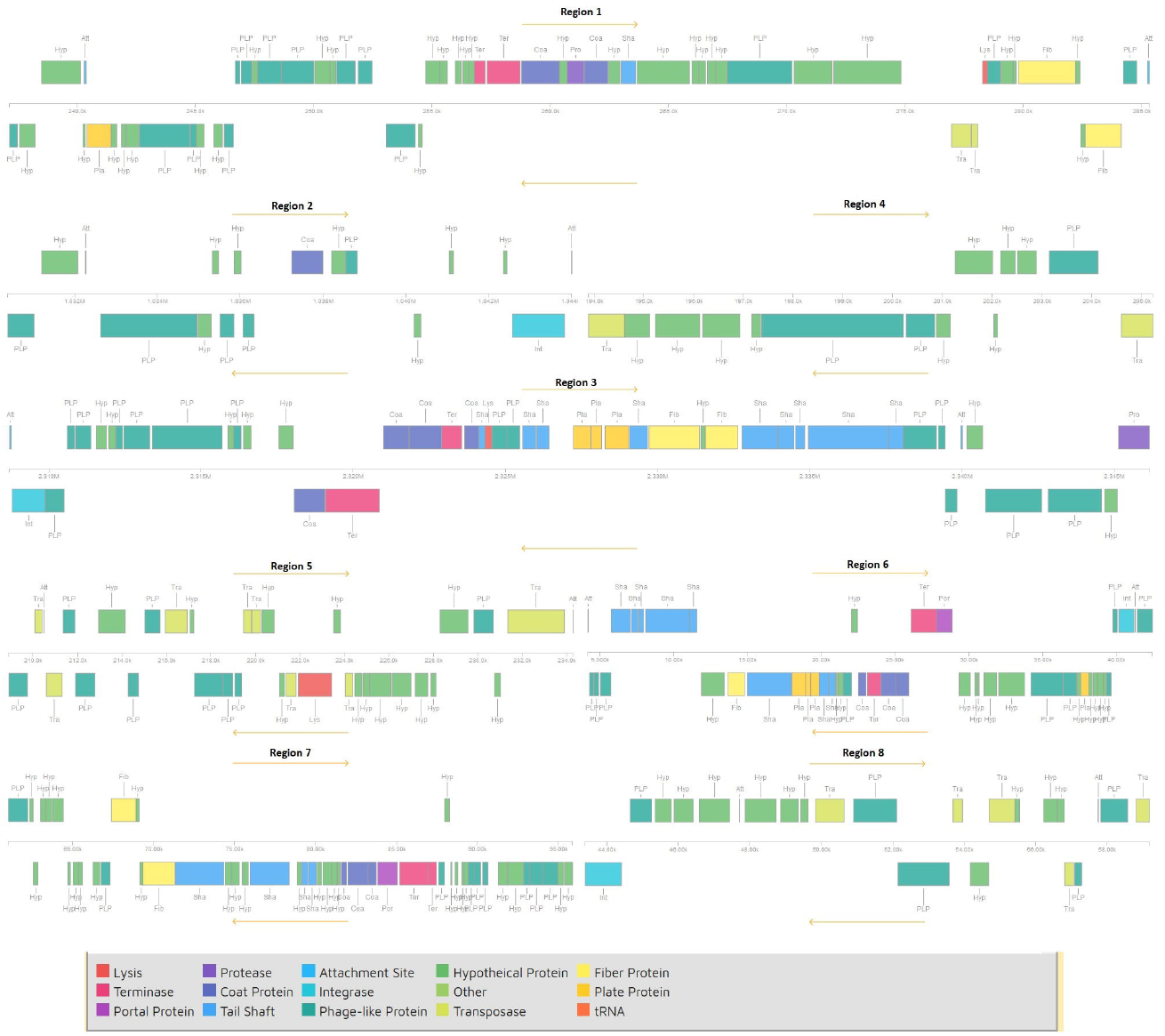

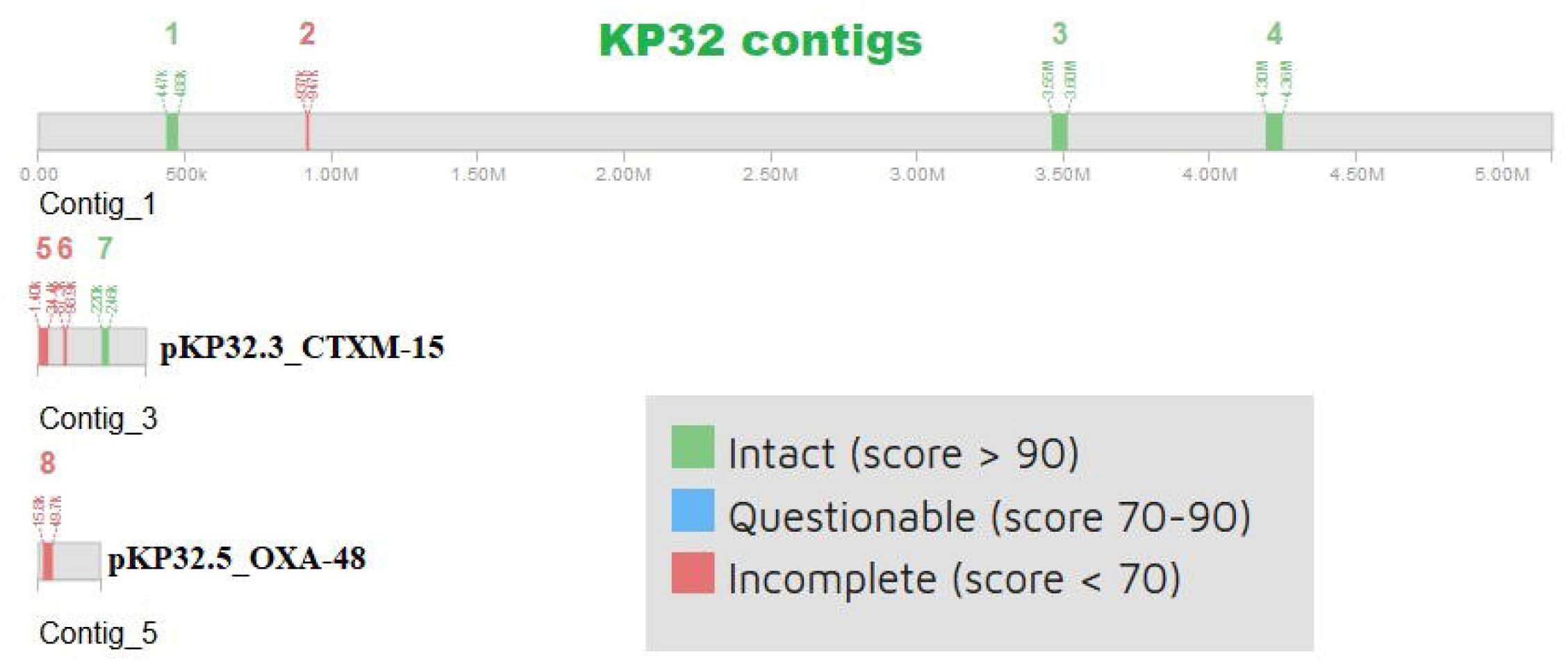

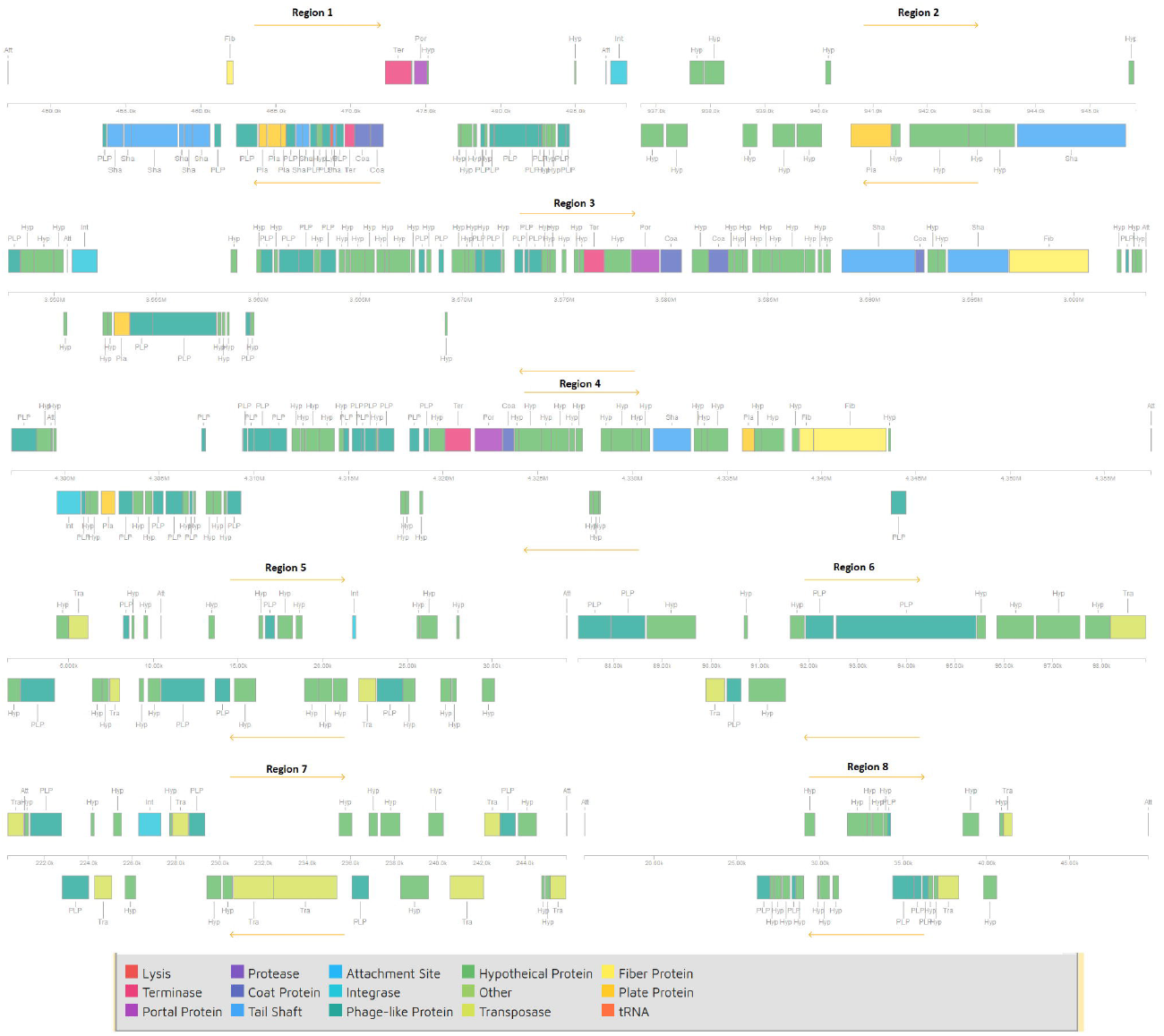

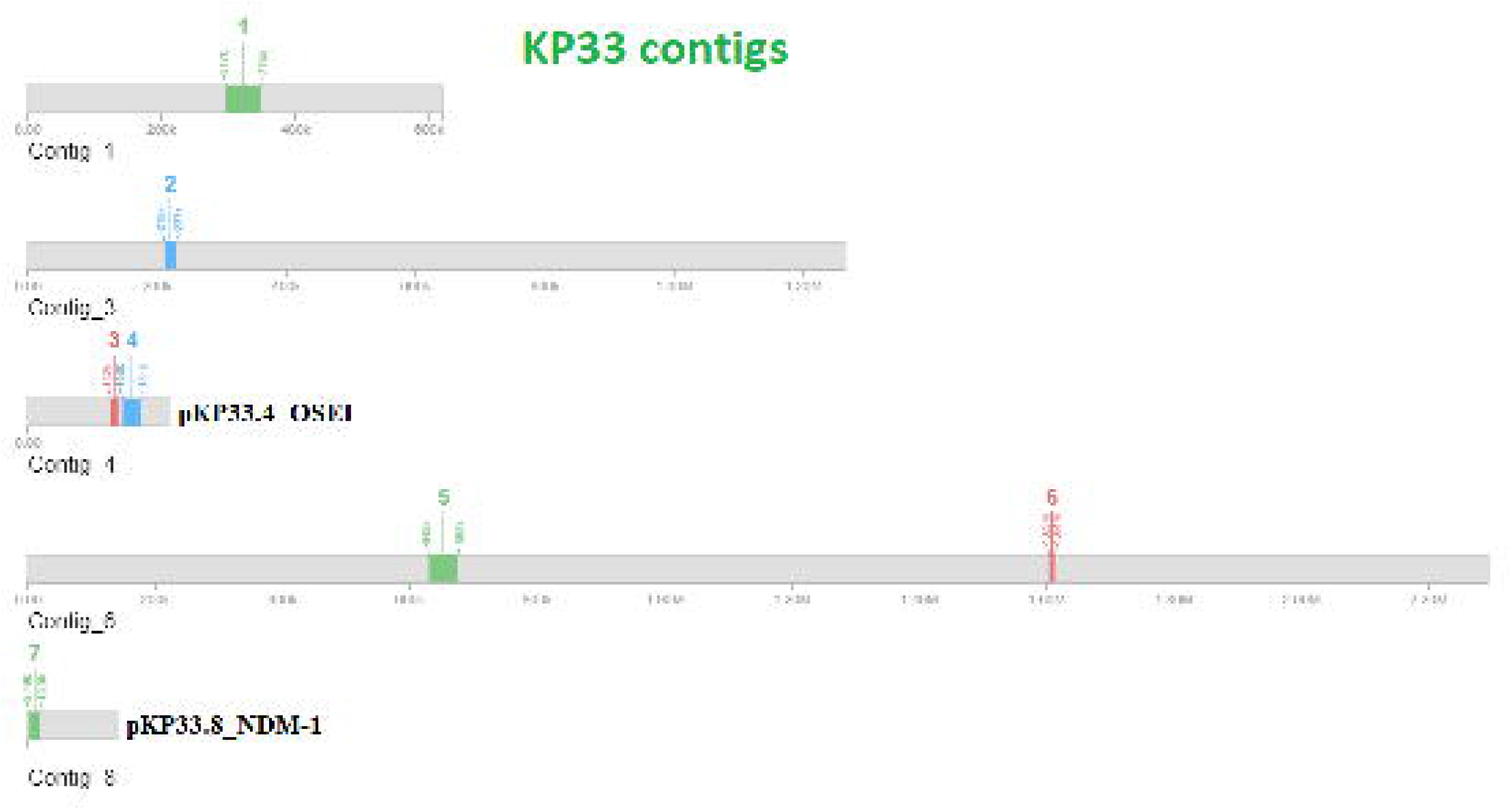

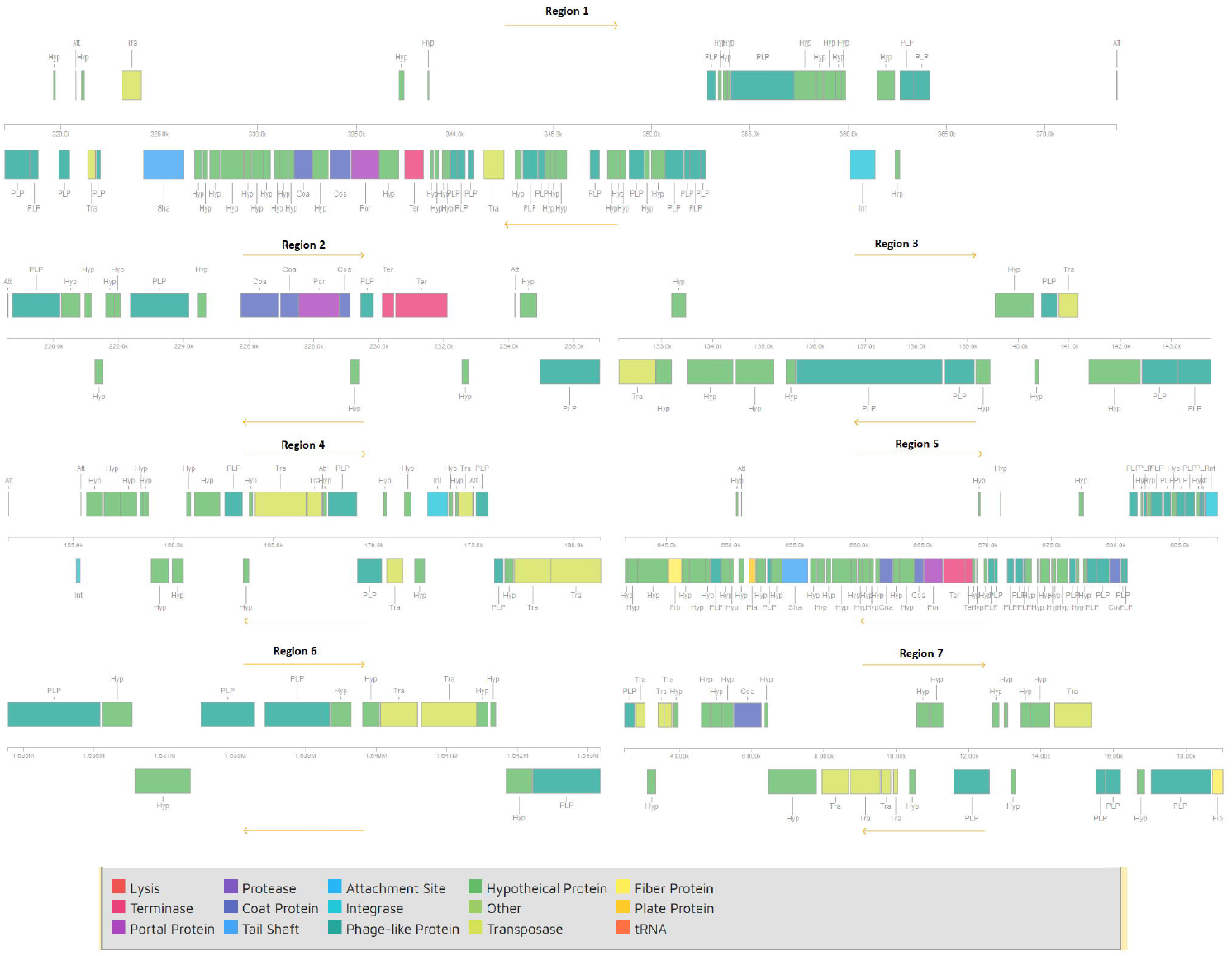

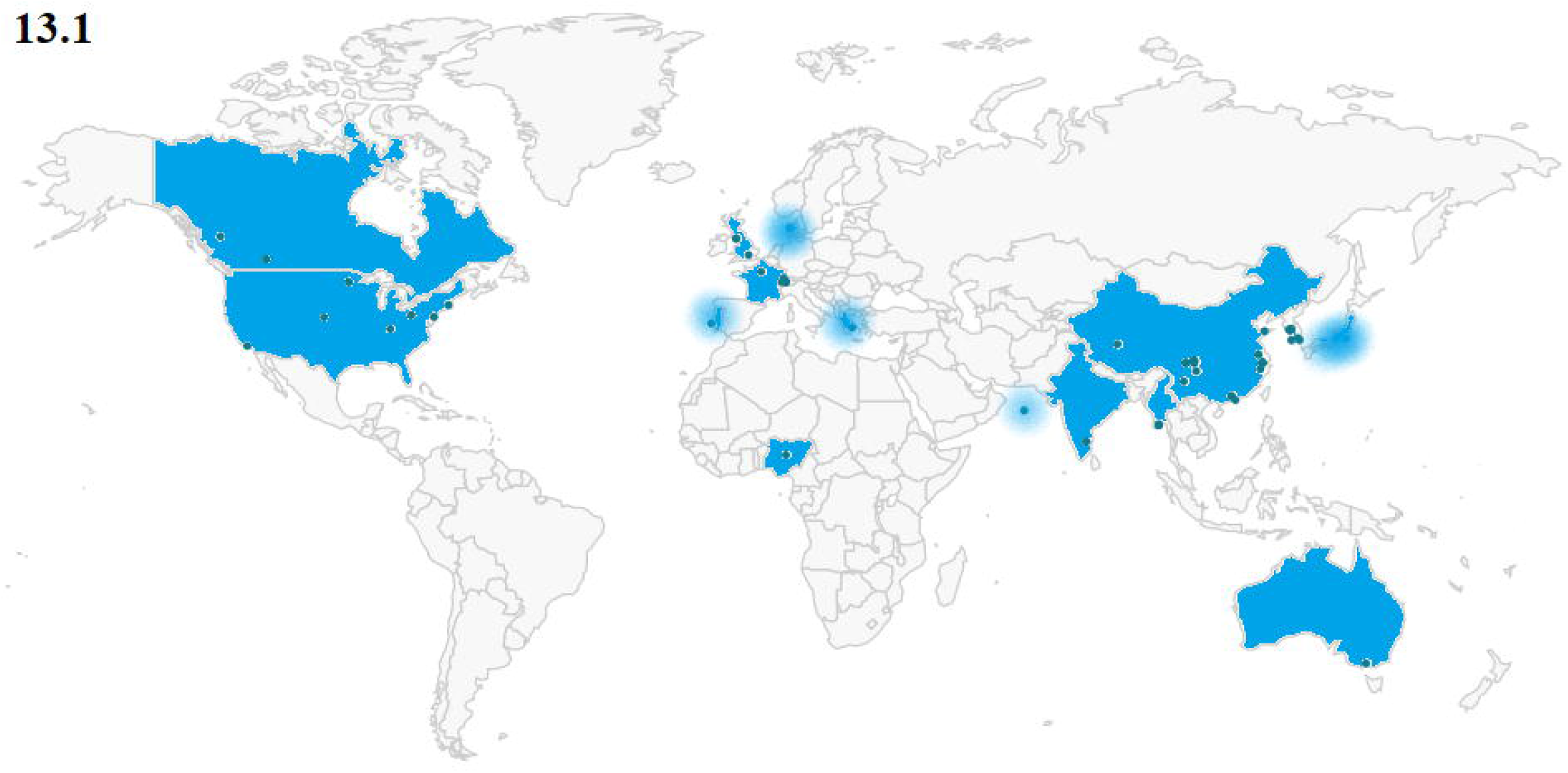

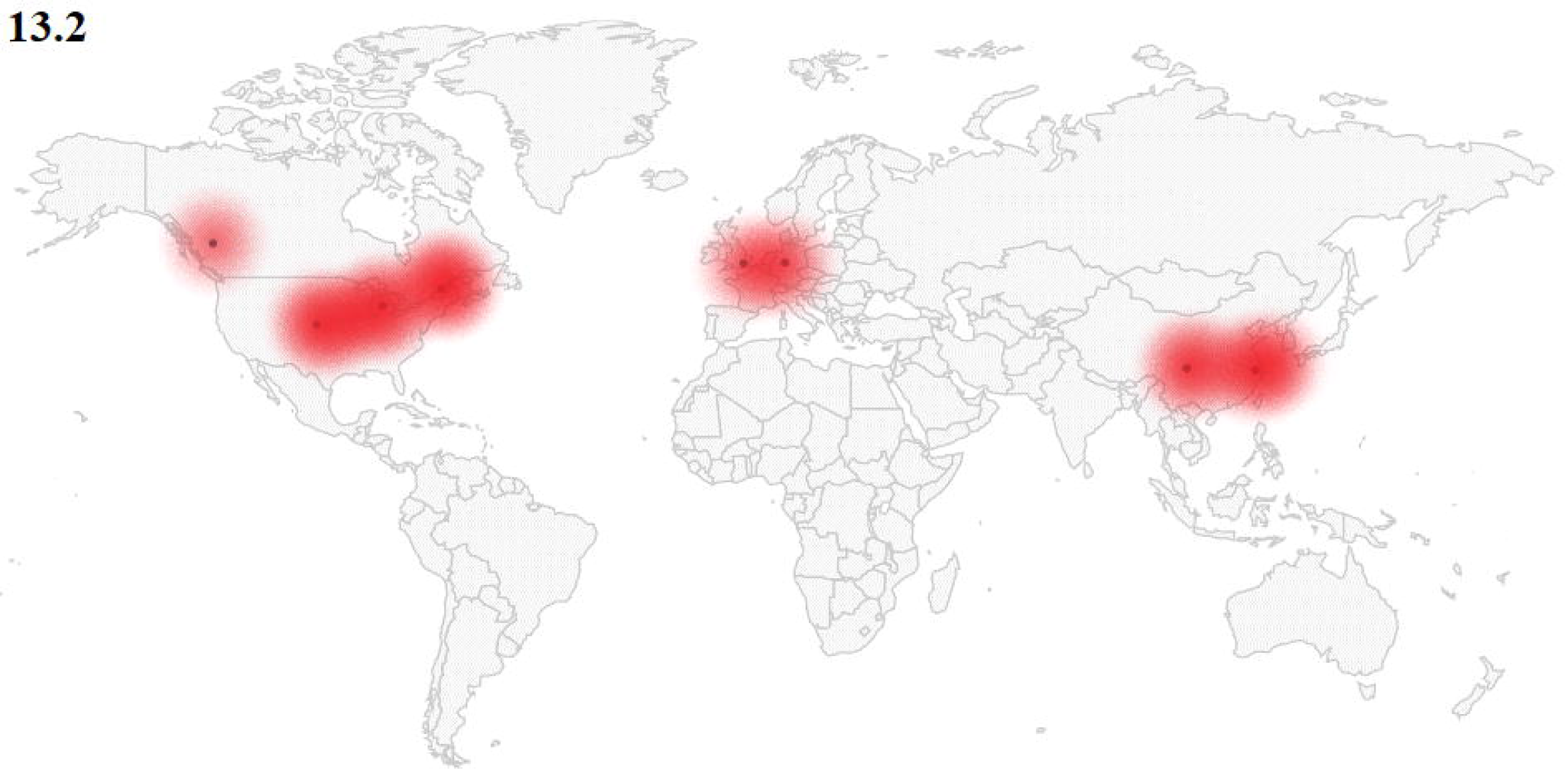

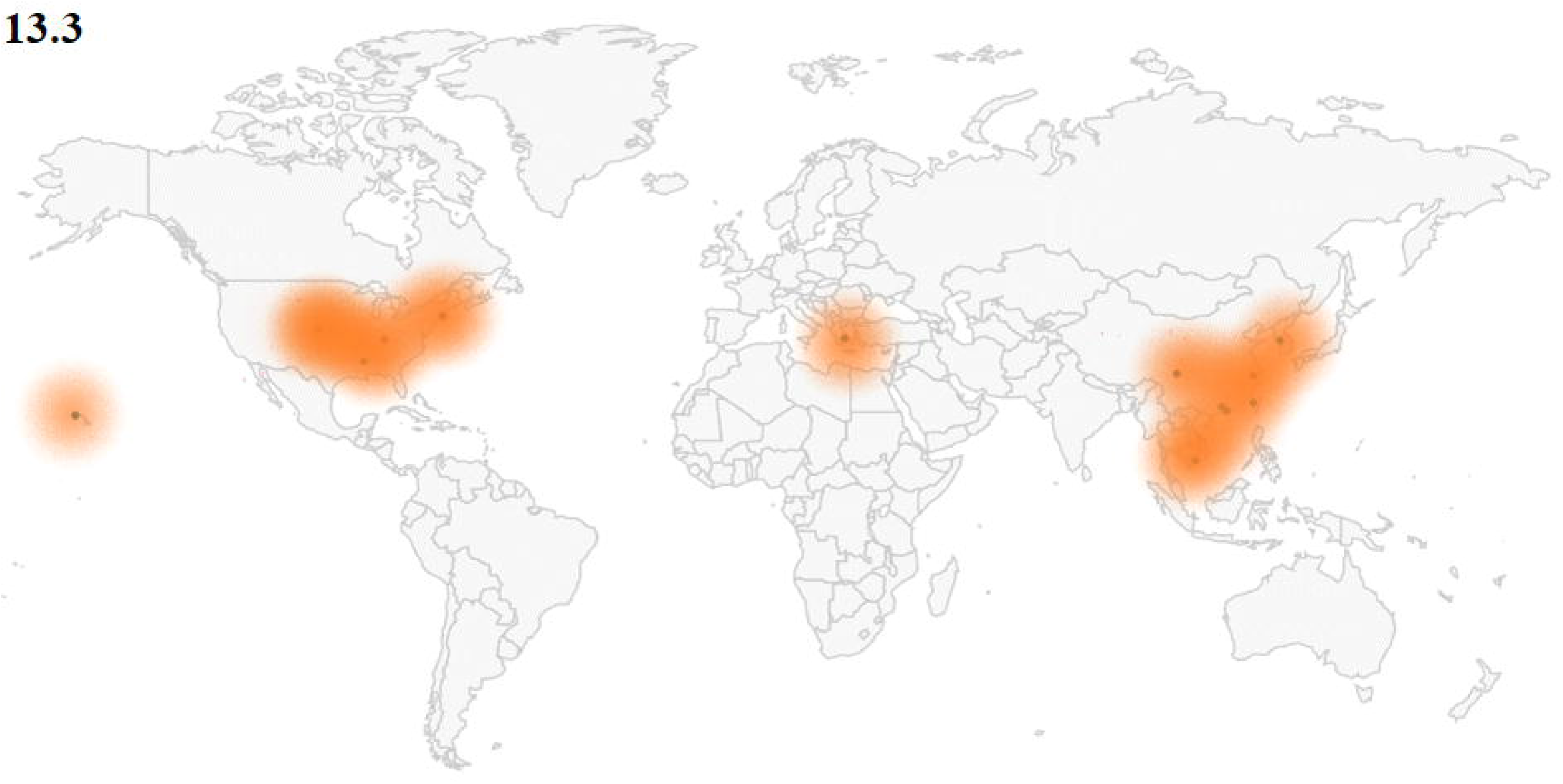

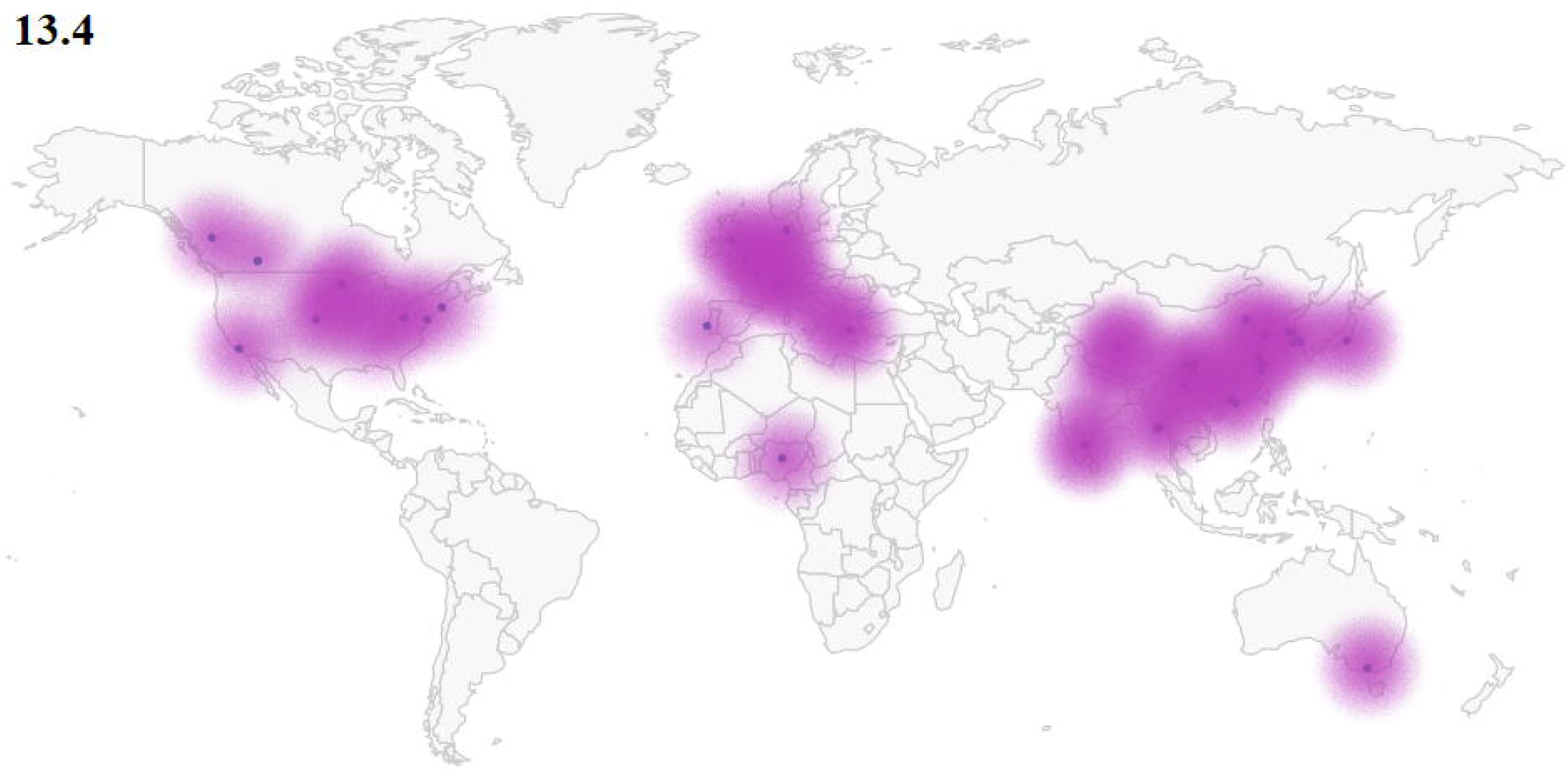

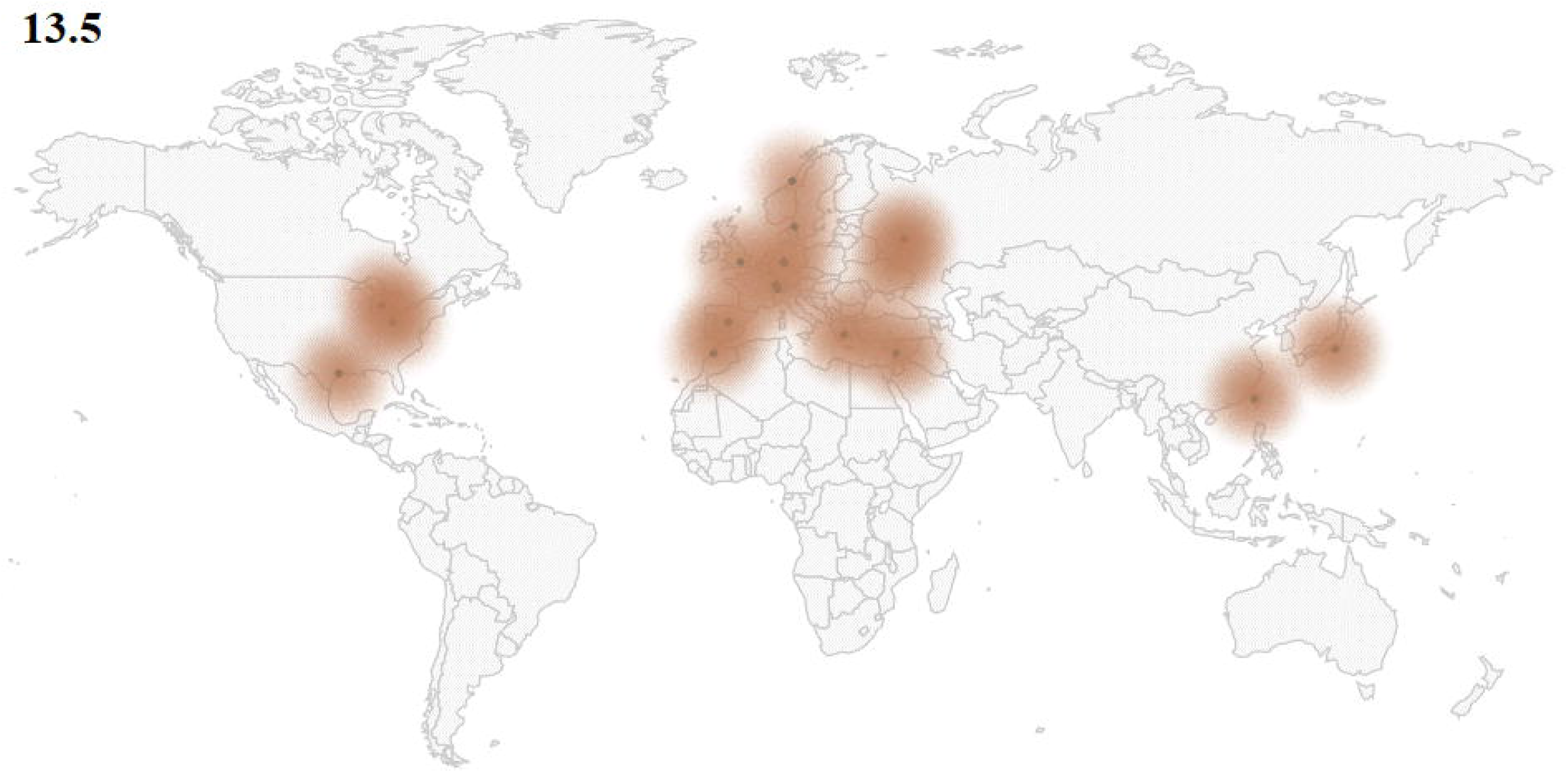

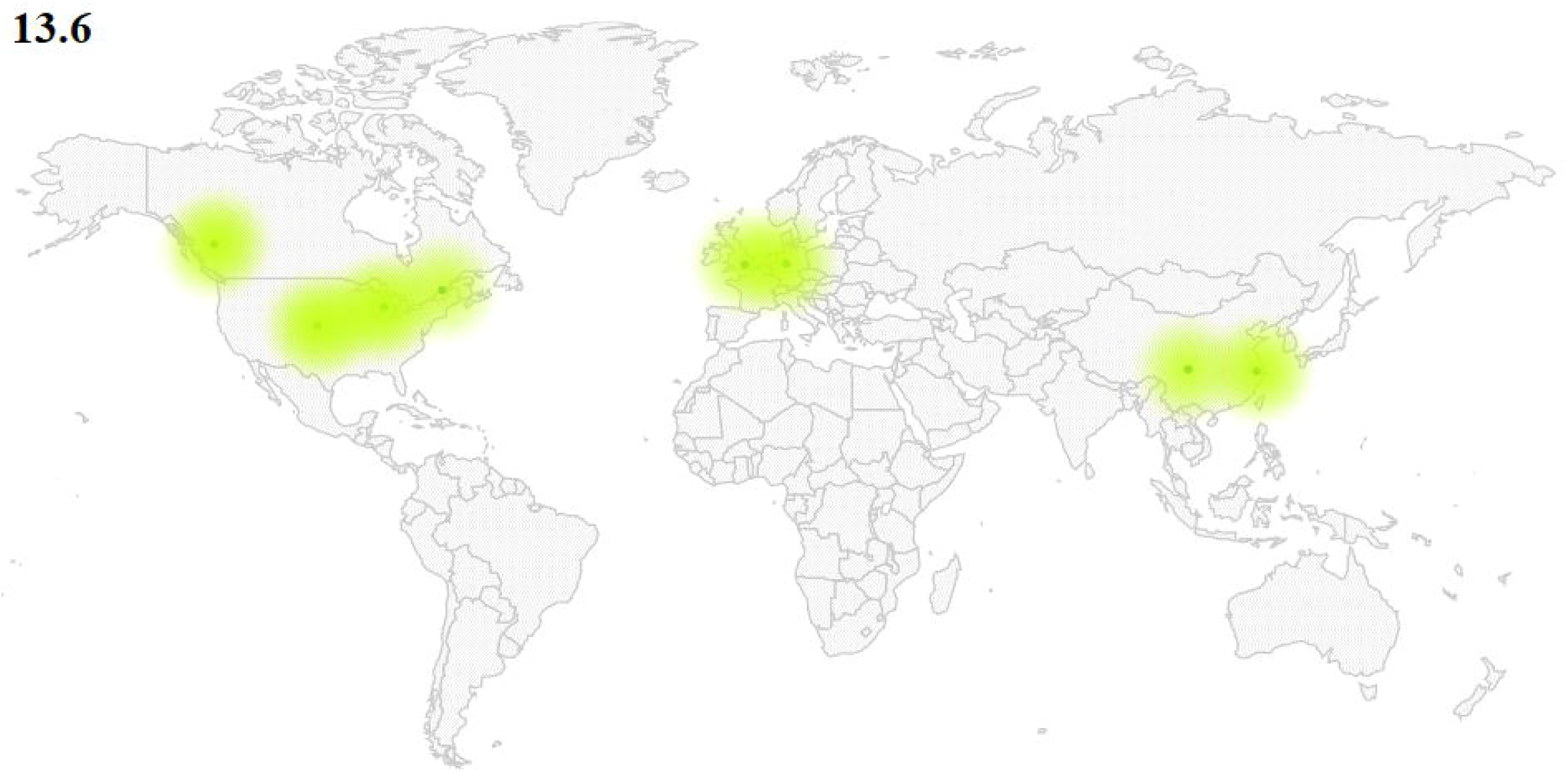

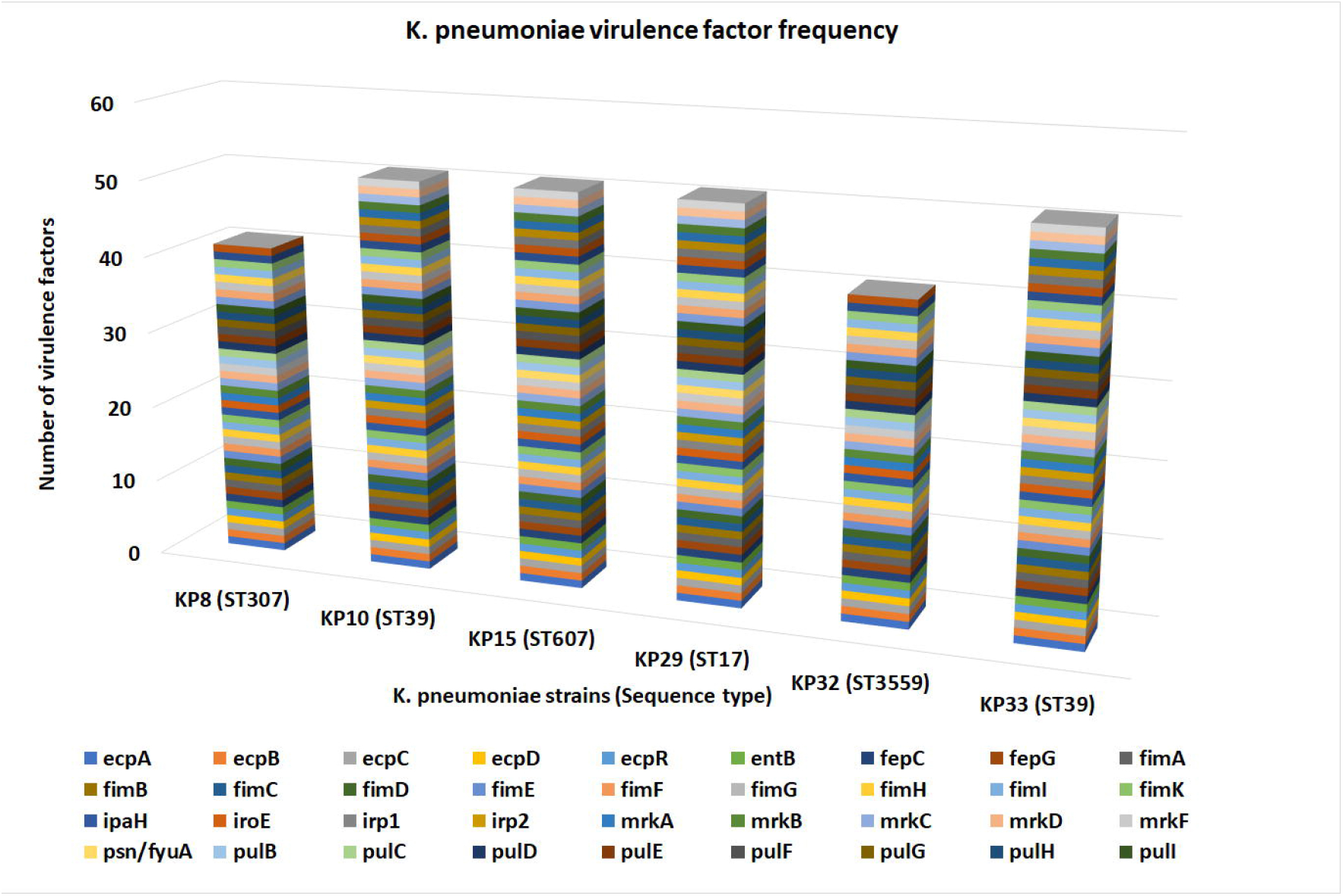

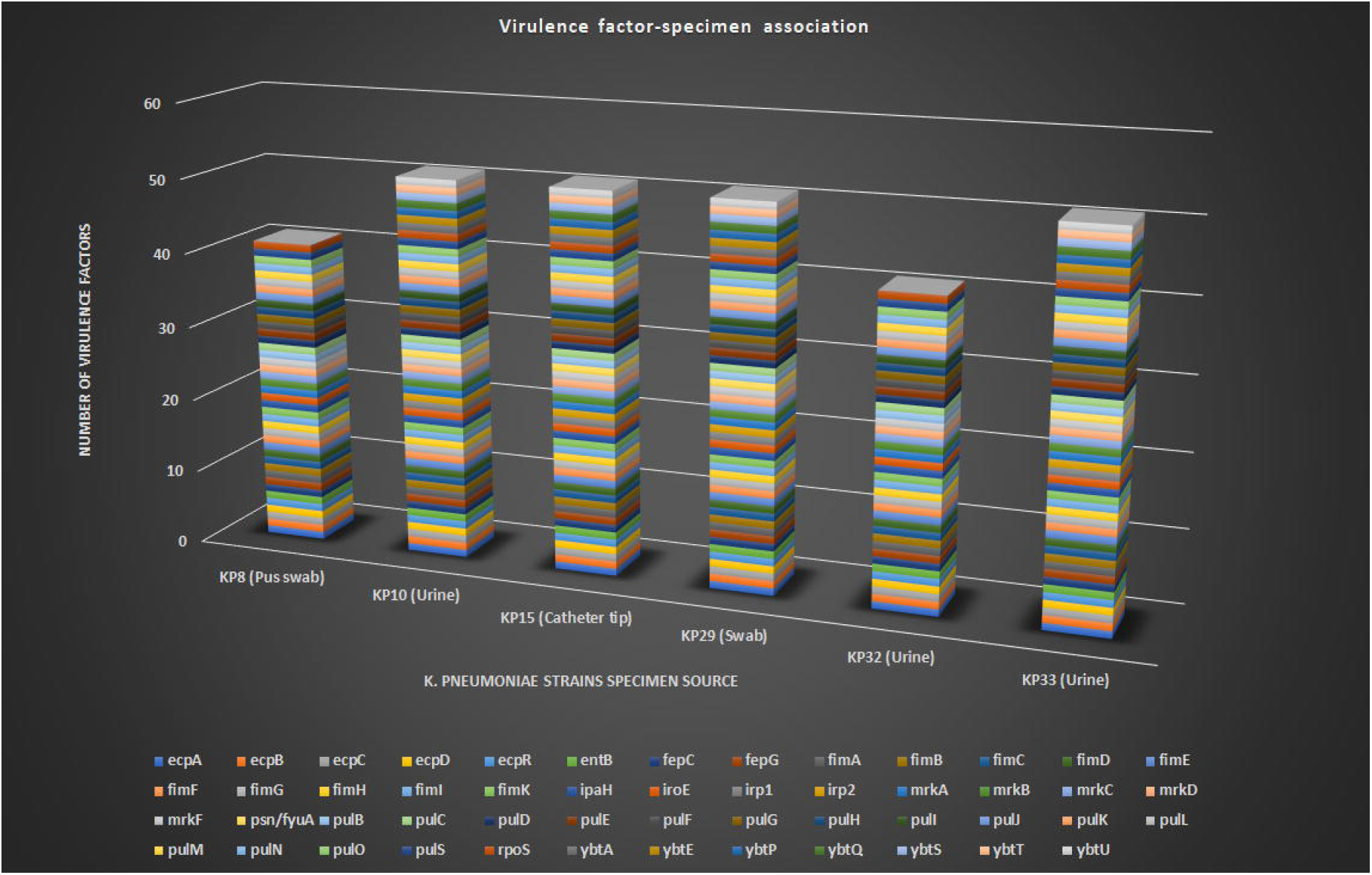

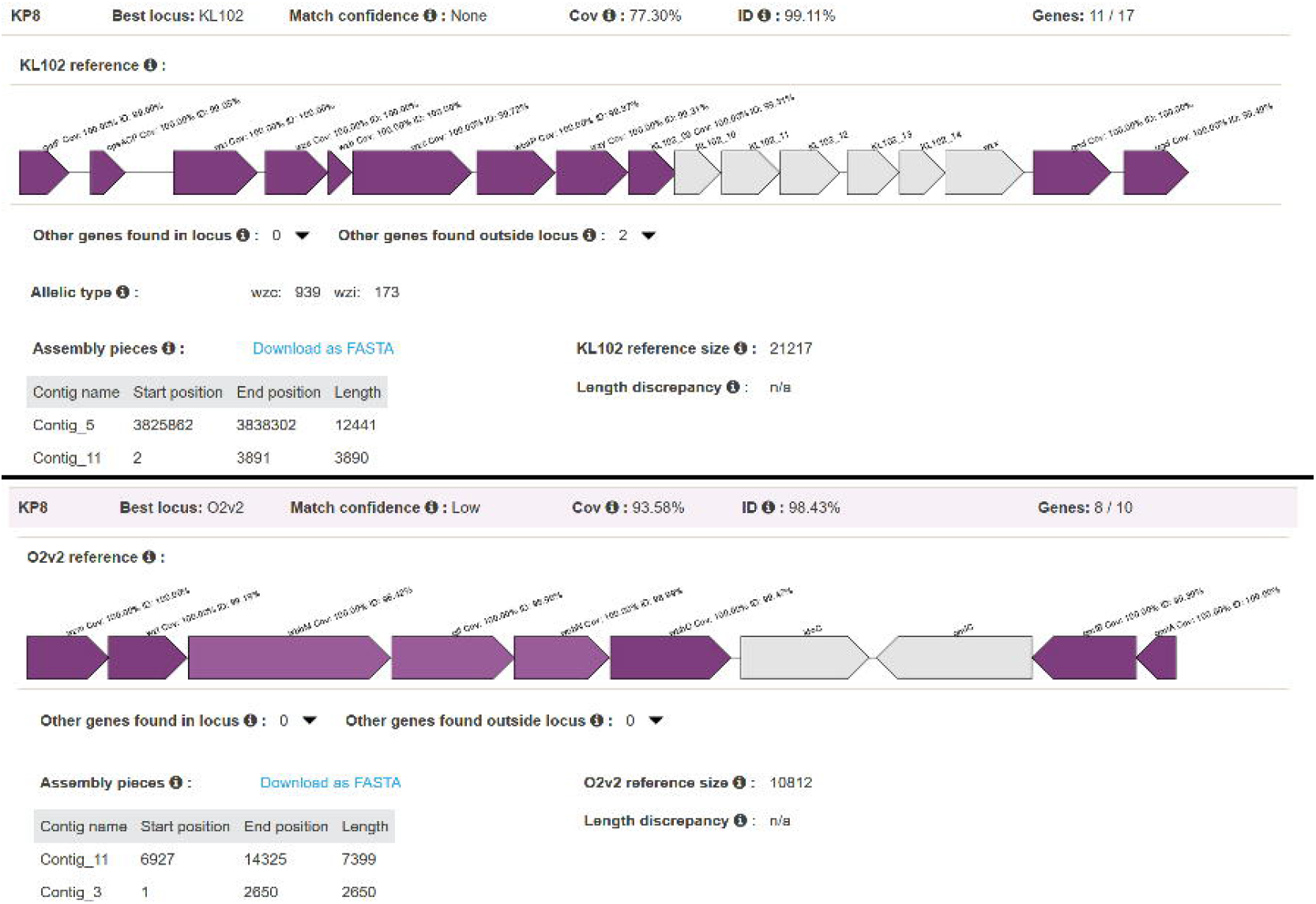

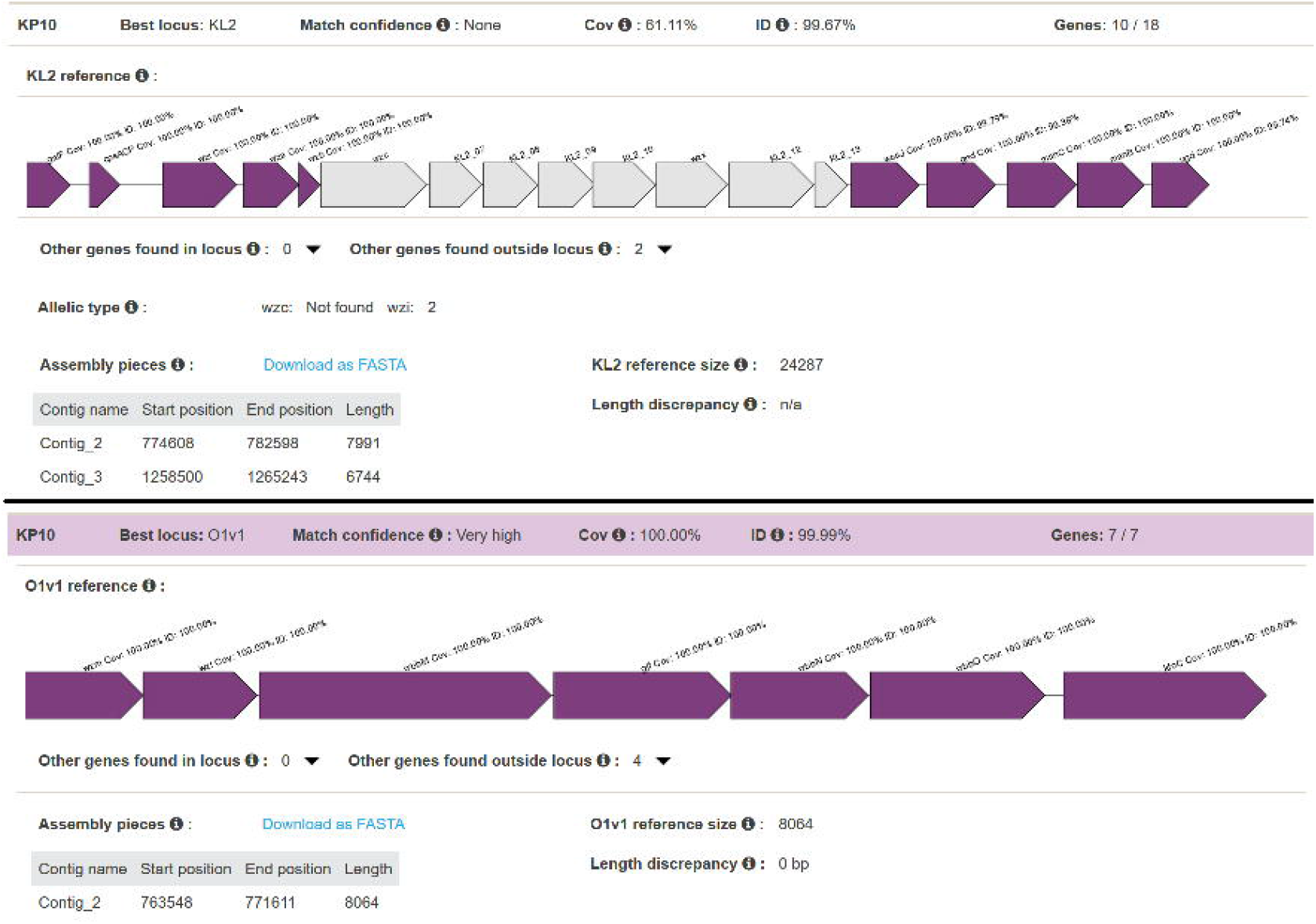

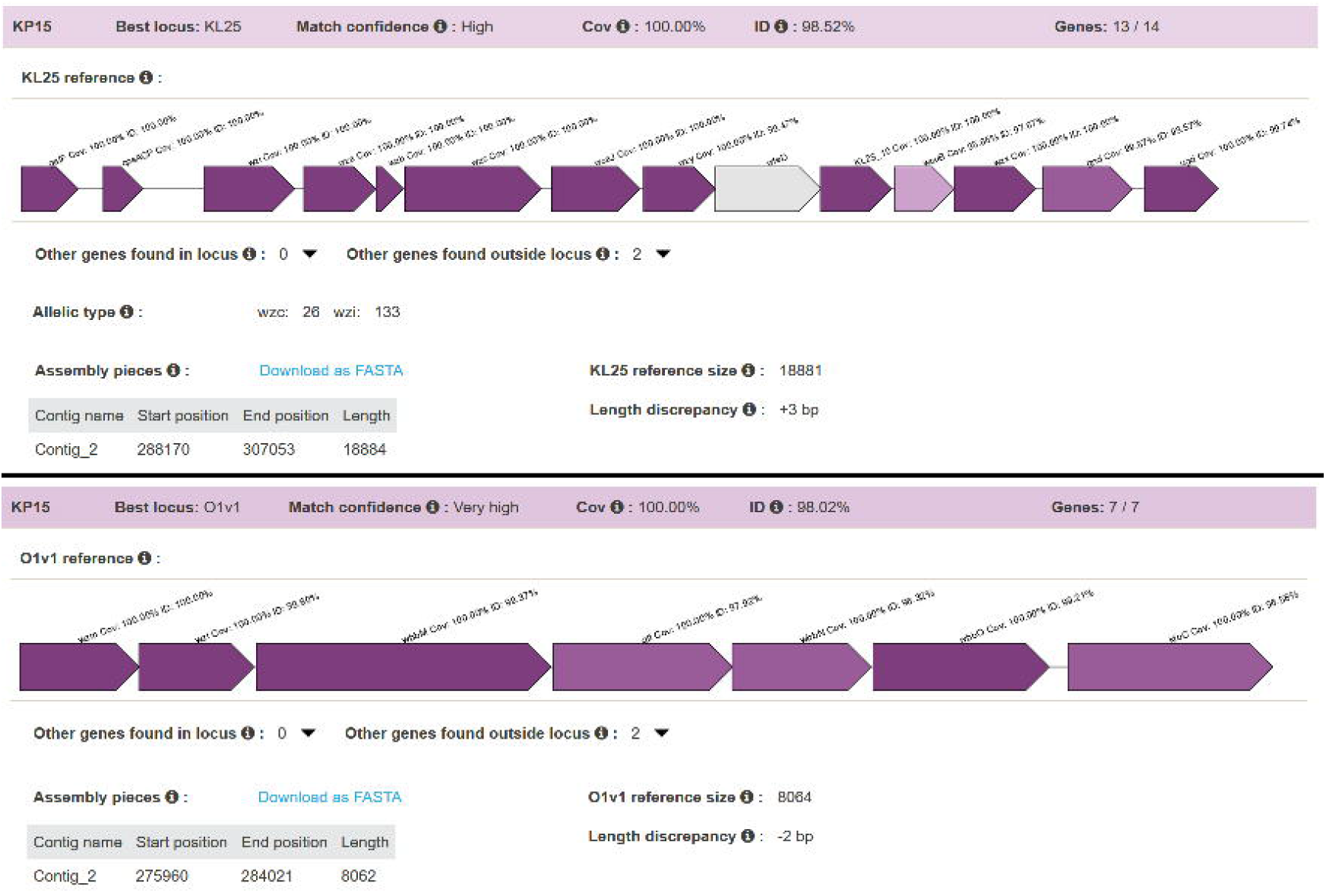

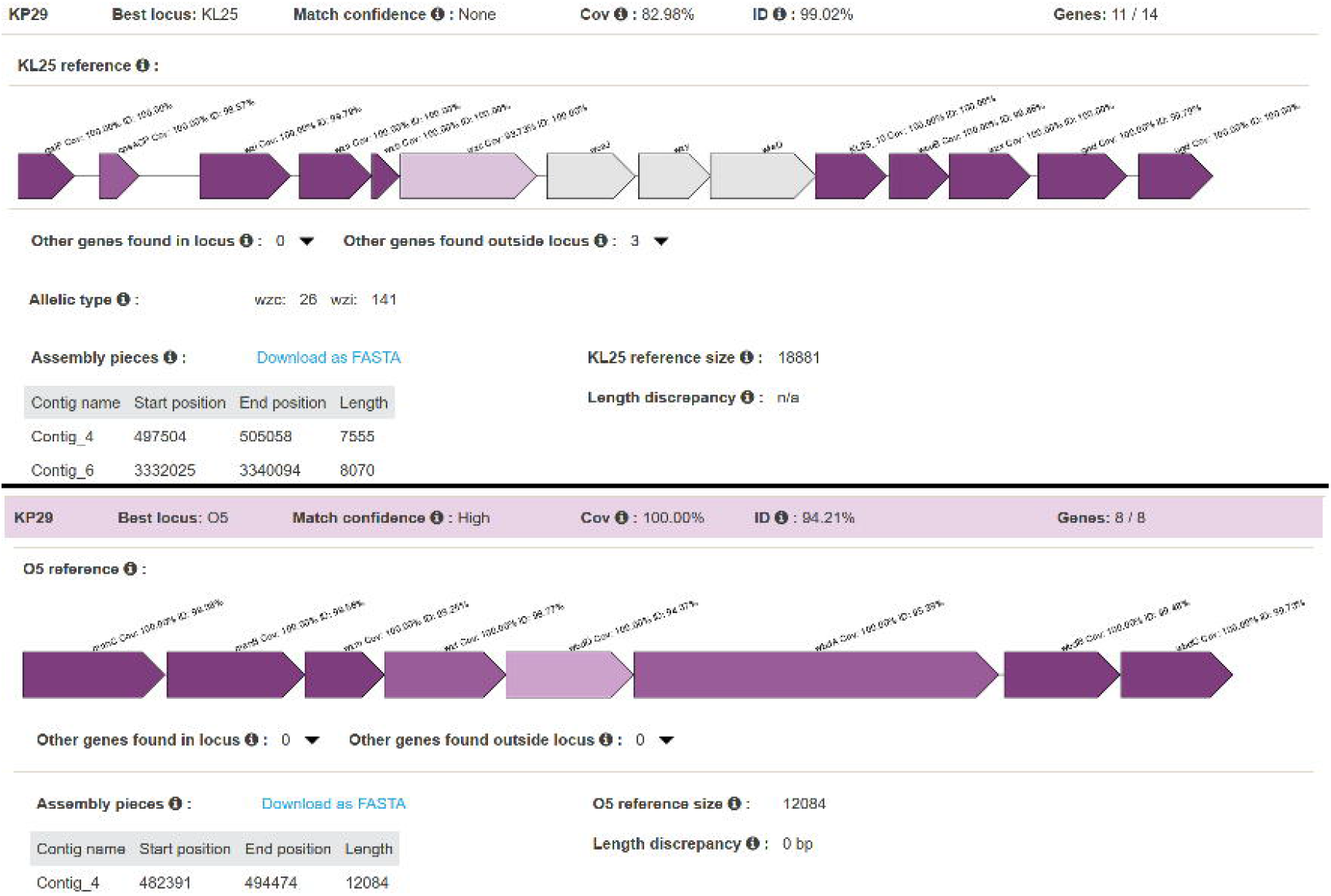

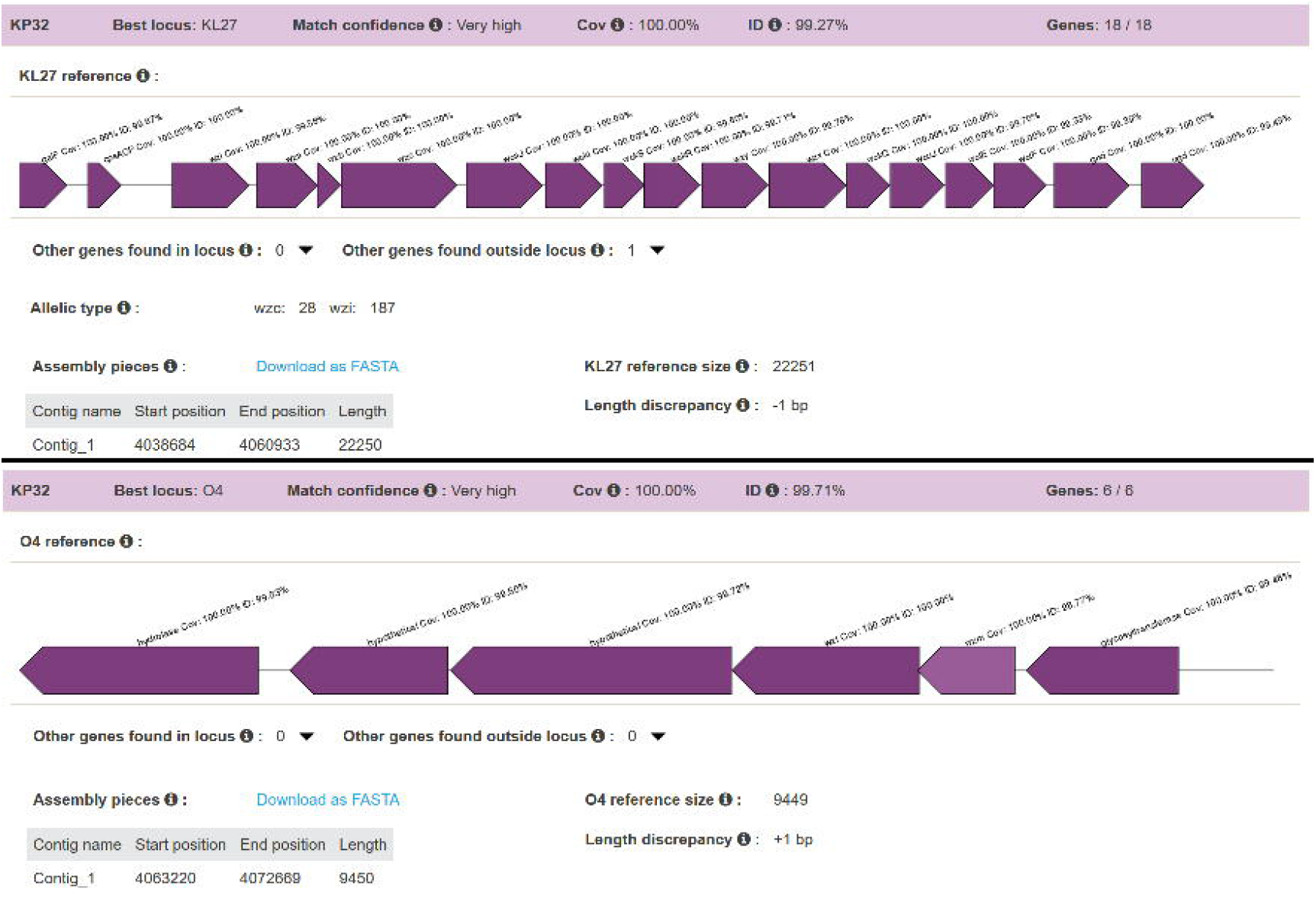

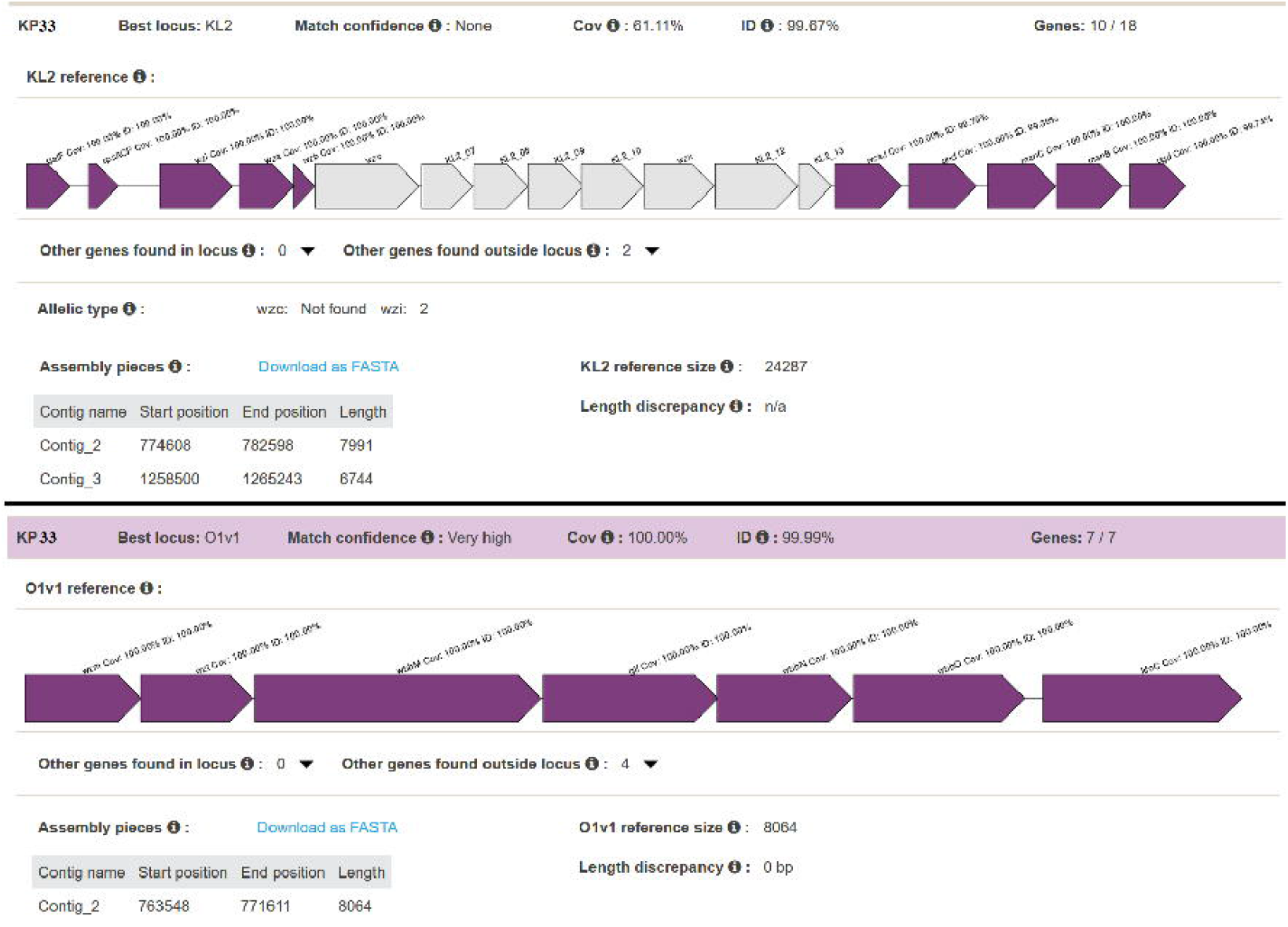

## Notes

### Competing Interest Statement

The authors have declared no competing interest.

### Funding Statement

This work was funded by the NHLS

### Author Declarations

Permission was for this study was obtained from the ethical review board of the faculty of health sciences, UP

